# Bridging microbiology and public health through simulation-based learning

**DOI:** 10.64898/2026.08.21.26361043

**Authors:** Kathryn C. Krupinsky, Denise Kirschner

## Abstract

Within our synchronous, online global health-focused upper-level microbiology course, we find that students struggle to translate learning to real-world applications. For examples, consider the recent measles outbreaks and major events such as the COVID-19 pandemic, which prompt many questions about how basic microbiological information is used by public health professionals. To address these points, we created a simulation-based curriculum that places students in an action role during an infectious disease outbreak. Our stand-alone curriculum walks through a historical measles outbreak that introduces outbreak investigation, community communication, and how these depend on microbiological knowledge. By using breakout groups, students have an opportunity to decide classifications, public messaging, and intervention metrics. We provide students with an outbreak investigation reference worksheet and interweave breakout rooms with didactic vignettes covering background information while revealing actual responses to outbreaks in conjunction with data obtained by responding scientists. Students synthesize material and apply it in real-time – allowing them to exercise critical thinking while bolstering relevance of microbiology and public health to popular media.

## INTRODUCTION

The modern college student is constantly exposed to media through traditional (i.e., TV, radio, print) and non-traditional (i.e., TikTok, Instagram) avenues. Media has also increasingly focused on topics relating to microbiology/public health partly in response to recent world events such as the 2020 COVID-19 pandemic, the 2022 M-pox epidemic, and the 2026 US cyclospora outbreak (1, 2). We anecdotally find that this coverage has inspired more students to come to our microbiology/public health classrooms with the aim of understanding why these events happen and how the scientific community contributes to media headlines. Further, we find that students want to learn how facts translate to headlines because, in their own lives, they are sometimes asked to comment on the microbiology discussed in the media. Within undergraduate courses, students are typically taught technical information with memorization-based assessments (3–5). Some classes include abstract discussion of how science is used to inform public policy actions as well as the interventions that are discussed and debated within the media. Nonetheless, often, students complete a course without enough information to begin to understand how technical information translates into headlines that they consume from media.

One major challenge of classical undergraduate education is the volume of information necessary to cover within a semester (3–7). Because of reasonable program and course learning outcomes, time constraints often do not allow for making connections between technical information and popular media. Further, making these connections is not trivial. Because translating basic biology to public health policy and interventions requires multiple disciplines and some knowledge of each (8). For example, to enact a “simple” public health intervention such as seasonal influenza vaccination, there must be a coordinated effort between virologists, immunologists, bioinformaticians, pharmaceutical manufacturers, public health communicators, politicians, doctors, and many more individuals (9). Multi-disciplinary courses are rarely taught and the tools to teach translation from classroom to applications are limited (10, 11). Given its complexity and the paucity of teaching opportunities and tools, it is no wonder that students struggle to grasp how the technical information that they learn within the classroom is translated into messaging within popular media.

One way to begin addressing this lack of connection between technical facts and implementation is to integrate real-world data into the classroom. This content can be incorporated through brief, targeted activities, such as scientist spotlights, disease highlights connected to unique mechanisms, or assigned popular article readings (12–15). However, we anecdotally find that these infrequent and isolated integrations insufficiently addressed the larger issue. Therefore, we sought to find an alternative teaching approach that puts students into the “driver’s seat” to see firsthand how basic biological information can be used to later create headlines in the media.

Simulation-based learning is a teaching technique that uses guided, immersive scenarios to recreate job-realistic situations within a controlled classroom environment. (16, 17). This technique is often used within allied-health classrooms where students are not only asked to work through a relevant scenario (i.e., treating a patient), but also are presented with additional elements that augment situations from contrived classroom examples to real life (i.e., working with an actor) (18, 19). These simulation-based learning experiences are preferred to real experiences since they provide an opportunity for an instructor to give live feedback on scenario progress in a low to no risk setting (17). Additionally, prior studies show that this approach is more effective and better prepares students to apply content learned in one course to other course and ultimately how this relates to their careers (17, 18, 20).

Here, we present a stand-alone simulation-based curriculum of an infectious disease outbreak investigation using a real-life historical measles outbreak. We model our curriculum on the training that U.S. Center for Disease Control and Prevention (CDC) Epidemic Intelligence Service (EIS) officers complete (21). In this curriculum, we ask students to take on the role of public-health professionals and work with their peers to complete the key steps utilized during an outbreak investigation and its management (22). Our curriculum is unique in that it also integrates crisis communication training and messaging practices. Initial field testing within a fully virtual, synchronous classroom shows that this curriculum is highly engaging and effective in accomplishing stated learning objectives. Further, our curriculum has shown success in increasing understanding for students about the role of basic biology within outbreak investigations and the headlines they see within the media. We support these claims through administration of pre- and post-surveys and analysis of written responses in our initial field testing as detailed below.

### Intended audience and prerequisite student knowledge

The intended audience for this curriculum is biology/public health undergraduate and master’s students at 4-year degree-seeking institutions, although it could easily be adapted for advanced high school students, students at 2-year degree-seeking institutions, or doctoral level programs. For initial field testing, this curriculum was part of a larger course that is open to upperclassman and master’s students that span all biology-related majors. The course has a key focus of advancing students’ perspectives and understanding of how infectious diseases shape not only health, but also economics, politics, war, culture, social structures, and the course of human history a both local and global levels. Students in this course typically complete are undergraduate microbiology and immunology courses. While we designed the curriculum for this specific course, it would also be appropriate for any courses that would benefit from discussion of outbreak investigation. Throughout the semester, students are expected to be familiar with basic public-health principles, although no specific instruction of field epidemiology (outbreak investigation) is assumed. Students are also generally aware of major infectious diseases that may be relevant to outbreak curricula. Yet, no in-depth prior knowledge is necessary for successful completion of the curriculum.

### Curriculum delivery and learning time

This curriculum is self-contained and can be completed within a single 120-min class session.

### Learning objectives

After completion of this curriculum, students should be able to (LO1) summarize key steps of outbreak investigation and assess relative importance of each, (LO2) compose a single overarching communications outcome (SOCO) with limited information, (LO3) recognize components of and be able to collaborate with others to construct a good case definition, and (LO4) explain the role of communication and publication after completion of an outbreak for the field of public health.

## PROCEDURE

### Materials

This curriculum requires that all students have access to a synchronous presentation and an ability to be placed into smaller collaborative groups. Initial field testing accomplished this with screen-sharing and breakout rooms within video-conferencing software (e.g., Zoom) for a virtual class session. This, alternatively, could be accomplished with a projector and movable tables for an in-person (F2F: Face to Face) class session. Prior to the class session, students should be given access to the outbreak notes worksheet. Following the class session, students should be provided with their own copy of the presentation and the follow-up information sheet. If formal assessment of student learning is desired, a pre- and post-lecture survey may be administered (see **Suggestions for determining student learning** for complete details). All materials discussed (presentation slides, worksheet, follow-up sheet) are provided within **SI 1-3**.

### Student instructions

Prior to the class session, students should download or print the provided outbreak notes worksheet (**SI 2**) and be prepared to participate actively in small groups. Given the intense collaborative and somewhat experimental nature of necessary participation, it is recommended that students have some familiarity with their classmates and are comfortable volunteering information vocally throughout the class session. During the class session, students are asked to follow along with the presented slides by taking notes on the provided worksheet. While some information may be predicted from reading through the worksheet or by prior knowledge of the case, we ask that students to not jump ahead or try to reveal information prior to that point of presentation within the curriculum. Following completion of the class session, students should view follow-up information and (if desired) read about the actual historical proceeding of the presented outbreak.

### Instructor instructions

#### Curriculum rationale and case selection

This curriculum is designed to present students with information about a real-life outbreak and invite them to examine questions and situations that are faced by public-health professionals. Thus, we chose a case that covers important and, at times, potentially controversial topics vital to modern public health conversations. Our goal is to expose this material first in a classroom setting so that we can provide the necessary safety for students to explore these topics and, in turn, to empower students to have these conversations within their everyday life.

The case chosen for this curriculum was a measles outbreak that originated in Hennepin County, Minnesota, USA in 2017. The complete outbreak proceedings can be found in (23). We will briefly detail key components here. In this outbreak, a total of 65 cases were confirmed with thousands of other close contacts identified. Investigation into patterns of cases revealed that most cases were in unvaccinated individuals of Somali-American descent. Based on community conversations, the low vaccination coverage was determined to arise from misinformation about the link between the MMR vaccine (a vaccine that confers protection against three pathogens, including measles) and autism.

This case was chosen because of the recognizability of the pathogen of concern (measles), the clearly identifiable pattern of cases, and the associated conversations that led to the final control strategy. While we use data that were published in association with this outbreak, we re-analyzed and processed these data in a way that is digestible to students and allows for a clear picture of how field epidemiology principles can be applied.

#### Curriculum detailed overview

This curriculum follows the basic steps of field epidemiology as outlined in (22, 24). Briefly, the steps are as follows: [1] prepare for fieldwork, [2] confirm the diagnosis, [3] determine existence of an outbreak, [4] identify and count cases, [5] determine person, place, and time, [6] develop and test hypotheses, [7] implement and evaluate control measures, and [8] communicate findings. The curriculum begins with an overview of the role that students will assume while going through the curriculum material. From there, the presentation slides switch between didactic material (shown as slides with a blue background) and outbreak-specific information (shown as slides with a white background). The curriculum features 6 breakout opportunities that are indicated on the slides (shown as statements with red text and a question mark icon). Suggested time periods for each breakout opportunity are stated on the slides. Following each breakout opportunity, the next consecutive slide shows logical follow-up information or answers to what was implemented during the historical outbreak. The curriculum concludes with recent outbreak news as discussed below.

#### Communications component

Given the aim of this curriculum to allow students to better understand translation of technical facts to media headlines, we integrate communication training directly into the curriculum. We do this by having one breakout opportunity where students must interact with a fictional local reporter asking questions about the outbreak. To guide their interactions, we teach specific guiding principles recommended by the World Health Organization (WHO) to construct messaging called the SOCO or <u>S</u>ingle <u>O</u>verarching <u>C</u>ommunication <u>O</u>utcome (25). The SOCO is a data-based communication principle that guides users to construct messaging by thinking about what the intended audience is and the specific actions desired by the audience in response to the messaging (26). We place this breakout session near the end of the curriculum to ensure students have an appropriate amount of biological/public health background information to inform their messaging. However, we also acknowledge and emphasize that there is not always an opportunity to have complete information prior to engagement with the media in emerging public health situations (27).

#### Problem-solving component

A secondary goal of the curriculum is to provide students opportunity to participate in problem-solving with real data. In our curriculum, we do this by presenting pre-processed data and giving students the opportunity to form their own conclusions. By withholding the answers, students are invited to be brave and use their problem-solving skills to present an understanding of the data. We pair this with introduction of novel concepts throughout didactic components of the curriculum to ensure students are not lost or too frustrated with the lack of a clear-cut answer to the presented questions.

#### Preparation for class

If not familiar with field epidemiology, we recommend that the instructor familiarize themselves with the general outbreak procedure by reading case reports (found within the CDC’s MMWR or other similar resources, see **SI 3**) or working through additional resources of field epidemiology. While external preparation is recommended, all necessary structure and information for execution of this curriculum can be found exclusively within the provided presentation slides (**SI 1**).

#### Tailoring of curriculum

Following completion of all steps of the outbreak investigation, there is space for inclusion of recent outbreak news within an instructor’s local community. We encourage instructors to update these slides immediately preceding the presentation of the curriculum with current outbreak news (found through a search engine query) to spur discussions with information that students may have recently consumed within their everyday life.

#### Curriculum implementation

During this curriculum, there are several moments where students are asked to work in small groups to construct various messaging or analyze presented data. For virtual presentation (as has been done in initial field testing), we recommend the use of breakout rooms to allow for optimal teamwork and collaboration. The instructor should be comfortable setting up breakout rooms or devising an alternative way to divide up the class into small (3-5 individuals per group) breakout activity groups prior to the class session. Finally, the instructor should share the worksheet (**SI 2**) with students and inform students that they need a way to access the worksheet and fill it out during the class session prior to arrival within the classroom space. We also suggest that the instructor rotate the group reporter through all members of each group to provide opportunities for everyone to practice leadership skills.

#### Post-curriculum follow-up

Following the class session, the instructor should disseminate slides and share follow-up information worksheets. Optionally, the instructor may ask students to fill out a reflection on both the curriculum and what they learned about the role of communication within public health/microbiology to further integrate the overarching message and motivation of the curriculum.

### Suggestions for determining student learning

Given the nature of the curriculum experience, we caution instructors about implementing strict evaluation on information retention. Instead, we suggest that instructors emphasize the overall process and team-based problem-solving skill building. However, if quantification is needed (as was the case in our initial field testing) we determine student learning using two sources of data in a timeline as outlined in **Figure 1**.

**Figure 1.**
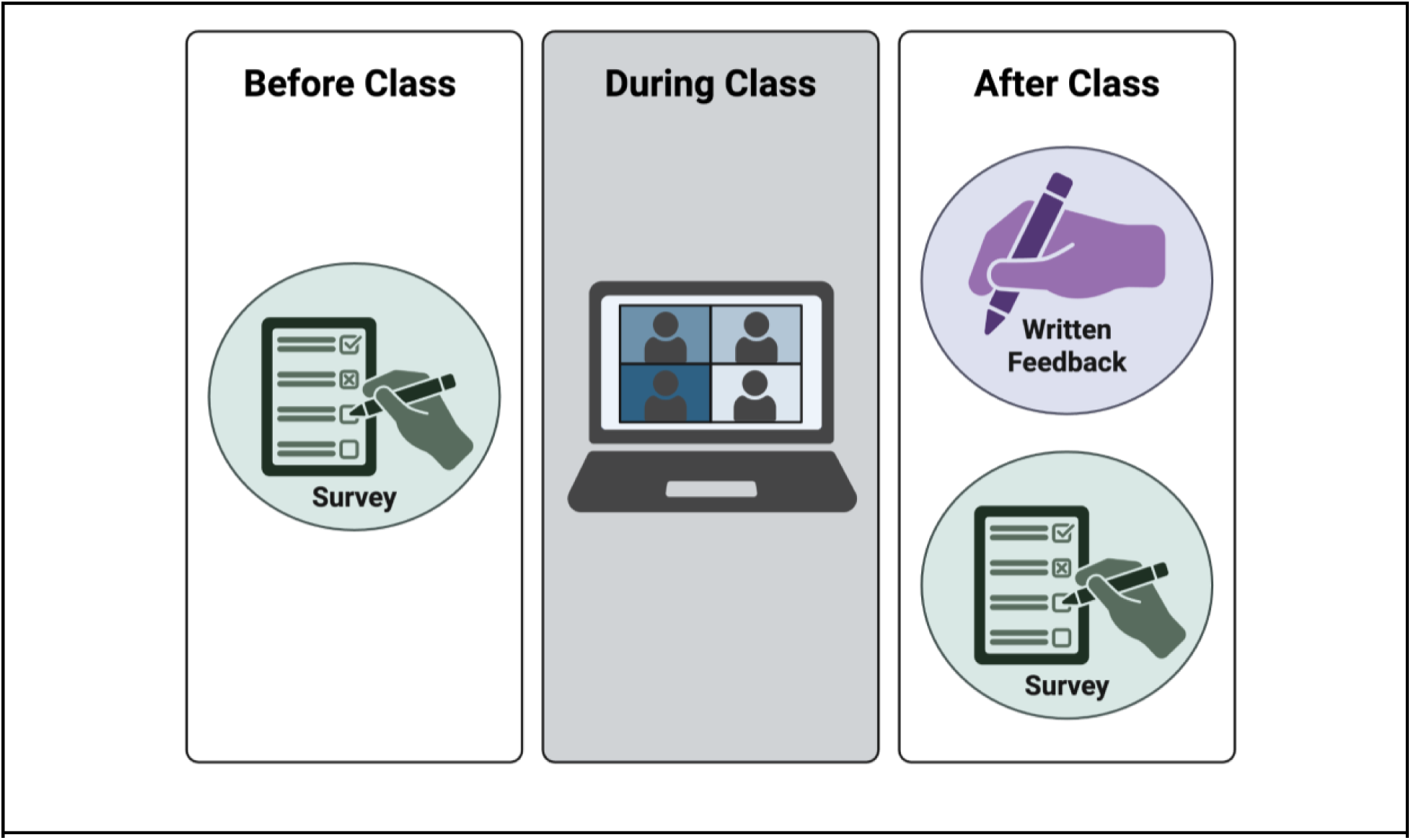
Curriculum and assessment design. Students were asked to complete two forms of feedback before and after the class session where the curriculum was presented. Before and after class (shown in green) students were asked to complete a short (5 question) survey with specific questions used in initial field testing shown in **Table 1**. Students were also asked to complete an after-class, long-form written feedback with the prompt: “WRITE A PARAGRAPH summarizing [the instructor]’s outbreak lecture format and [their] case. Also evaluate [their] presentation style.” This figure was created in Biorender.

**Table 1.** Pre- and post-curriculum survey questions. These questions were provided to students in the exact format as listed using institution’s learning management system (Canvas). For Q1 and Q4, bolded text indicated the correct answer. For Q2, Q3, and Q5, the question does not have a specific correct answer. Learning objectives were assigned based on investigator experience with specific learning objectives corresponding to numbers found in the **Learning Objectives** section of **Introduction**.

|  |  |
| --- | --- |
| **Learning Objective** | **Question and choices** |
| LO1 | Q1: Which of the following is NOT a step in outbreak investigation?<br>A. Determine existence of an outbreak<br>B. Identify and count cases<br>C. Communicate findings<br>D. Confirm diagnosis<br>**E. Develop and test a vaccine** |
| LO1 | Q2: Of the following, which is the most important step of outbreak investigation?<br>A. Decide on a working case definition<br>B. Identify and count cases<br>C. Implement control measures<br>D. Communicate findings<br>E. Confirm diagnosis |
| LO2 | Q3: On a scale from (not confident) 1 to 5 (highly confident), during an emerging outbreak, how confident would you be constructing messaging to speak to the media about details?<br>A. 1 (not confident)<br>B. 2<br>C. 3<br>D. 4<br>E. 5 (highly confident) |
| LO3 | Q4: Which of the following correctly describes the components of a good case definition?<br>A. Time, place, origin, clinical features<br>**B. Person, place, time, clinical features**<br>C. Person, time, origin, clinical features<br>D. Person, place, time, transmission route<br>E. Place, origin, clinical features, transmission route |
| LO4 | Q5: What is the role of communication and publication after completion of an outbreak for the field of public health? *[Free response]* |

For written feedback, students are asked to reflect on their experience with the curriculum and receive a grade based on completion. We include an analysis of this information within the sample data section below; however, accuracy or opinions were not included within grading criteria. For the survey, we implemented a short (1-5 question) pre- and post-survey that included a mix of multiple choice and free response questions. The questions posed to students for both the pre- and post-survey were identical and were administered within the institution’s learning management system (Canvas). Specific questions used and corresponding LOs are in **Table 1**.

### Safety issues

There are no known safety issues associated with this curriculum. The student feedback portion of this study has been reviewed and approved by the University of Michigan IRB (HUM00285792).

### Sample data

We analyzed both sources of data (survey and written feedback) to determine student achievement of learning objectives. All data was deidentified prior to initiation of analysis. Quantitative questions (Q1-4, **Table 1**) for the pre- and post-surveys were examined for changes in overall trends. Direct comparison of individual student pre- and post-survey responses were not available. Qualitative questions (Q5, **Table 1**) from the survey and written feedback were thematically coded with representative responses of each thematic code available in **Table S1 and S2**. Each response was given 1-5 thematic codes based on investigator analysis of the full written response. Specific codes were selected based on perceived sentiment of responses with similar codes collapsed based on relative frequency and uniqueness. Qualitative analysis was conducted manually using Microsoft Excel (v16.112). All quantitative data were analyzed, and all data graphs were generated using the R programming language (v4.5.2). Code to perform these analyses and create visualizations can be found at https://github.com/kkrupins/measlesSimulatedOutbreak.

The first questions of the survey address a student’s ability to successfully summarize key steps of outbreak investigation and assess relative importance of each (LO1). We find that students can more accurately identify steps included within the investigative process following exposure to the curriculum material (**Fig 2A**). We also find that students changed their choice of the most important step of the investigative process (**Fig 2B**) suggesting an alteration in their perception of relative importance of each. When asked to rate their confidence in constructing messaging to speak to the media about an emerging outbreak (LO2), we find that there is a significant increase (p < 0.01) in overall student scores when comparing pre- and post-survey answers (**Fig 2C**). In the pre-survey, a minority of students correctly select the correct components of a good case definition (LO3); meanwhile, the majority correctly answer this question in the post-survey (**Fig 2D**).

**Figure 2.**
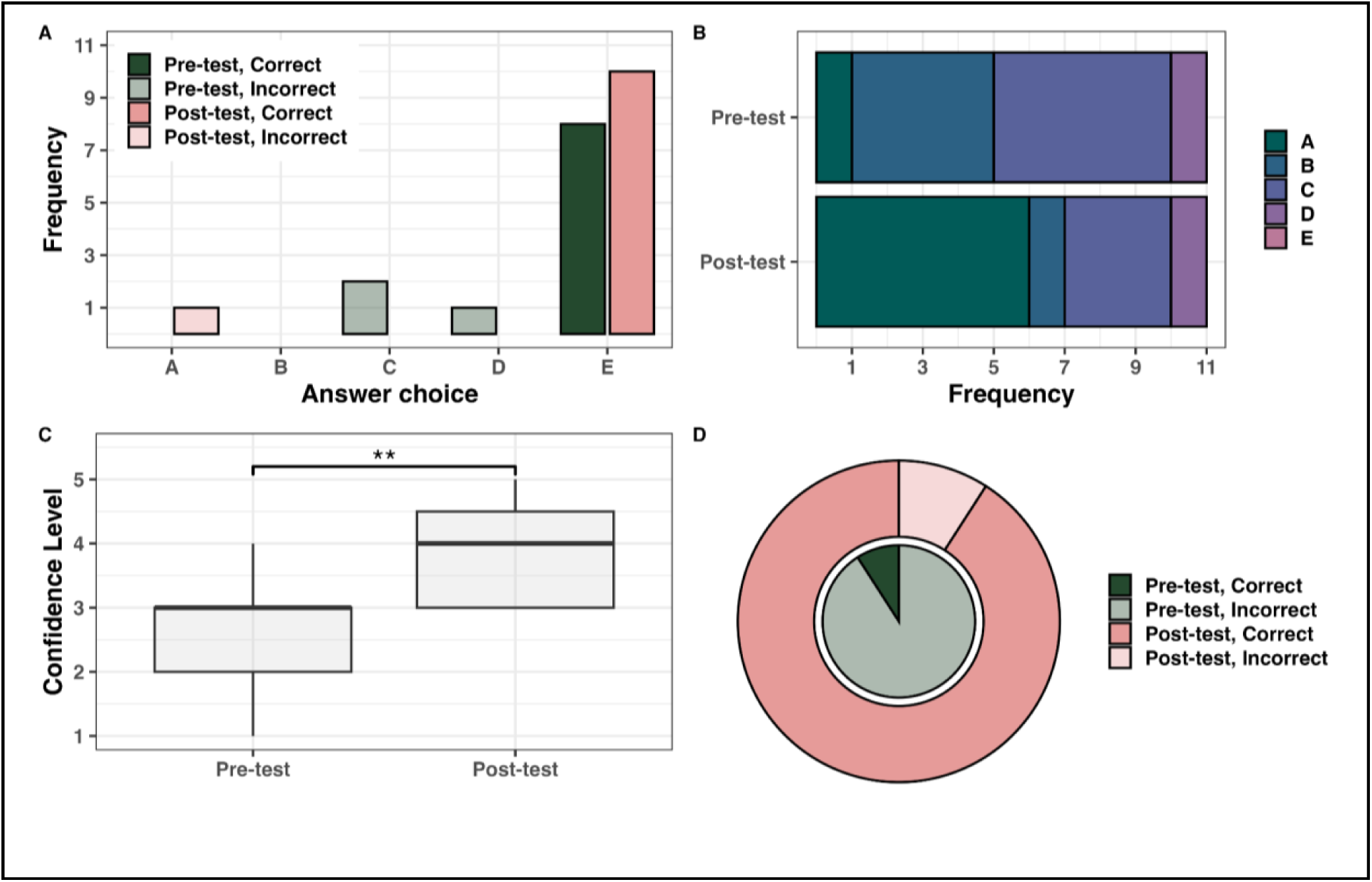
LO1-3 pre- vs. post-test results. Student pre- (n = 11) and post-survey (n = 11) response to (A) Question #1, “Which of the following is NOT a step in outbreak investigation?”, (B) Question #2, “Of the following, which is the most important step of outbreak investigation?”, (C) Question #3, “On a scale from (not confident) 1 to 5 (highly confident), during an emerging outbreak, how confident would you be constructing messaging to speak to the media about details?”, and (D) Question #4, “Which of the following correctly describes the components of a good case definition?”. For (C), significance was determined using a student’s t-test and alpha = 0.01.

The final survey question asked students to explain the role of communication and publication after completion of an outbreak for the field of public health (LO4). As expected, before exposure to the curriculum material, students primarily focus their responses on a handful of themes (transparency, public education, avoiding future outbreaks, and long-term improvement) easily accessible through non-technical exposure to public health. Following exposure to the curriculum material, we observe a diversification of opinions and thoughts about the importance of communication (**Fig 3**). Together, these data show that students have an increase in mastery of stated learning objectives following exposure to the outbreak curriculum material.

**Figure 3.**
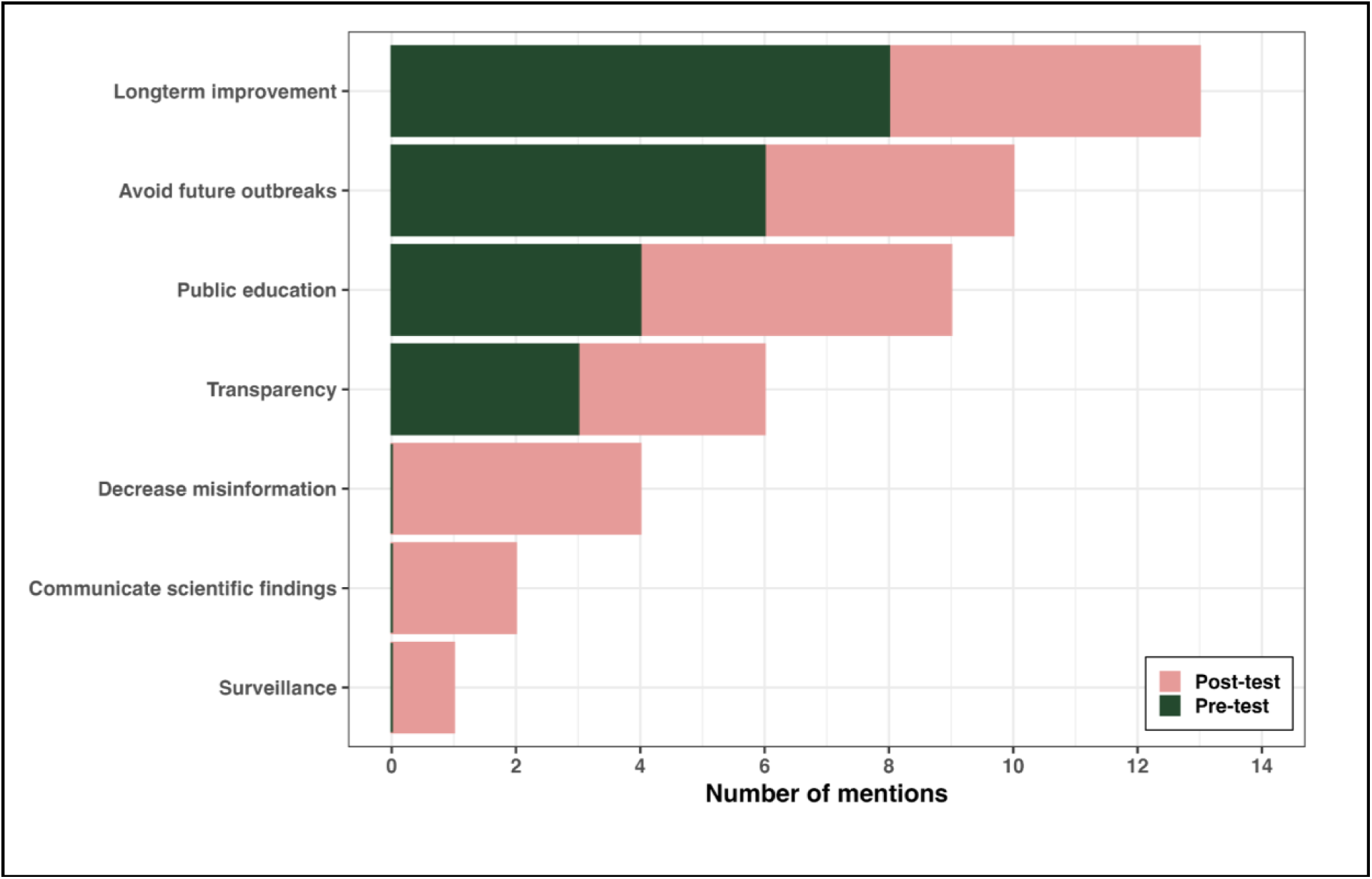
LO4 qualitative data results. Responses to the question: “What is the role of communication and publication after completion of an outbreak for the field of public health?”. N = 11 students.

In addition to learning objectives mastery, we are also interested in attitudes of students towards the curriculum material. We find that students indicate a strong preference of the curriculum, real-world practicality of the material, worksheet component, and lecturer enthusiasm (**Fig 4**). This suggests that students positively perceive the curriculum material and format in addition to learning stated learning objectives (as supported by the survey results, **Fig 2-3**).

**Figure 4.**
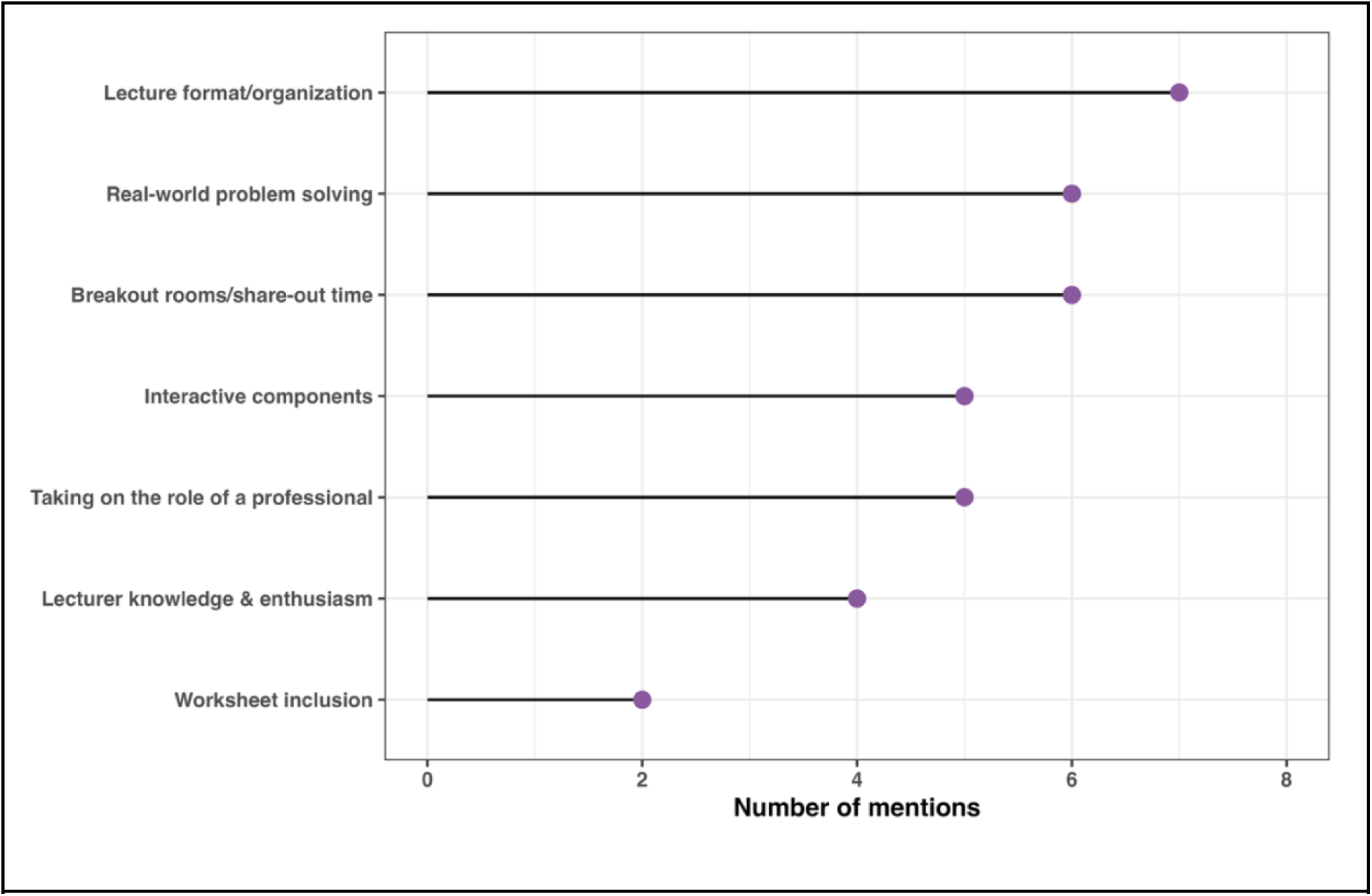
Written feedback results. Responses to the question: “WRITE A PARAGRAPH summarizing [the instructor]’s outbreak lecture format and [their] case. Also evaluate [their] presentation style.” N = 9 students.

## DISCUSSION

We designed a simulation-based learning experience that puts students into the “driver’s seat” to understand the interactions between basic biology/microbiology concepts, public health professionals, and the media during an infectious disease outbreak. In this stand-alone curriculum, students are guided through a historical measles outbreak and asked to complete fundamental field epidemiology tasks blindly (i.e., without prior knowledge of reality) such as construction of a case definition, analysis of epidemic frequency data, and responding to local reporters. Through initial field testing, we find that this curriculum was successful in increasing overall student understanding and achievement of stated learning objectives. We also find that students respond well to the innovative format of the curriculum and its ability to allow the student to actively participate and understand the roles they may take on within a workplace related to classroom material.

### Field testing

This curriculum has been implemented for two years within a small, virtual, synchronous learning environment. In this testing, the curriculum was implemented as a guest lecture and, aside from the title, students were not aware of the format or teaching style of the guest lecturer. Sample data are available only for the second year; however, the curriculum was similarly well received in both years of testing.

### Evidence of student learning

Initial field-testing data suggests that this curriculum is highly effective at increasing student understanding of learning objectives and is overall well received by the tested student population. Pre- vs post-survey changes (**Fig 2-3**) show an increase in single-answer question accuracy (**Fig 2A, D**) suggesting an increased ability to answer memorization-based questions. We also find that students more deeply think about questions relating to relative importance and the greater context of the materials as indicated by changes in answer distribution (**Fig 2B**). Students additionally indicate an increased confidence in completing curriculum-related tasks as evidenced by the changes in mean messaging construction confidence score (**Fig 2C)**. Qualitative responses (**Fig 3**) within the pre-versus post-survey show an increase in diversity of themes suggestive of a broadening of the perceived roles of professionals and overall role of public health messaging following exposure to the curriculum.

### Observations about curriculum format

Within written feedback, students repeatedly comment about their enjoyment of the curriculum format and the ability to engage in problem-solving behaviors (**Fig 4**). As instructors, we observe that the pace and success of this activity was highly dependent on comfort levels of students with each other and their willingness to engage in this experiential learning modality. For some students, this was a difficult learning experience because its success is dependent on active participation. However, for many, this was stated as one of their favorite class sessions of the semester. Overall, we find that the format of this curriculum is well-received and gives students a new positive experience with active learning modalities, in this case simulation-based curricula, as well as a new way to view their engagement with biology and its role within what they see in the media.

### Possible modifications

In the course used for initial field testing, each class session was 120-minutes and, likewise, our curriculum is formatted for this longer session. Even with the longer class session, time management is a difficult feature of the curriculum format especially when lively breakout discussions occurred. To address this, we found that removing some of the later breakout opportunities or limiting the number of responses to be very helpful. If the course has shorter sessions, this curriculum could be sectioned for two separate class sessions with the curriculum being broken in the middle or with all didactic background material placed in the first session and the actual case placed in the second. Alternatively, presentation of all didactic background material slides could be filmed and placed as pre-lecture required viewing material. In this case, we recommend constructing a short survey that confirms students have viewed this material given the vitality of having proper background disease and epidemiology information to successfully participate and solve the curriculum outbreak. Initial field testing of this curriculum has been restricted to a small, synchronous, virtual classroom; however, we posit that this curriculum could also be altered to be implemented in an in-person classroom through use of turning to partners instead of breakout rooms. If the class is larger, there may be reliance on instructional aides (i.e., undergraduate or graduate teaching assistants) to manage classroom engagement in an efficient manner. Alternatively, the instructor could implement polling software instead of necessitating large-scale feedback sessions that may be time-consuming or hard to optimally facilitate.

## Supporting information

This supplement contains the PowerPoint slides used for and to be provided to students following the class session.

This supplement contains the outbreak notes worksheet students should download or print prior to the class session.

This supplement contains the additional follow-up information students should be provided following the class session.

## Data Availability

All data produced are available online at https://github.com/kkrupins/measlesSimulatedOutbreak.

## Acknowledgements

We thank University of Michigan Department of Microbiology and Immunology for their support and provision of the course to implement this innovative curriculum. KK is financially supported by a Rackham Predoctoral Fellowship from the Rackham Graduate School at the University of Michigan. Lastly, we thank the students who have graciously participated in this study and whose enthusiasm inspires our teaching.

## SUPPLEMENTAL TABLES

**Supplemental Table 1.** LO4 qualitative data results representative responses. Selected responses and corresponding codes to the question: “What is the role of communication and publication after completion of an outbreak for the field of public health?”

|  |  |
| --- | --- |
| **Theme** | **Representative quote(s)** |
| Long-term improvement | “...to provide an unbiased account of the events that took place during the outbreak to guide response to future outbreaks” |
| Avoid future outbreaks | “...to keep them informed on how to best recognize symptoms and other features to prevent future outbreaks” |
| Public education | “The goal should be to inform the public about the disease and implement education on the outbreak so that the public can know how to protect themselves.” |
| Transparency | “...allows for transparency to the public about the events, which also acts as a means of educating readers...” |
| Decrease misinformation | “To educate people about the outbreak and dispel any myths or misinformation” |
| Communicate scientific findings | “...help public health officials share what was learned about the disease, including how it was spread and how it was controlled.” |
| Surveillance | “It also allows for long term data on outbreak rates of certain pathogens by year, allowing for the monitoring of new emerging strains.” |

**Supplemental Table 2.** Written feedback responses. Selected responses and corresponding codes to the question: “WRITE A PARAGRAPH summarizing [the instructor]’s outbreak lecture format and [their] case. Also evaluate [their] presentation style.”

|  |  |  |
| --- | --- | --- |
| **Number of references (%)** | **Theme** | **Representative quote(s)** |
| 7 (20.0) | Lecture format/ organization | “This format helped me recognize where my reasoning was incorrect and how I could improve my approach” “I thought that this lecture was super fun and different from any lecture I’ve been in.” |
| 6 (17.1) | Real-world problem solving | “I really enjoyed being able to actively participate in coming up with solutions to a real-world problem.” “I liked the way we didn’t know everything at the beginning, since that’s what it’s like as a real EIS officer” |
| 6 (17.1) | Breakout rooms/ share-out time | “Being allowed to engage with classmates in smaller group discussions really drilled the information in.” “I liked that we stayed within the same team...and then sharing as a whole group, it was almost like a conference with coworkers.” |
| 5 (14.3) | Interactive components | “...took more of a ‘put yourself in the shoes of a researcher’ approach and prompted the class to think about potential resources and solutions for solving infectious disease outbreaks.” |
| 5 (14.3) | Taking on the role of a professional | “...helped me think through the problem like real public health investigators.” “In a way, it felt like I was actually participating in the outbreak investigation myself.” |
| 4 (11.4) | Lecturer knowledge & enthusiasm | “I really liked [the lecturer’s] presentation style because you could also tell [they were] genuinely interested and enthusiastic about the topic...” |
| 2 (5.7) | Worksheet inclusion | “The epidemiology notes also made the experience interactive because there was space to write my own thoughts and how I would investigate the case” |

