## Supplementary material for "Bridging microbiology and public health through simulation-based learning": This supplement contains the PowerPoint slides used for and to be provided to students following the class session.

### Slide 1
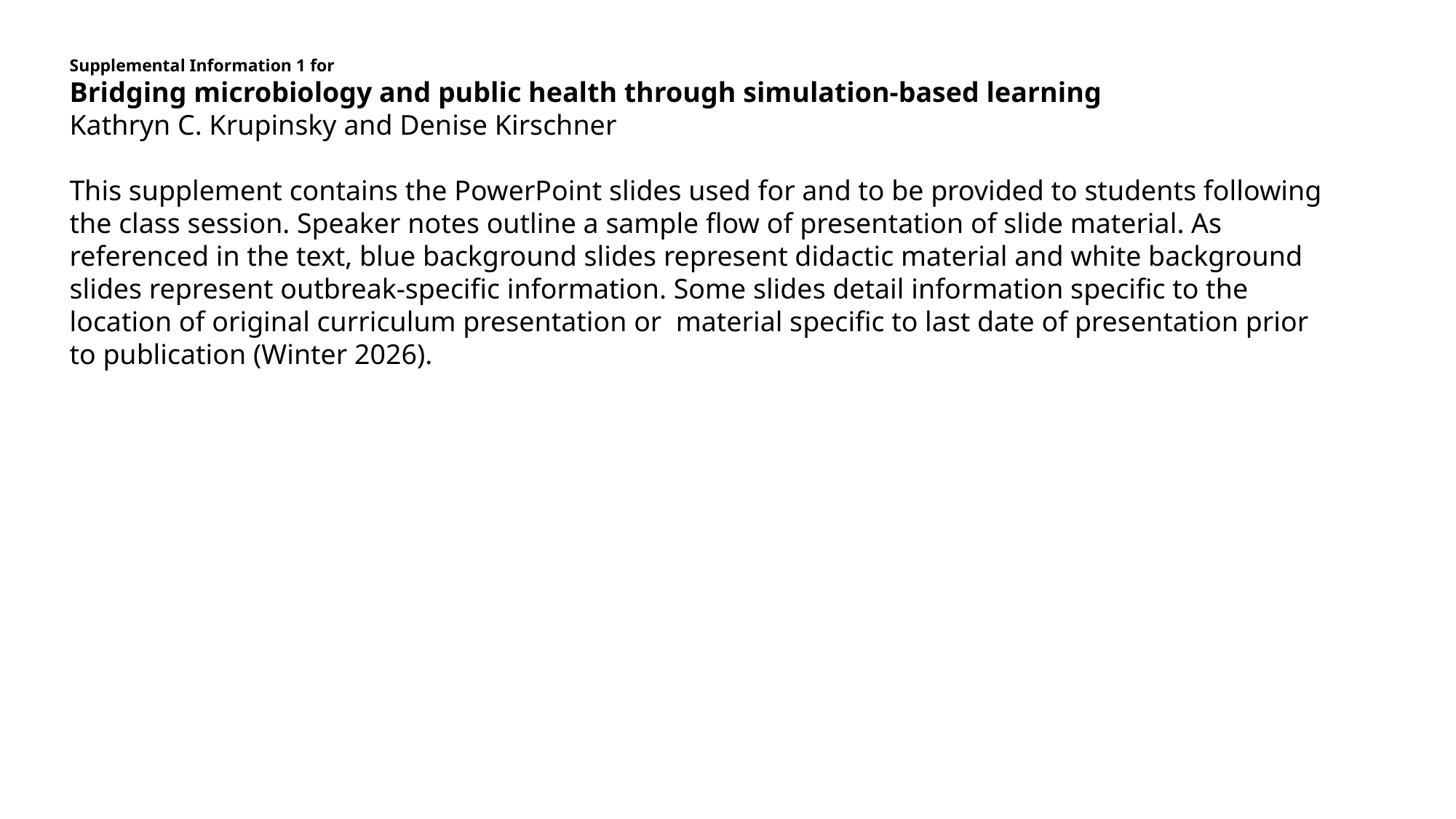

Supplemental Information 1 for
Bridging microbiology and public health through simulation-based learning
Kathryn C. Krupinsky and Denise Kirschner
This supplement contains the PowerPoint slides used for and to be provided to students following the class session. Speaker notes outline a sample flow of presentation of slide material. As referenced in the text, blue background slides represent didactic material and white background slides represent outbreak-specific information. Some slides detail information specific to the location of original curriculum presentation or material specific to last date of presentation prior to publication (Winter 2026).

### Slide 2
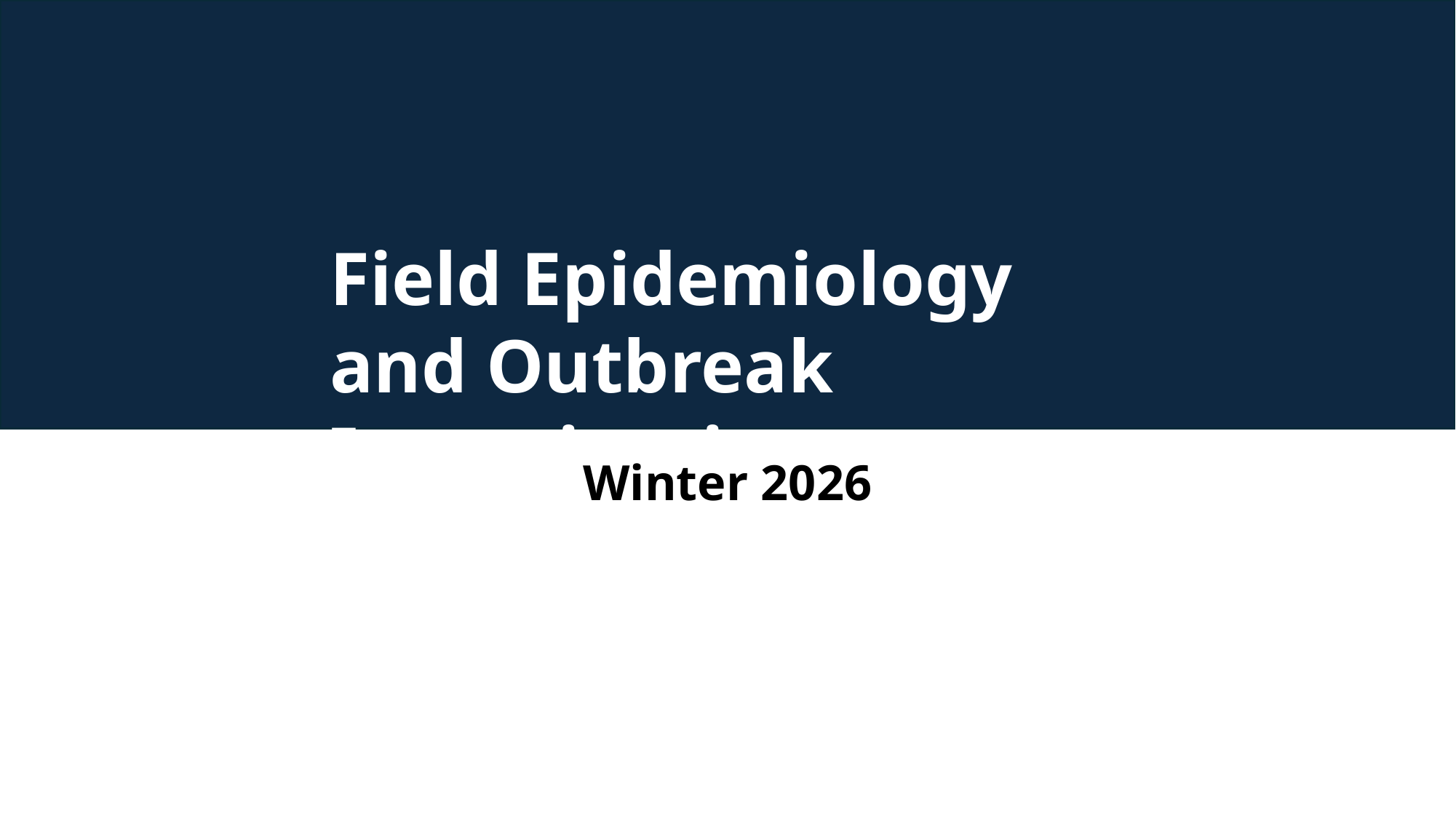

Field Epidemiology and Outbreak Investigation
Winter 2026

### Slide 3
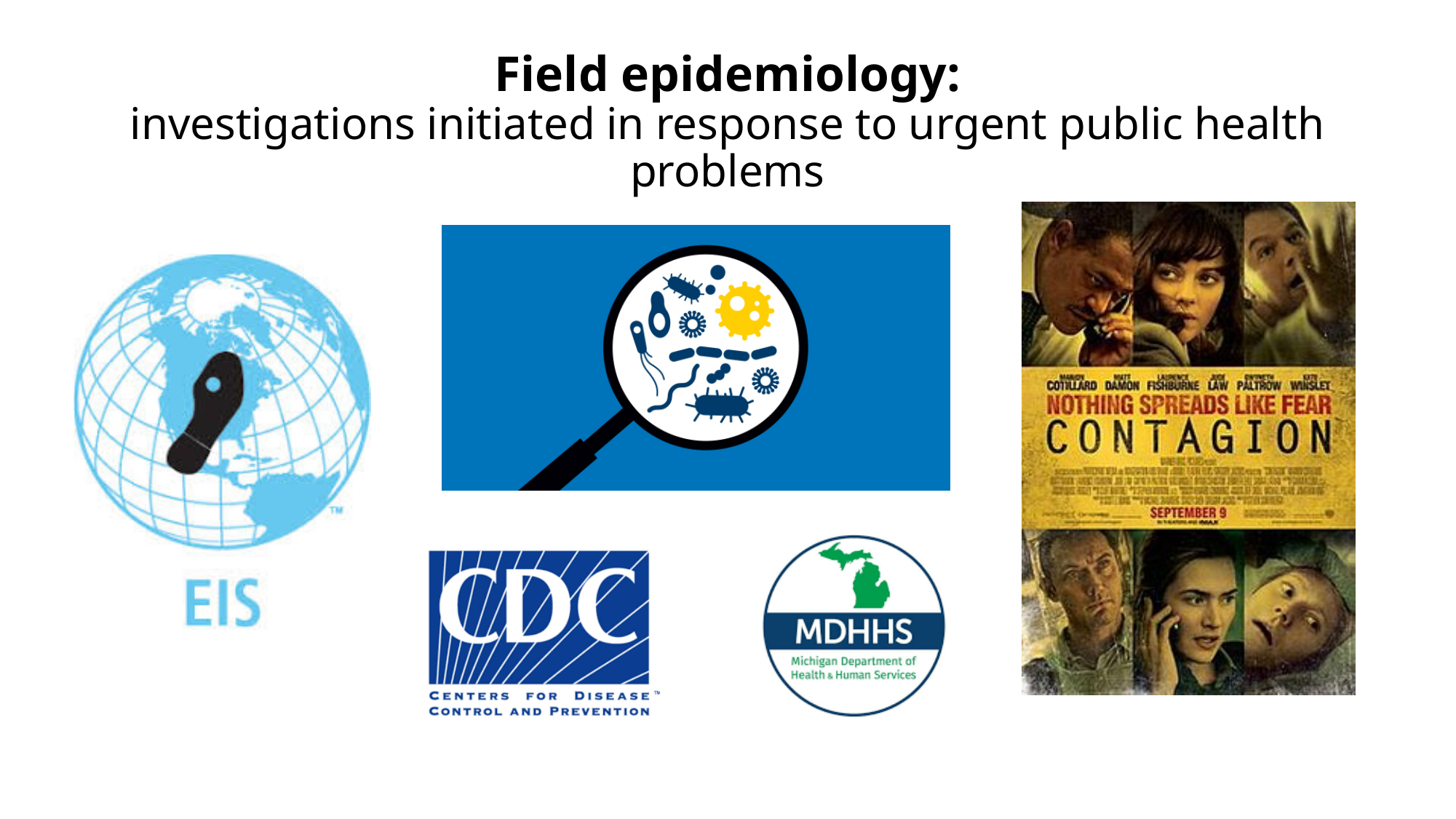

#### Field epidemiology: investigations initiated in response to urgent public health problems

### Slide 4
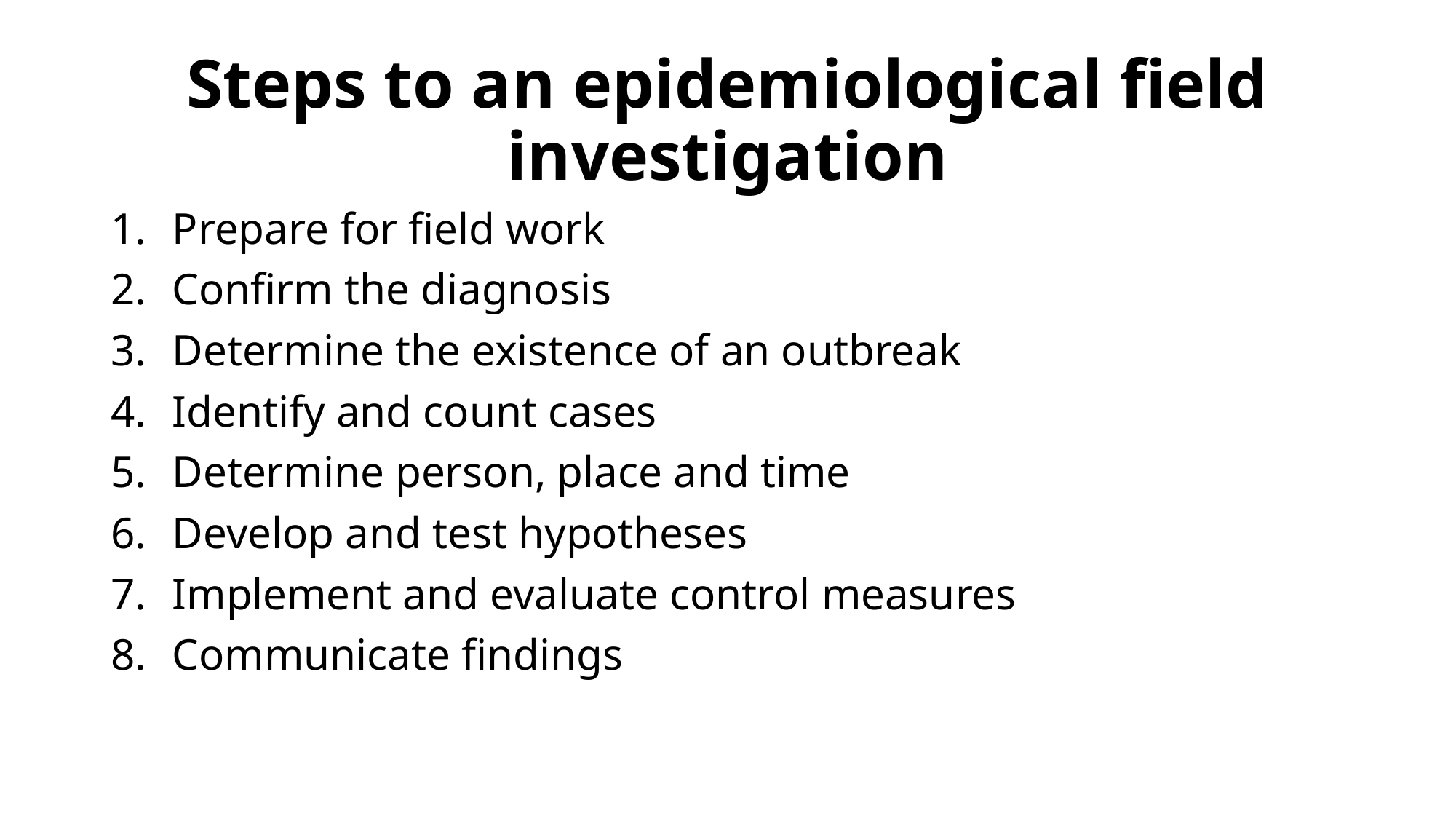

#### Steps to an epidemiological field investigation
Prepare for field work
Confirm the diagnosis
Determine the existence of an outbreak
Identify and count cases
Determine person, place and time
Develop and test hypotheses
Implement and evaluate control measures
Communicate findings

### Slide 5
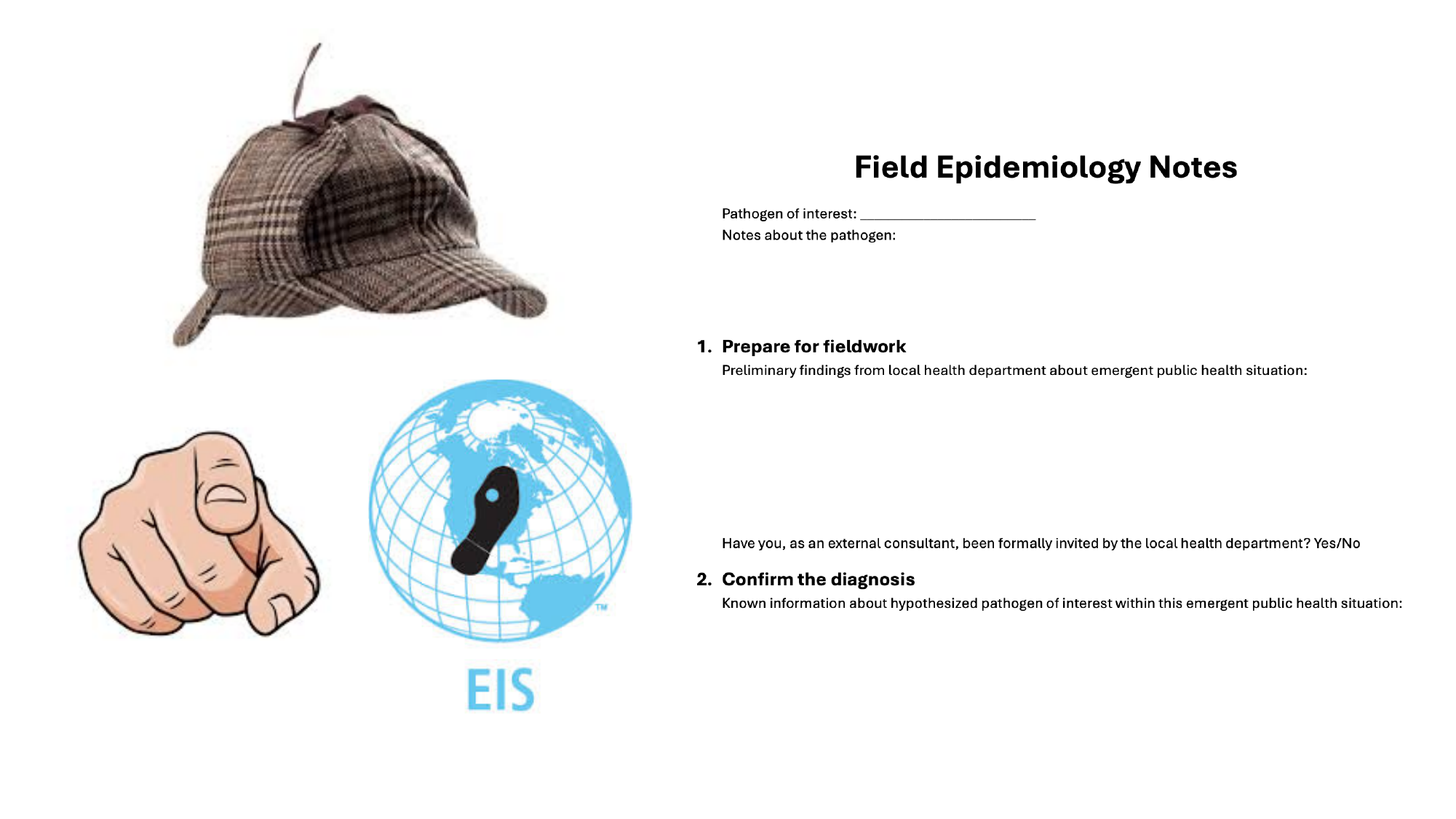

### Slide 6
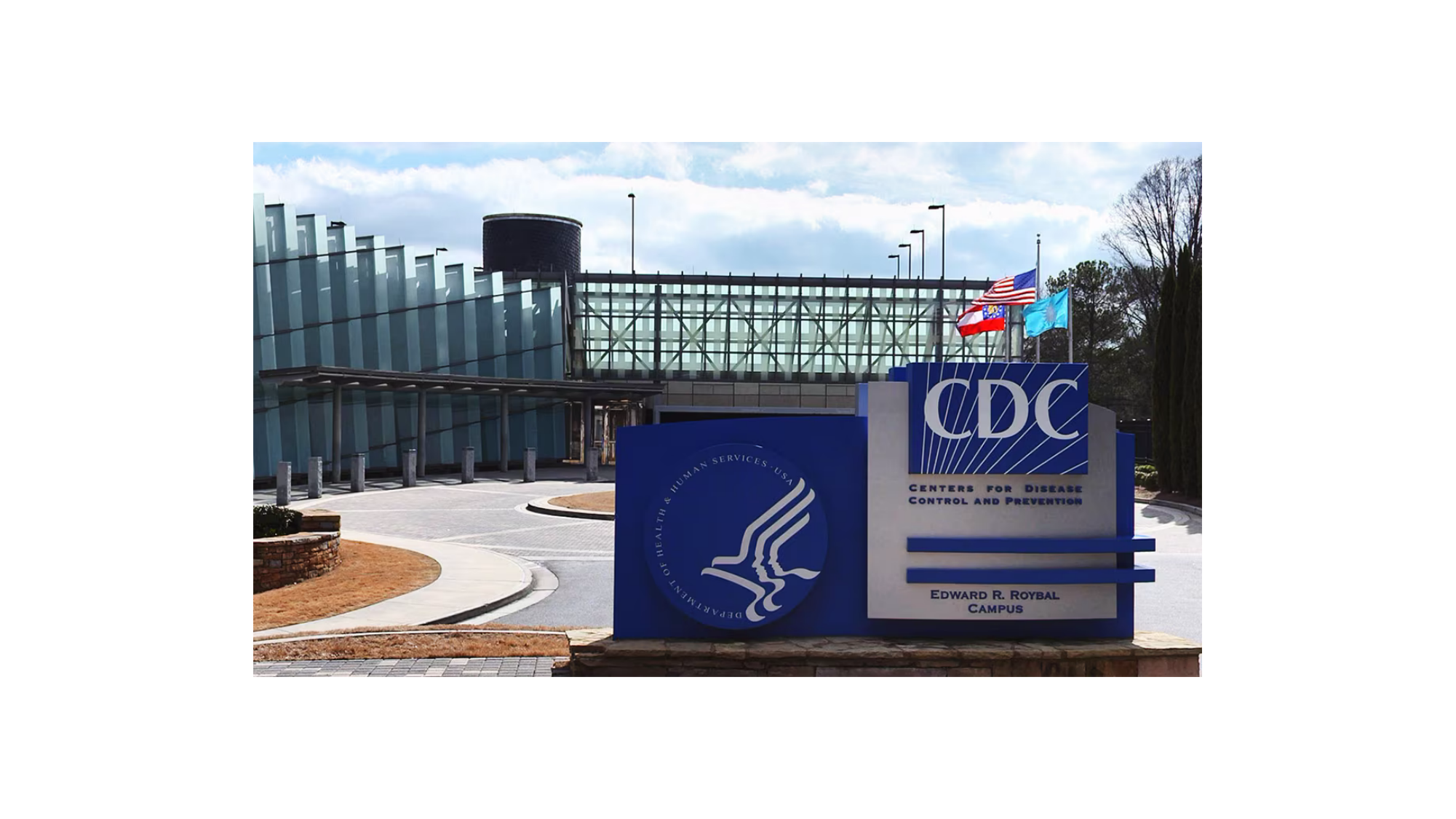

### Slide 7
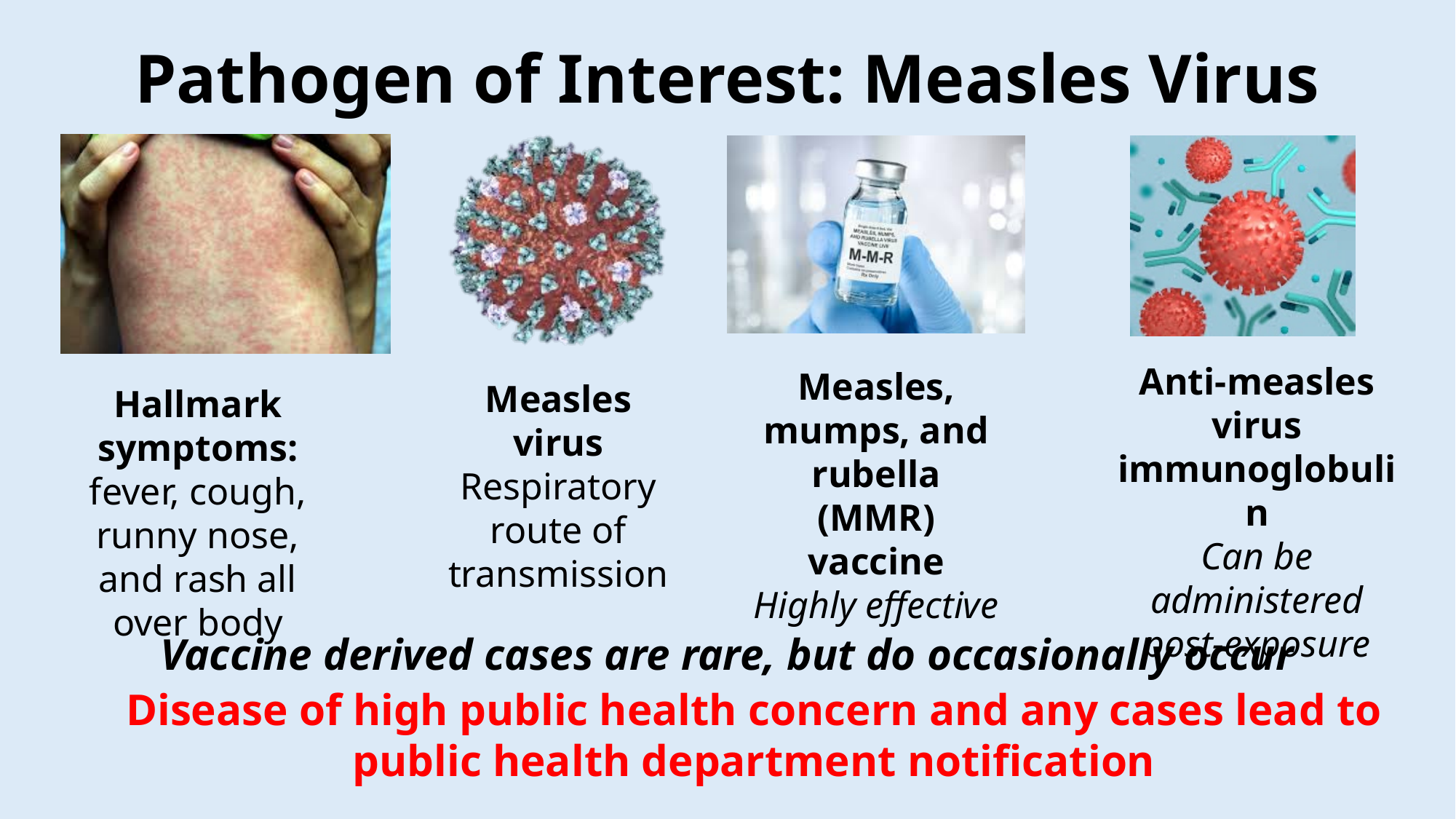

#### Pathogen of Interest: Measles Virus
Measles virus
Respiratory route of transmission
Anti-measles virus immunoglobulin
Can be administered post-exposure
Measles, mumps, and rubella (MMR) vaccine
Highly effective
Hallmark symptoms: fever, cough, runny nose, and rash all over body
Vaccine derived cases are rare, but do occasionally occur
Disease of high public health concern and any cases lead to public health department notification

### Slide 8
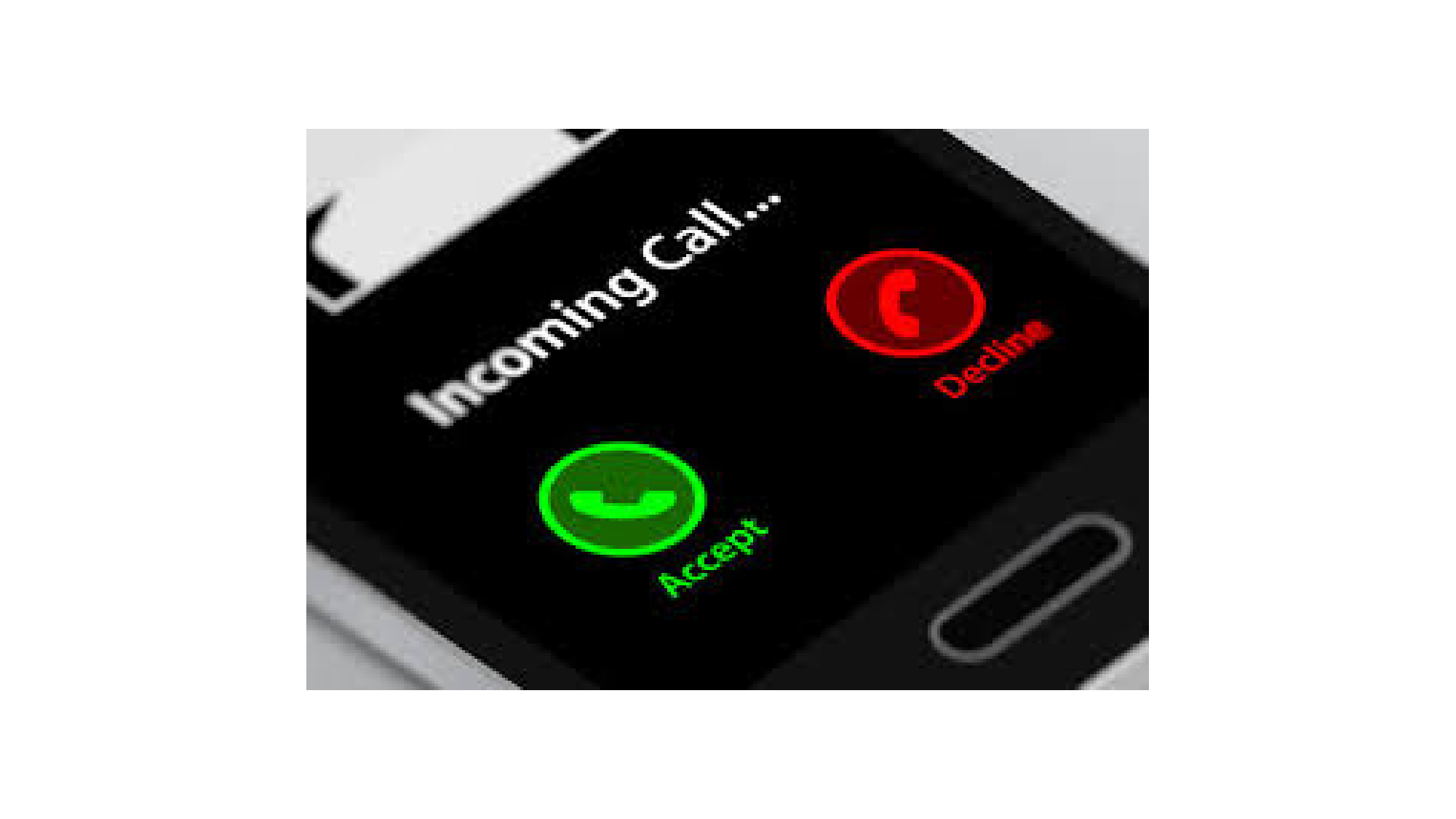

### Slide 9
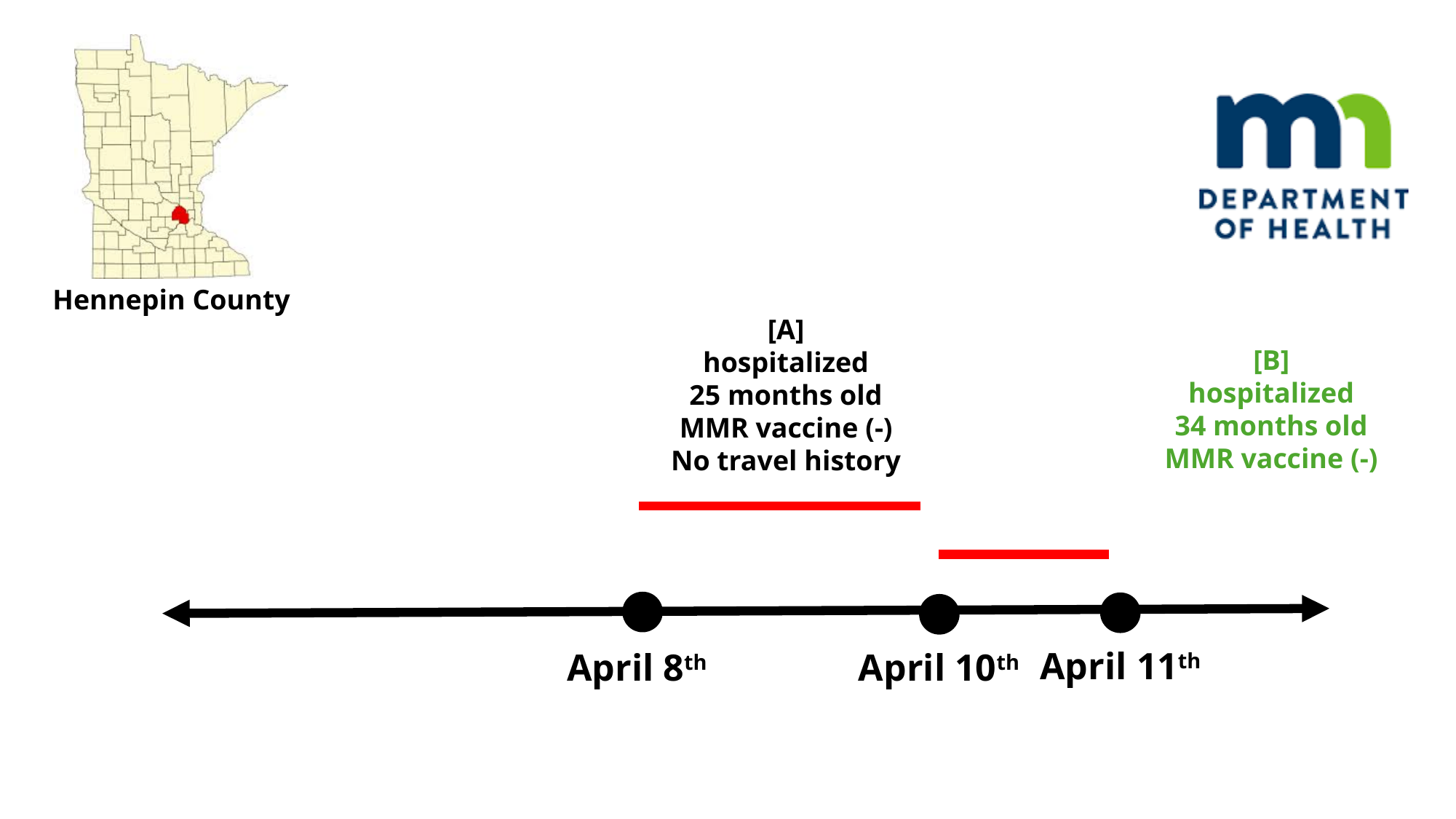

Hennepin County
[A]
hospitalized
25 months old
MMR vaccine (-)
No travel history
[B]
hospitalized
34 months old
MMR vaccine (-)
April 8^th^
April 11^th^
April 10^th^

### Slide 10
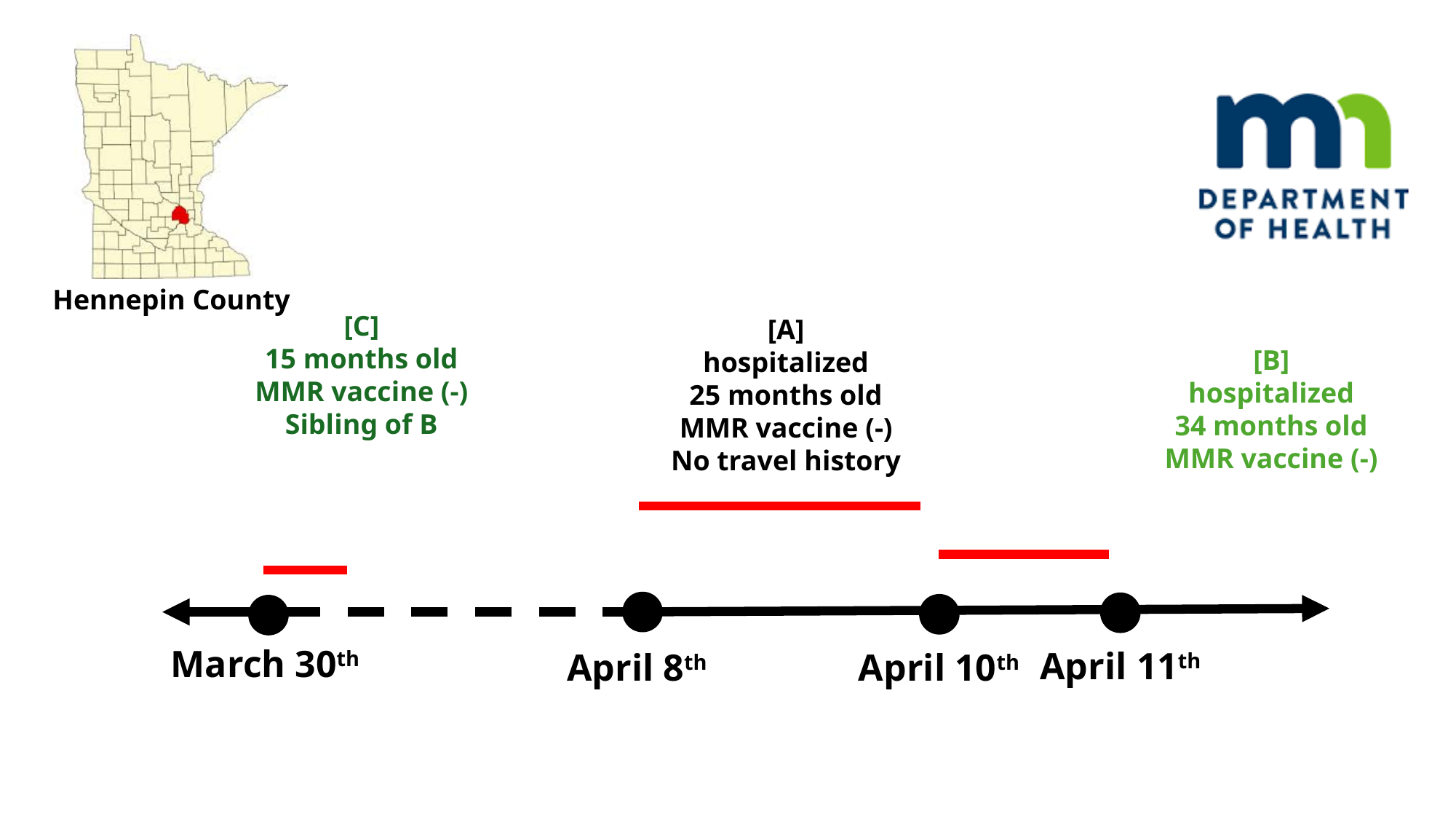

Hennepin County
[C]
15 months old
MMR vaccine (-)
Sibling of B
[A]
hospitalized
25 months old
MMR vaccine (-)
No travel history
[B]
hospitalized
34 months old
MMR vaccine (-)
April 8^th^
March 30^th^
April 11^th^
April 10^th^

### Slide 11
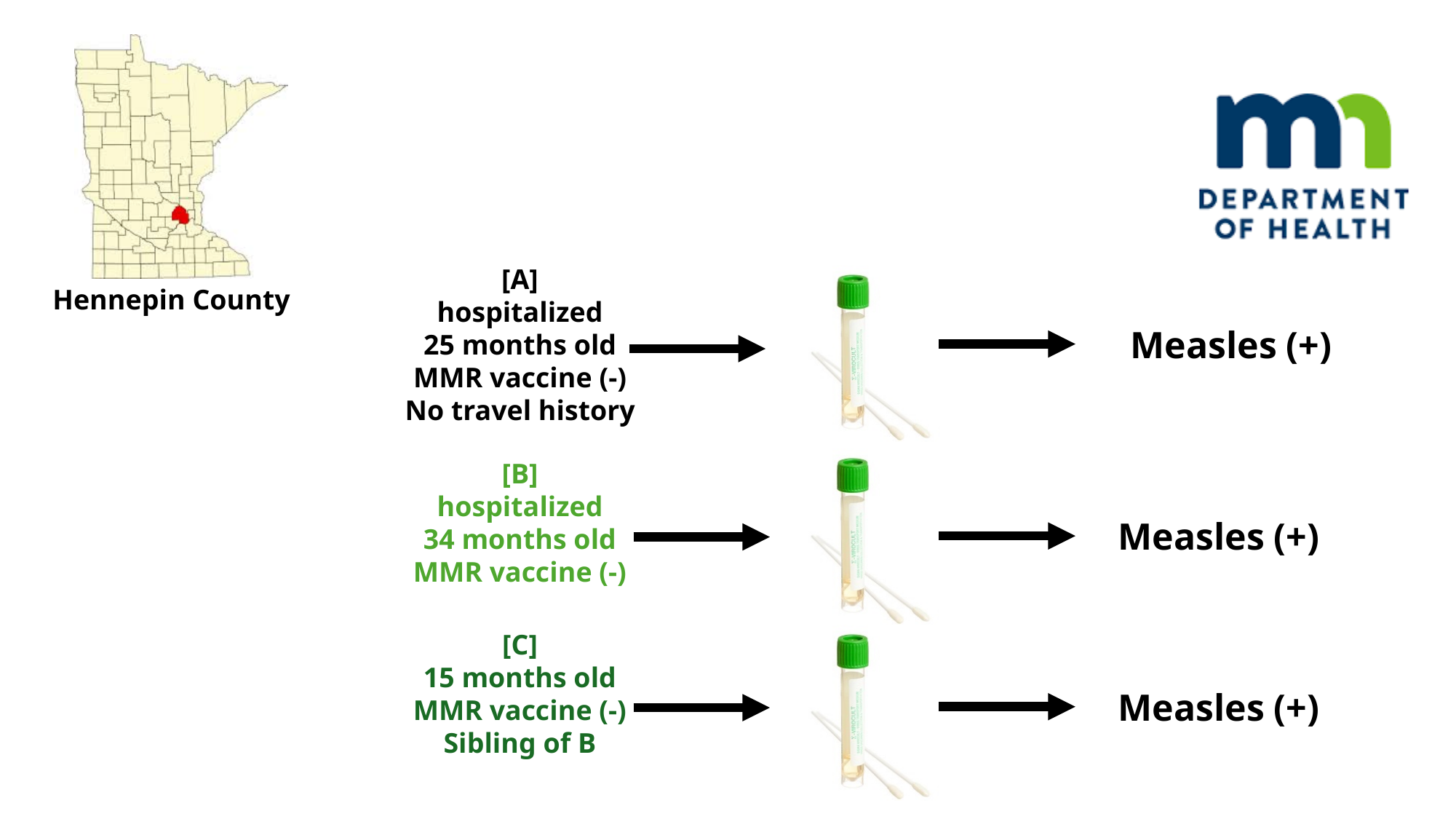

[A]
hospitalized
25 months old
MMR vaccine (-)
No travel history
Hennepin County
Measles (+)
Measles (+)
Measles (+)
[B]
hospitalized
34 months old
MMR vaccine (-)
[C]
15 months old
MMR vaccine (-)
Sibling of B

### Slide 12
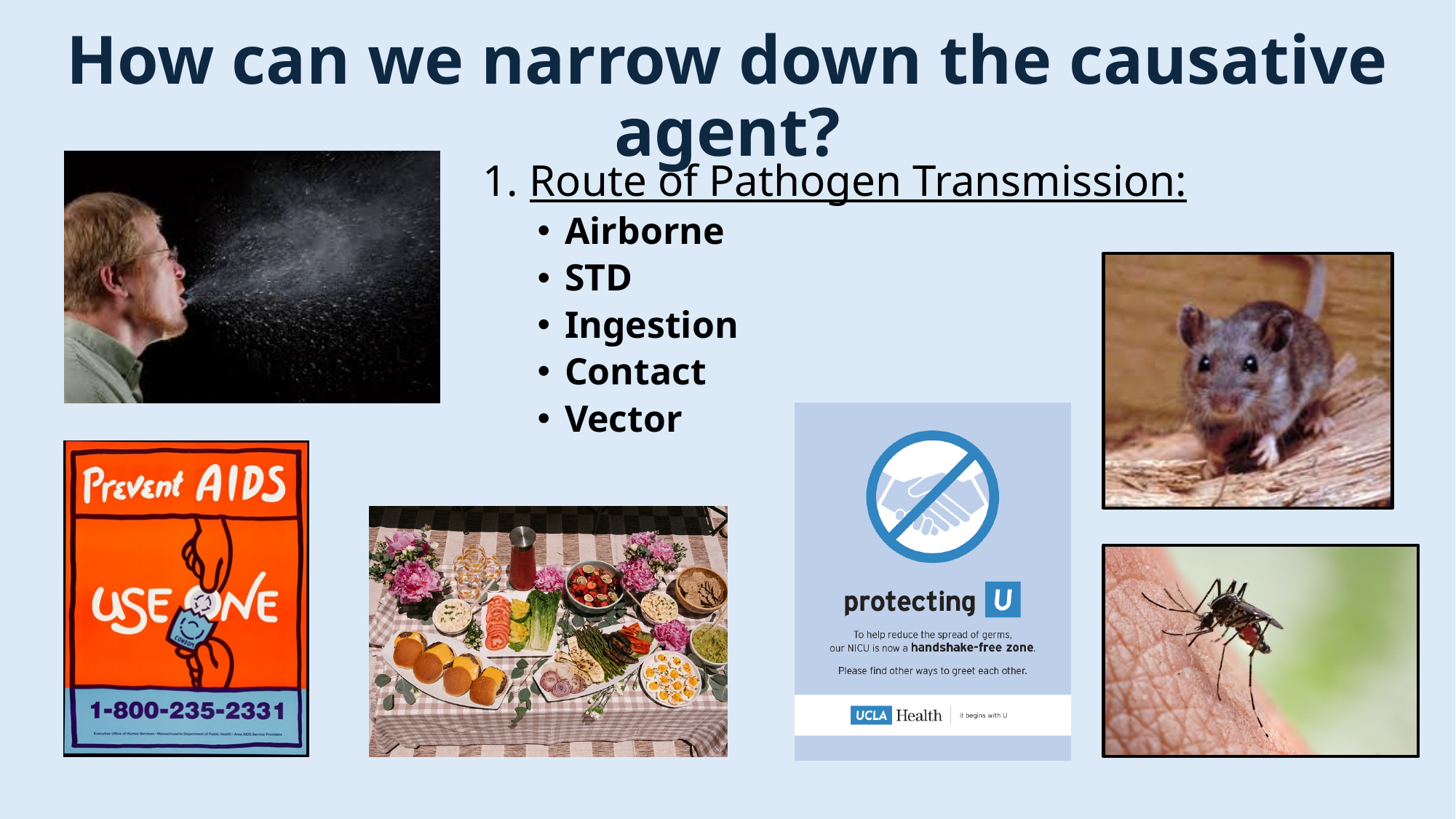

#### How can we narrow down the causative agent?
1. Route of Pathogen Transmission:
Airborne
STD
Ingestion
Contact
Vector

### Slide 13
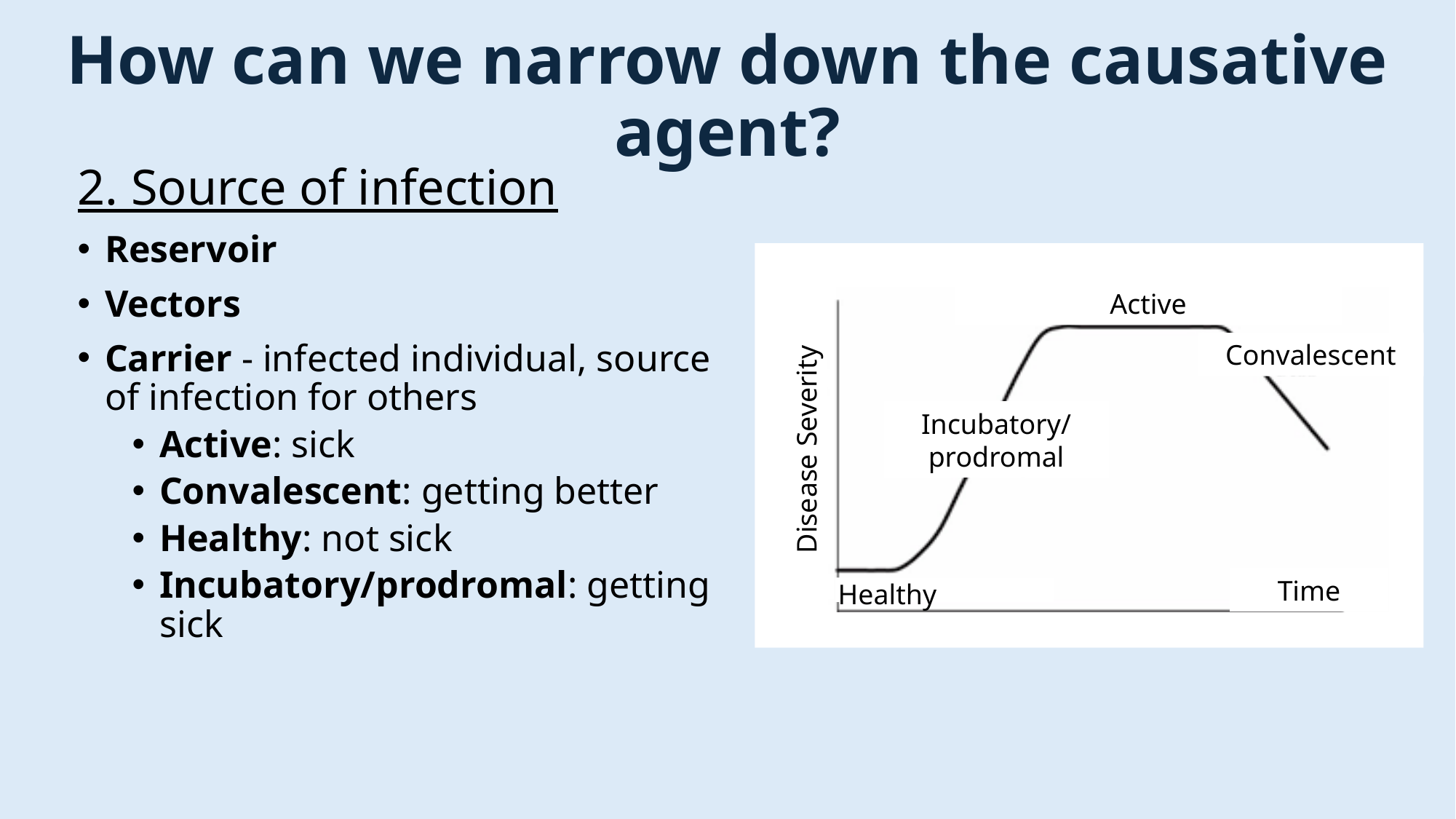

#### How can we narrow down the causative agent?
2. Source of infection
Reservoir
Vectors
Carrier - infected individual, source of infection for others
Active: sick
Convalescent: getting better
Healthy: not sick
Incubatory/prodromal: getting sick
Active
Convalescent
Incubatory/
prodromal
Disease Severity
Time
Healthy
***Note: Vectors and Carriers are the same thing

### Slide 14
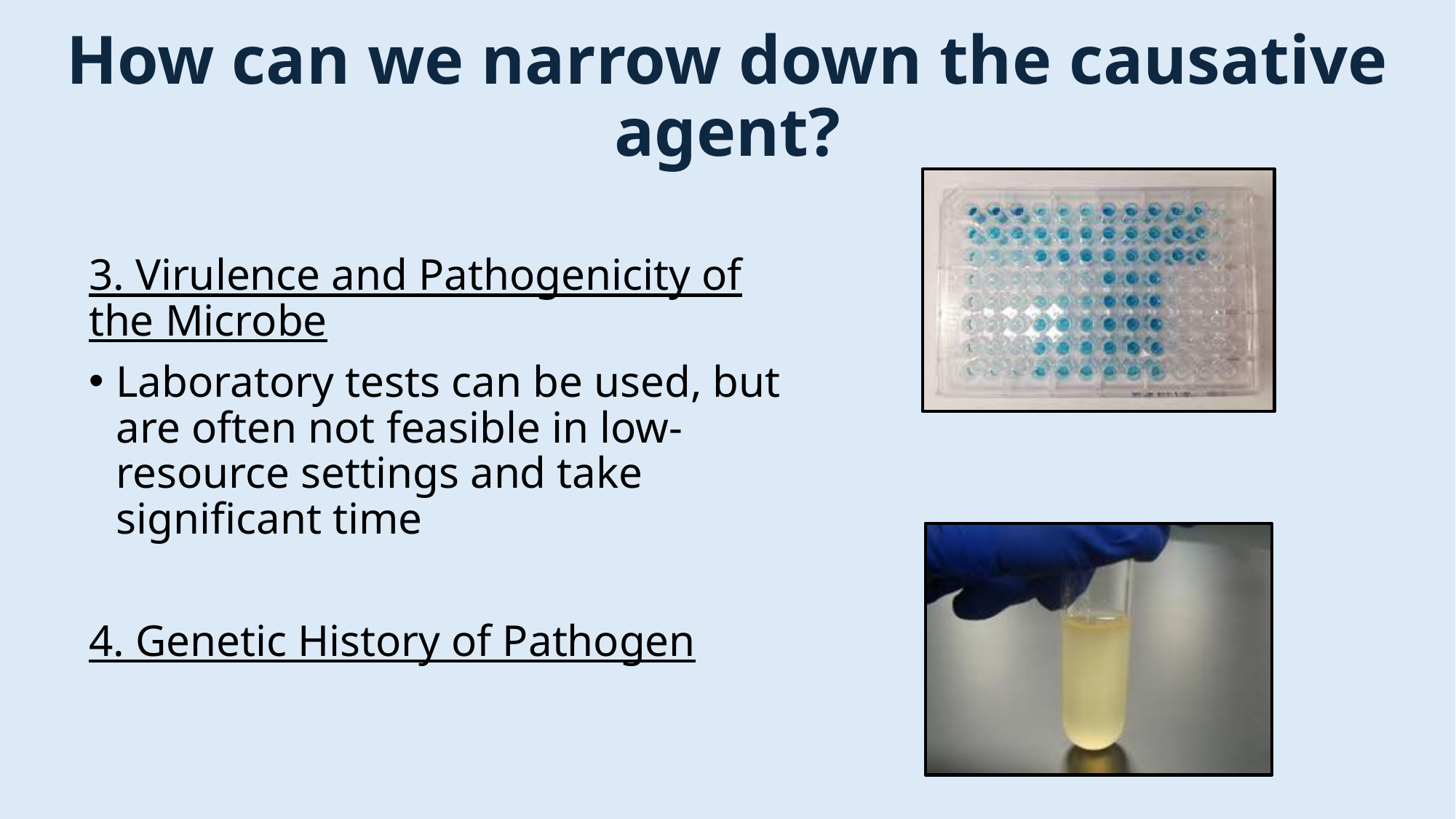

#### How can we narrow down the causative agent?
3. Virulence and Pathogenicity of the Microbe
Laboratory tests can be used, but are often not feasible in low-resource settings and take significant time
4. Genetic History of Pathogen

### Slide 15
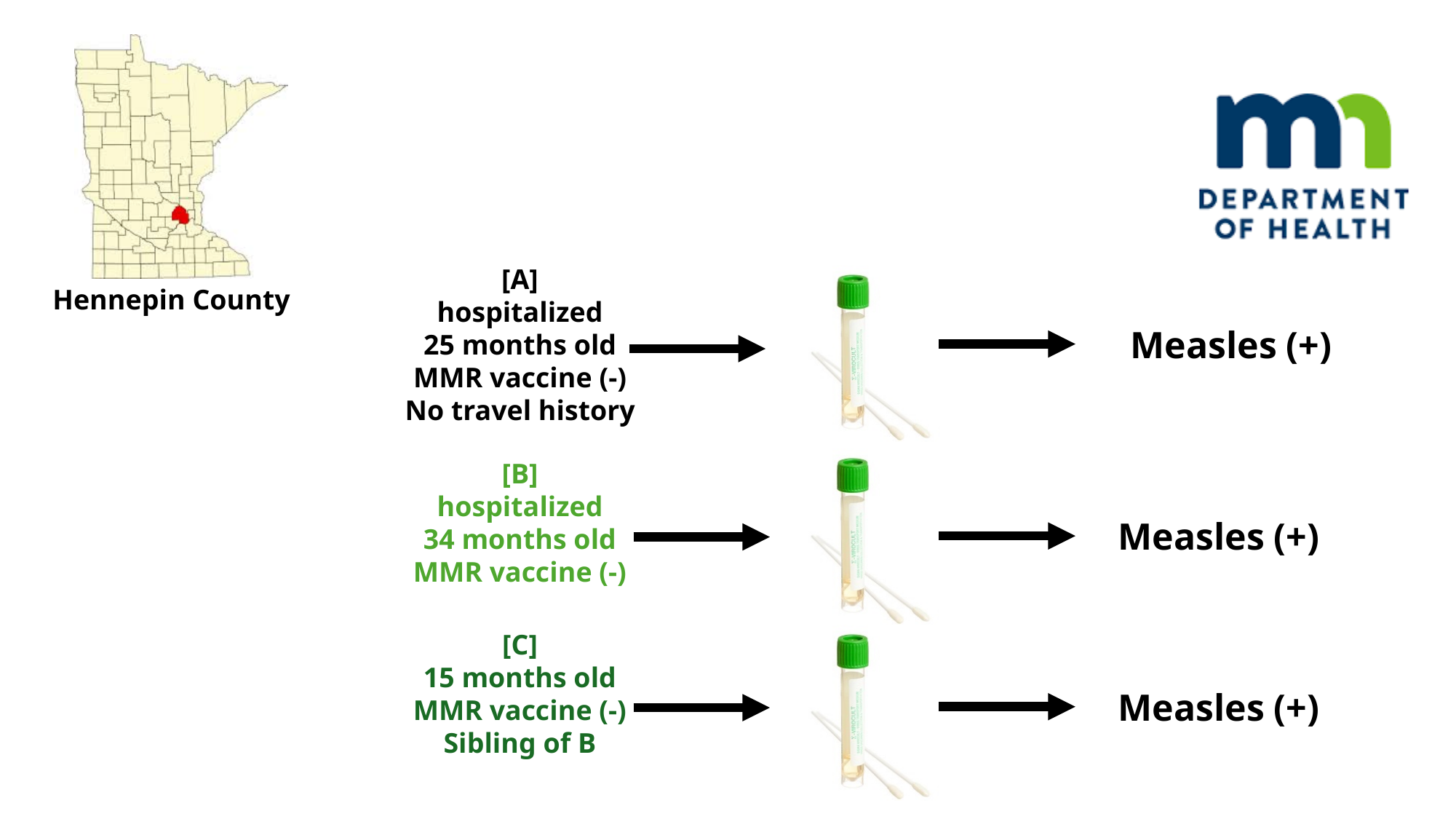

[A]
hospitalized
25 months old
MMR vaccine (-)
No travel history
Hennepin County
Measles (+)
Measles (+)
Measles (+)
[B]
hospitalized
34 months old
MMR vaccine (-)
[C]
15 months old
MMR vaccine (-)
Sibling of B

### Slide 16
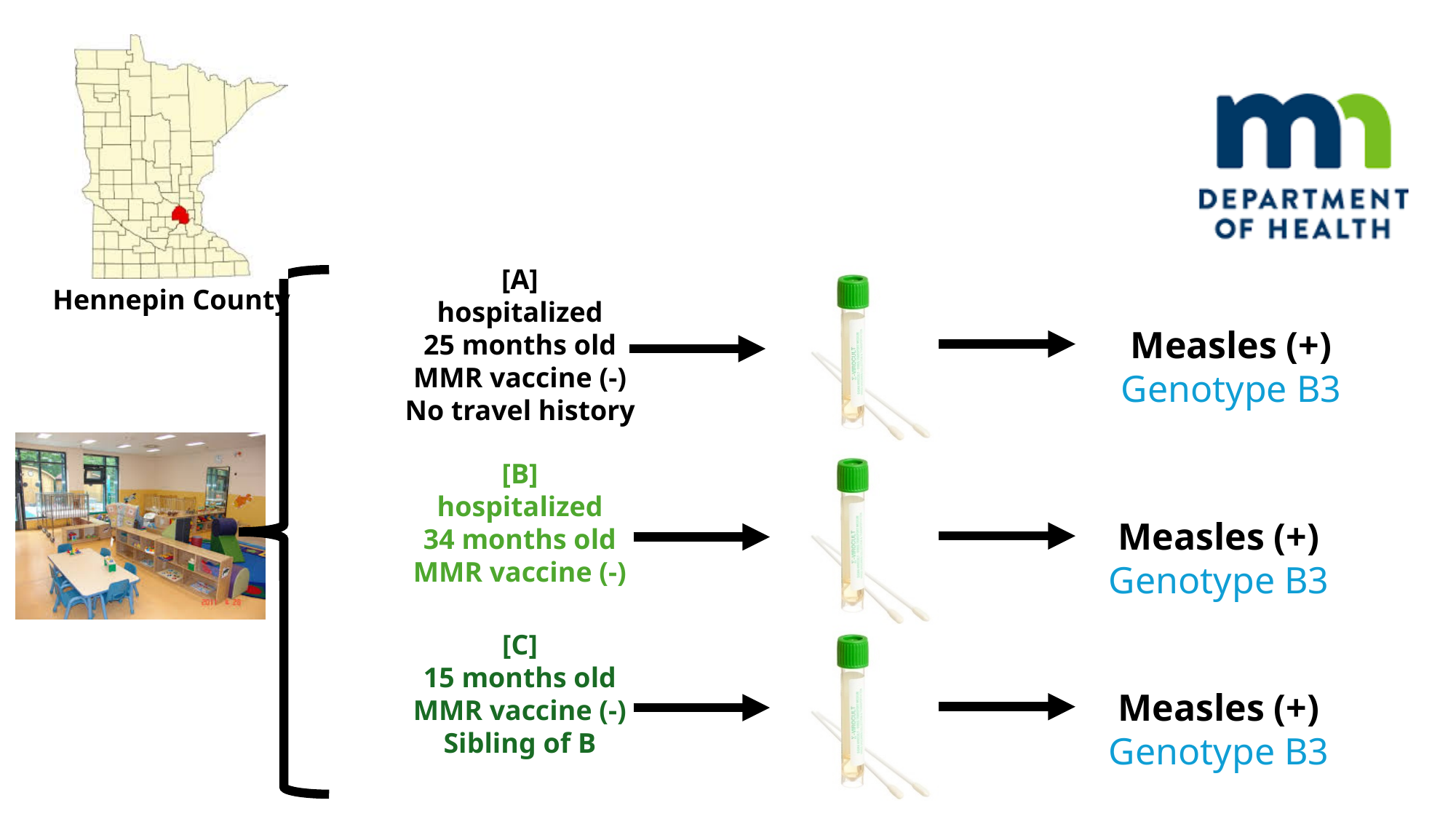

[A]
hospitalized
25 months old
MMR vaccine (-)
No travel history
Hennepin County
Measles (+)
Genotype B3
Measles (+)
Genotype B3
Measles (+)
Genotype B3
[B]
hospitalized
34 months old
MMR vaccine (-)
[C]
15 months old
MMR vaccine (-)
Sibling of B

### Slide 17
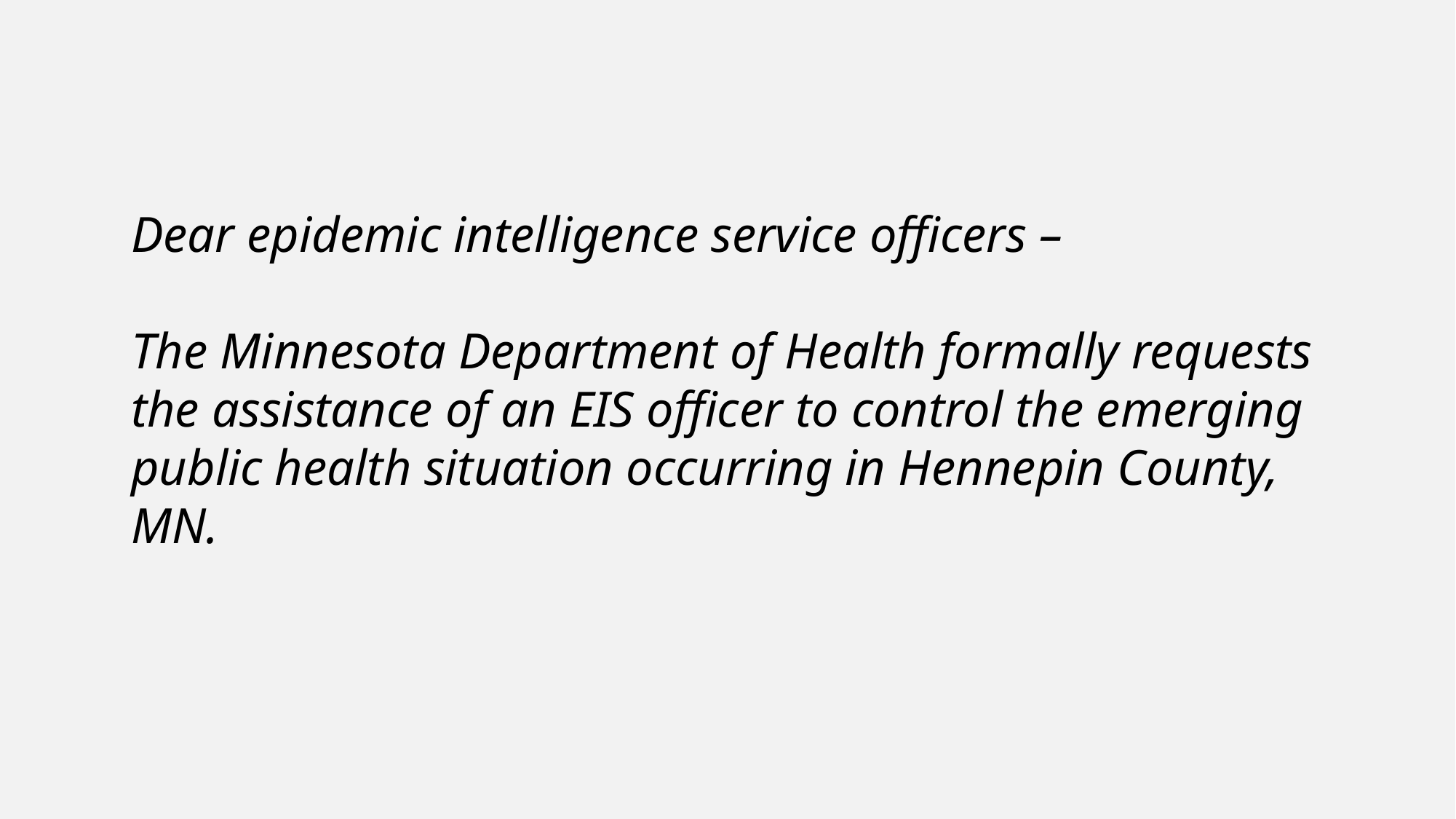

Dear epidemic intelligence service officers –
The Minnesota Department of Health formally requests the assistance of an EIS officer to control the emerging public health situation occurring in Hennepin County, MN.

### Slide 18
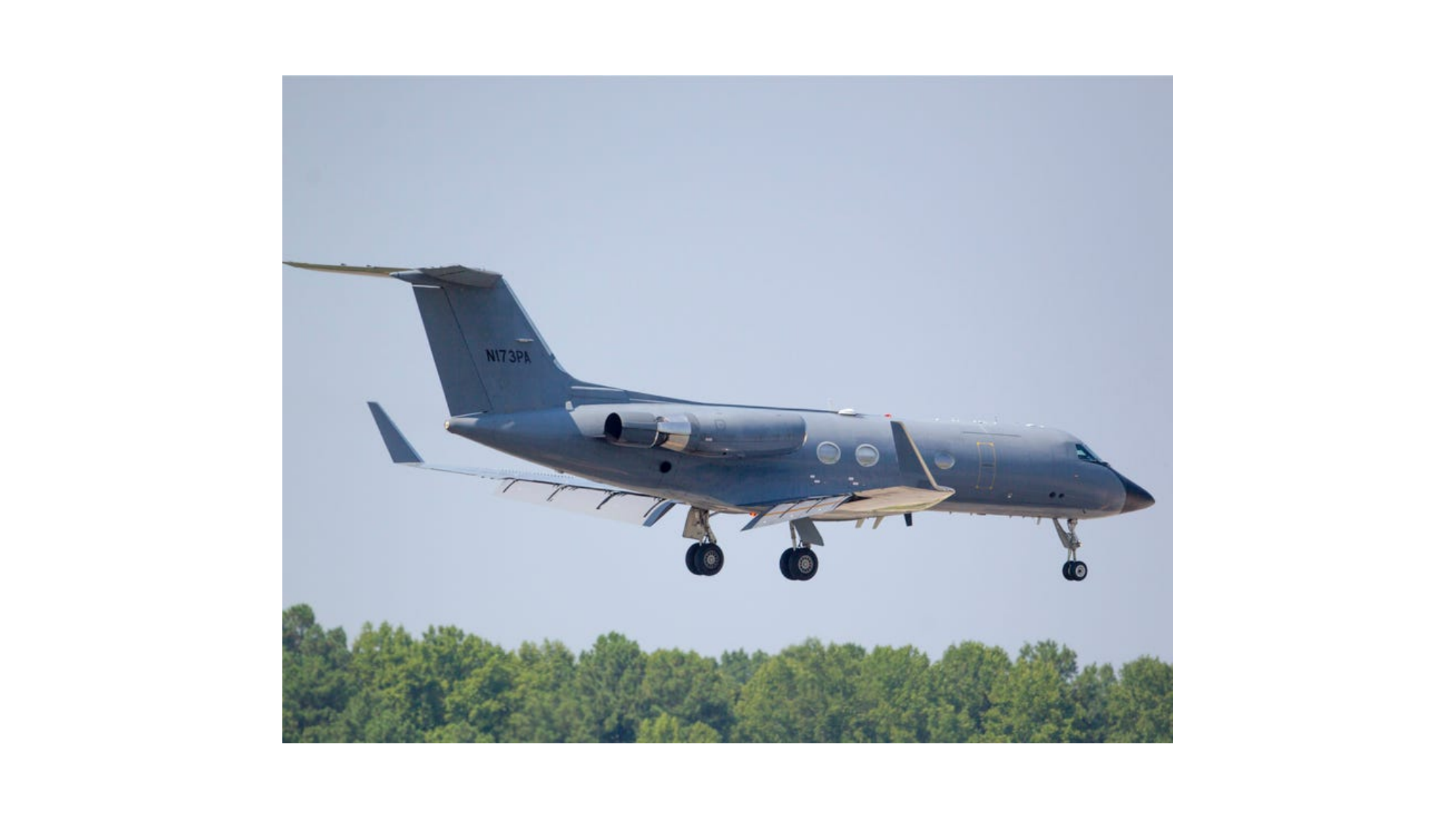

### Slide 19
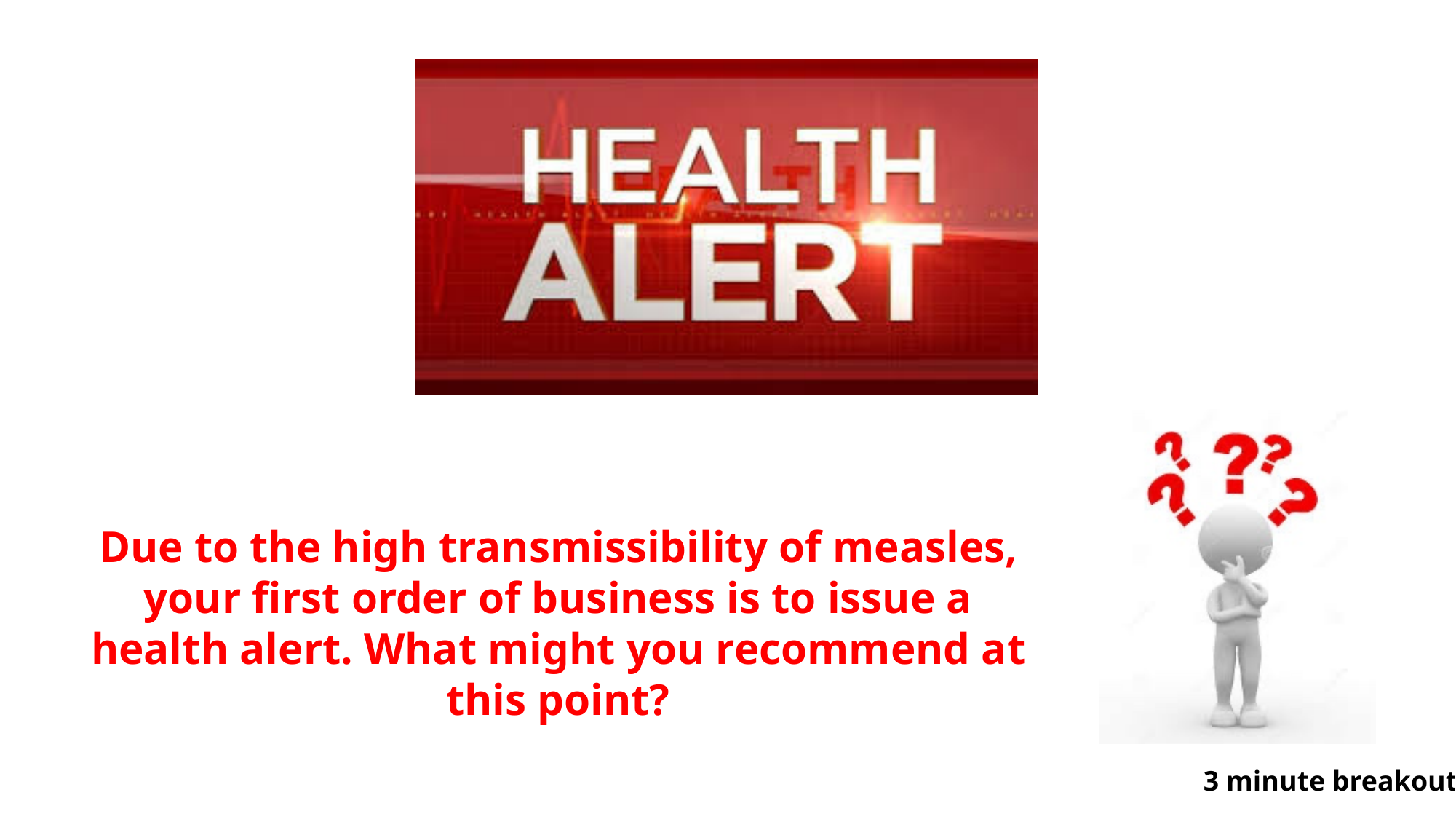

Due to the high transmissibility of measles, your first order of business is to issue a health alert. What might you recommend at this point?
3 minute breakout

### Slide 20
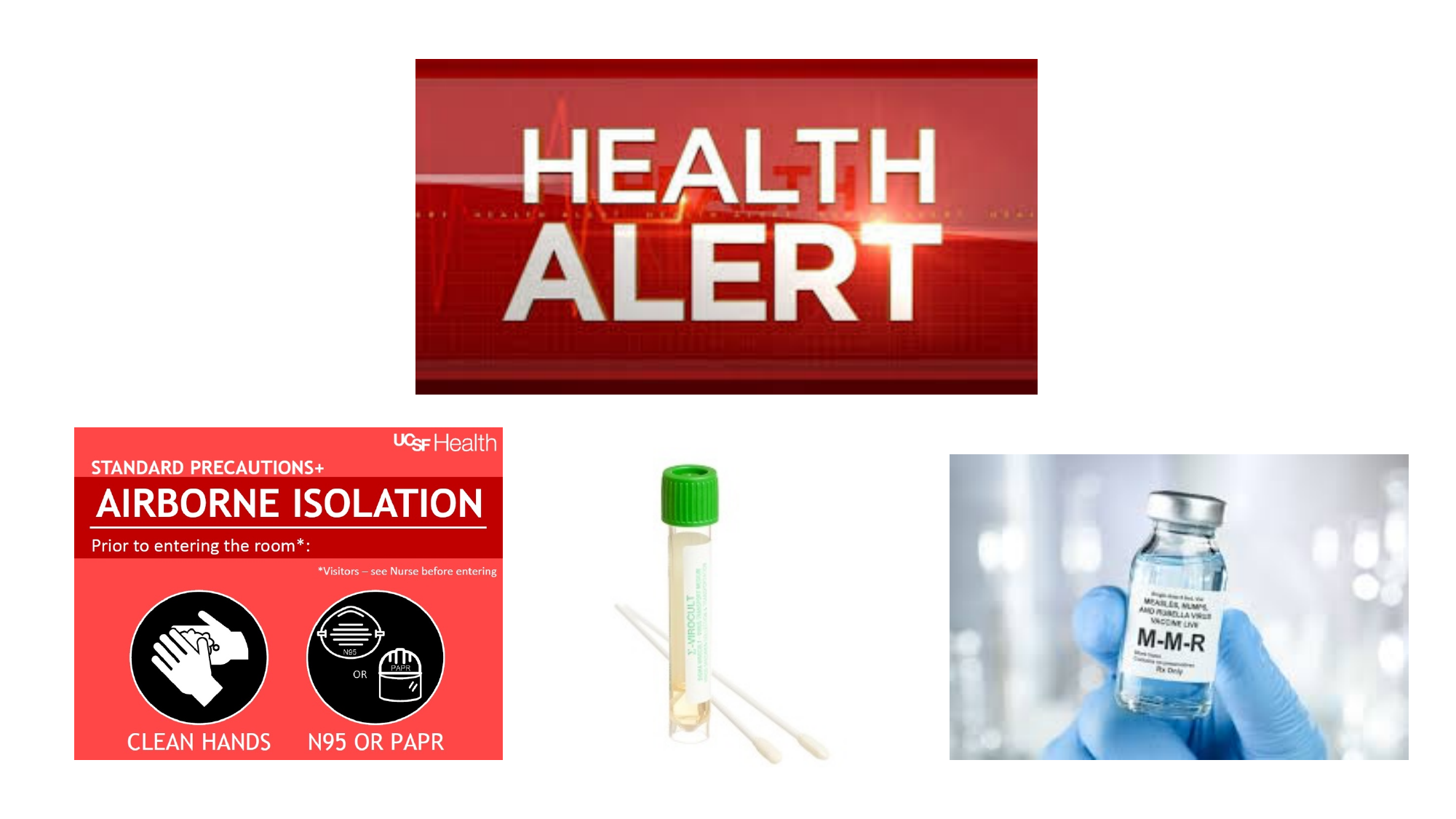

### Slide 21
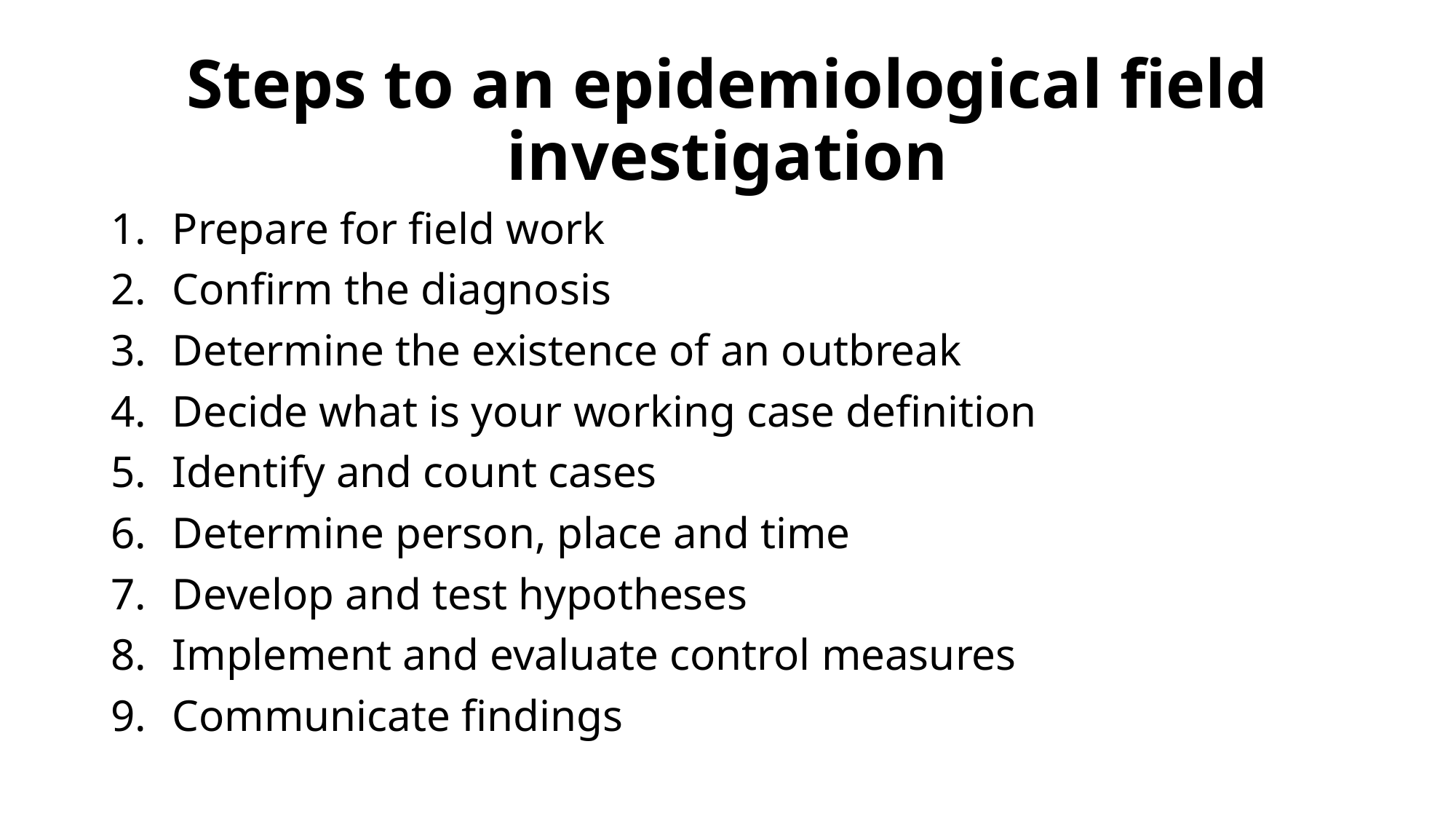

#### Steps to an epidemiological field investigation
Prepare for field work
Confirm the diagnosis
Determine the existence of an outbreak
Decide what is your working case definition
Identify and count cases
Determine person, place and time
Develop and test hypotheses
Implement and evaluate control measures
Communicate findings

### Slide 22
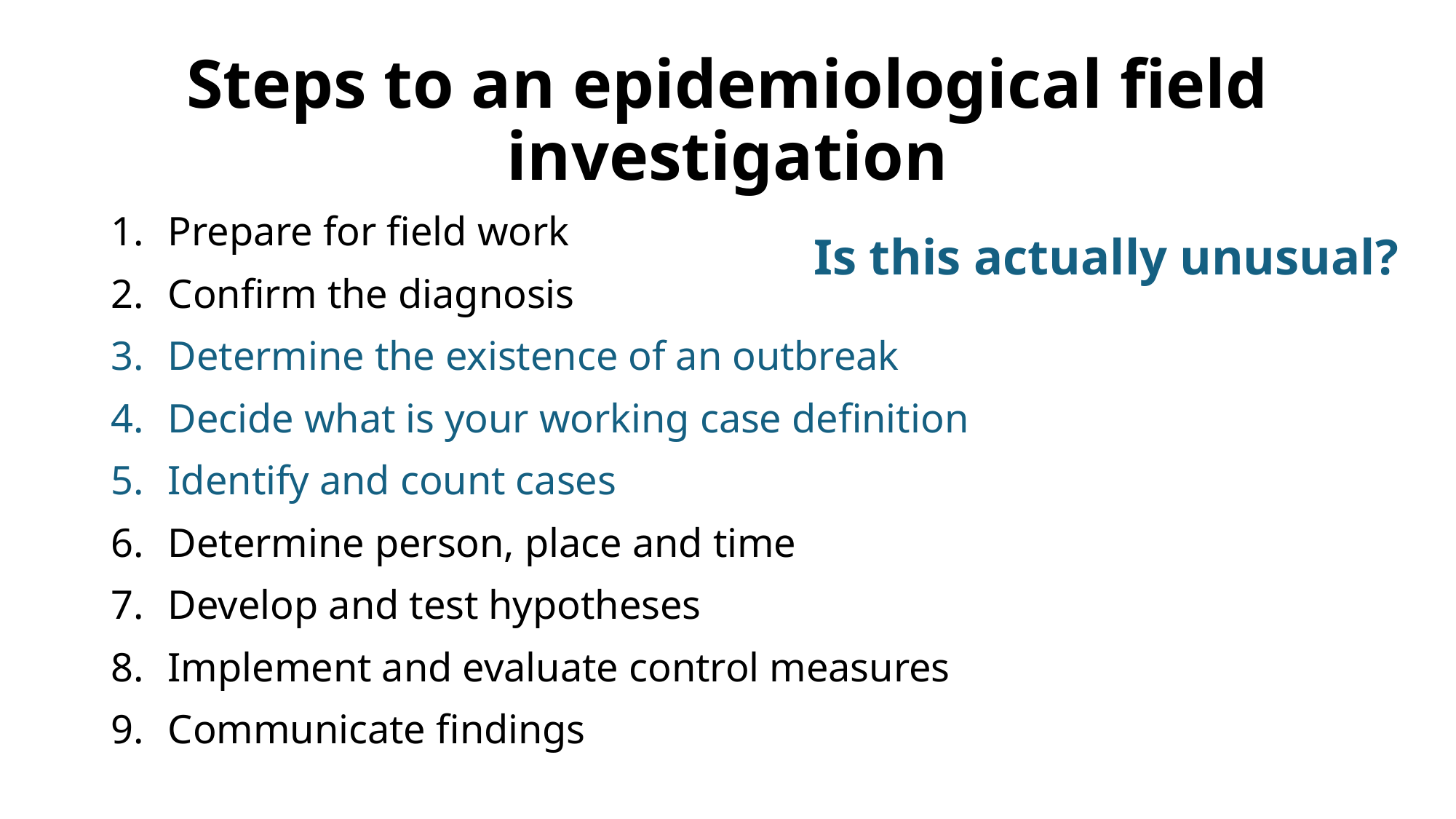

#### Steps to an epidemiological field investigation
Prepare for field work
Confirm the diagnosis
Determine the existence of an outbreak
Decide what is your working case definition
Identify and count cases
Determine person, place and time
Develop and test hypotheses
Implement and evaluate control measures
Communicate findings
Is this actually unusual?

### Slide 23
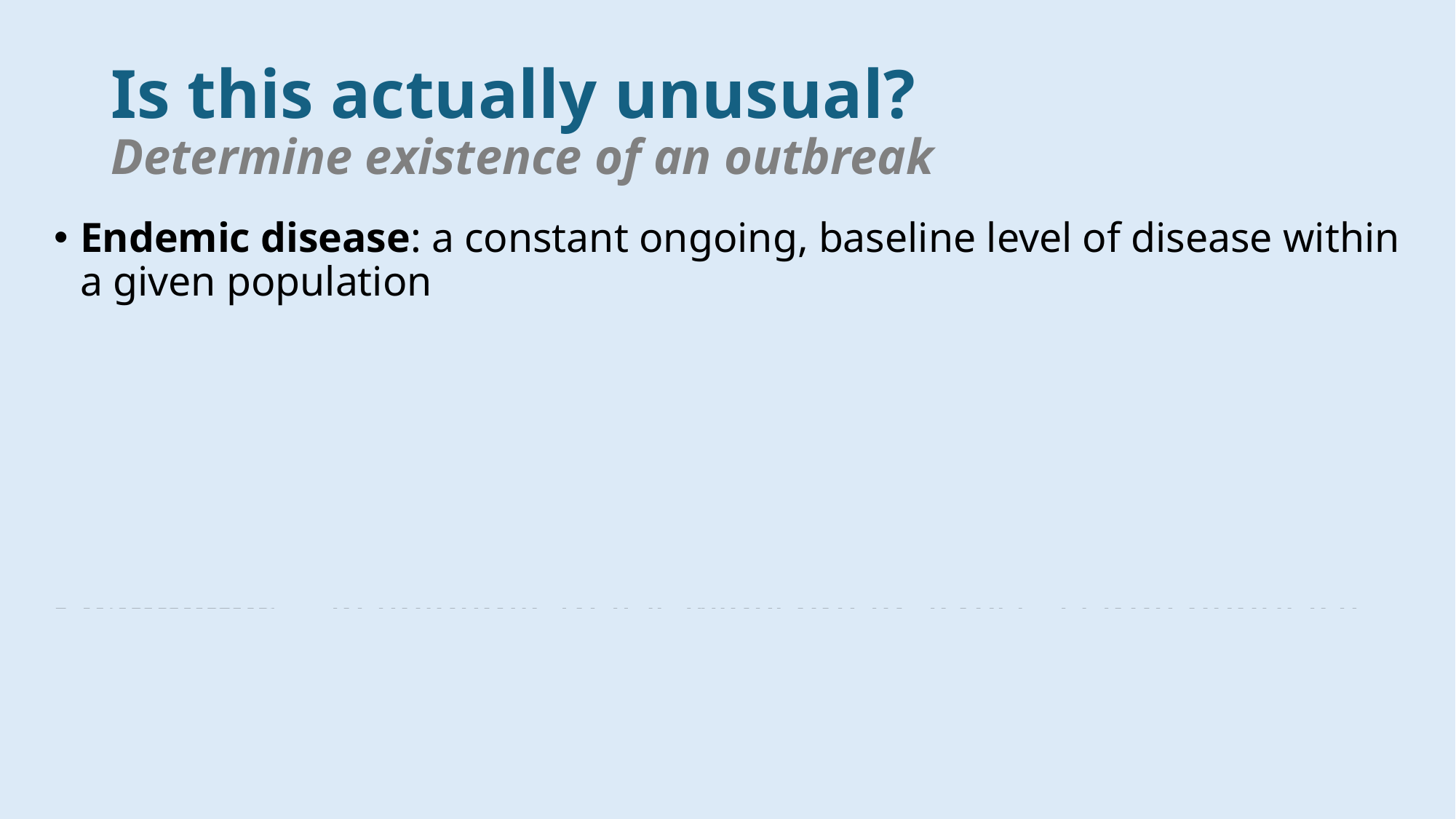

#### Is this actually unusual? Determine existence of an outbreak
Endemic disease: a constant ongoing, baseline level of disease within a given population
Outbreak/Epidemic – occurrence of disease at a higher-than-baseline level in a given population
Can be a single case in a new population
“Outbreak” tends to be used more frequently by the media and “epidemic” tends to be used more in academic settings
Examples: Yellow Fever (1793), Typhoid Fever (1906-1907), Ebola (2014)
Pandemic – an epidemic that is widespread across a large populace and across multiple geographic regions/locations (i.e., worldwide)
Example: HIV (1981 – present), H1N1 flu (2009-2010), COVID-19 (2020 – present)

### Slide 24
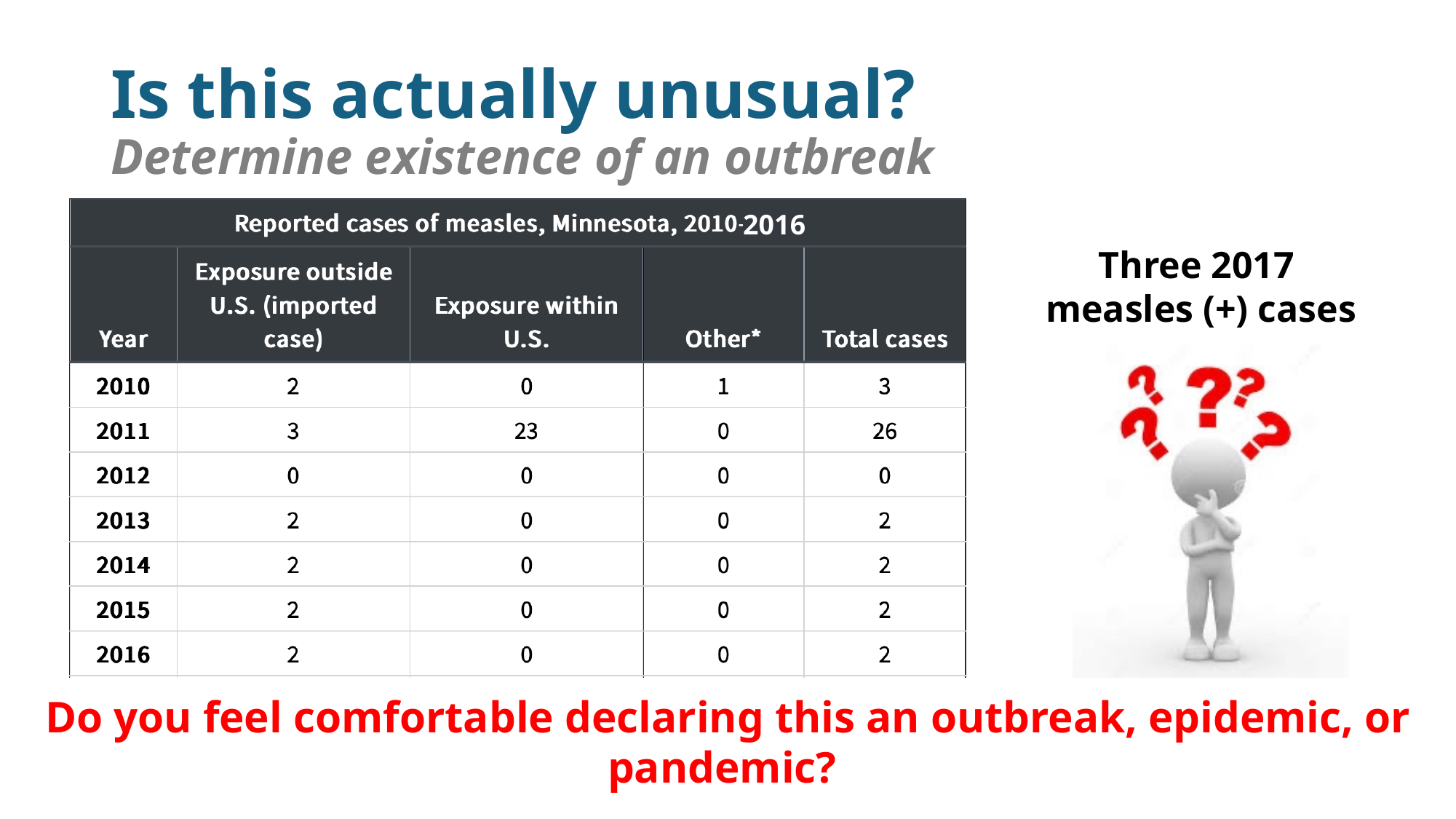

Three 2017
measles (+) cases
#### Is this actually unusual? Determine existence of an outbreak
2016
Do you feel comfortable declaring this an outbreak, epidemic, or pandemic?

### Slide 25
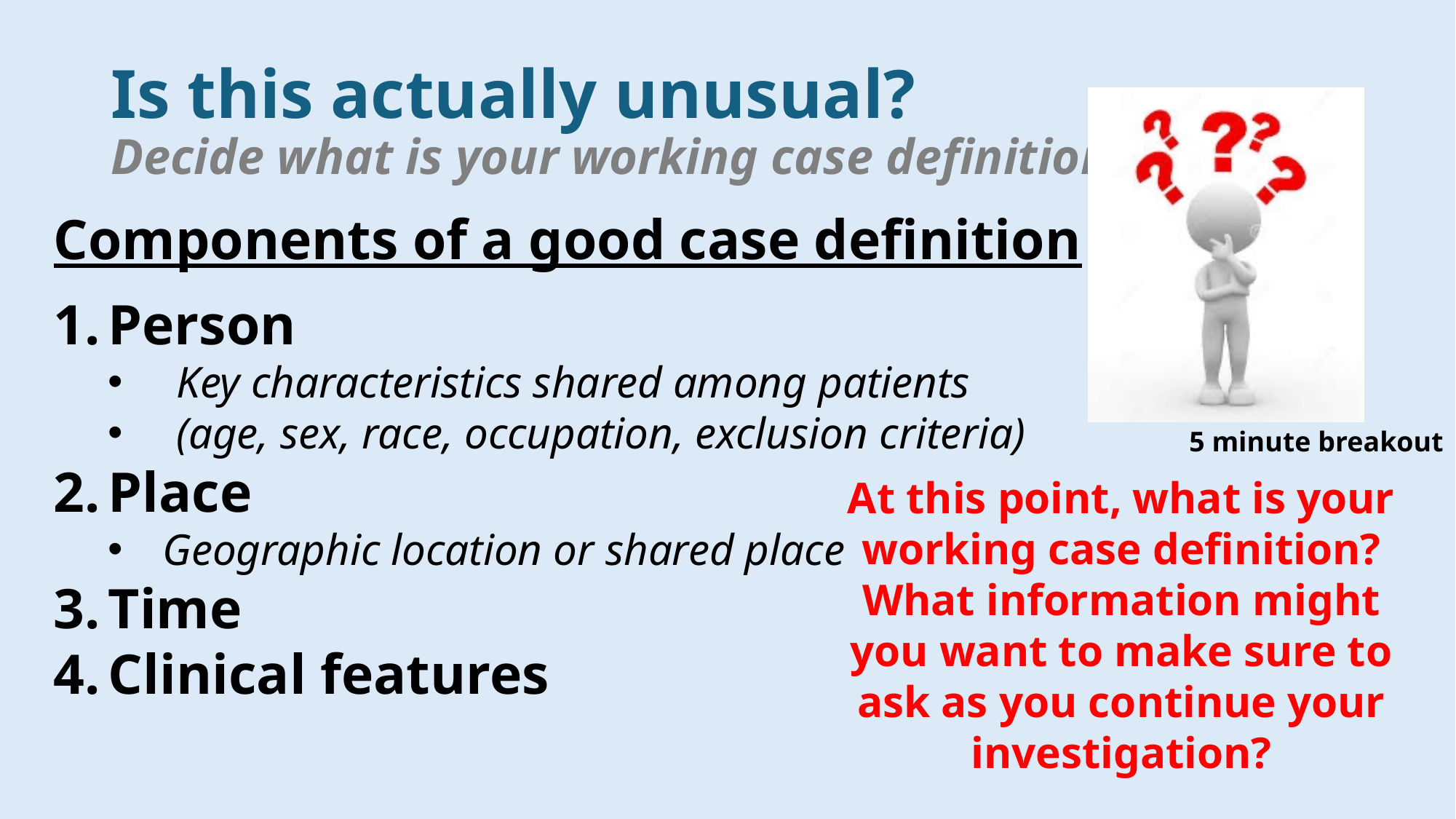

#### Is this actually unusual? Decide what is your working case definition
At this point, what is your working case definition? What information might you want to make sure to ask as you continue your investigation?
5 minute breakout
Components of a good case definition
Person
Key characteristics shared among patients
(age, sex, race, occupation, exclusion criteria)
Place
Geographic location or shared place
Time
Clinical features

### Slide 26
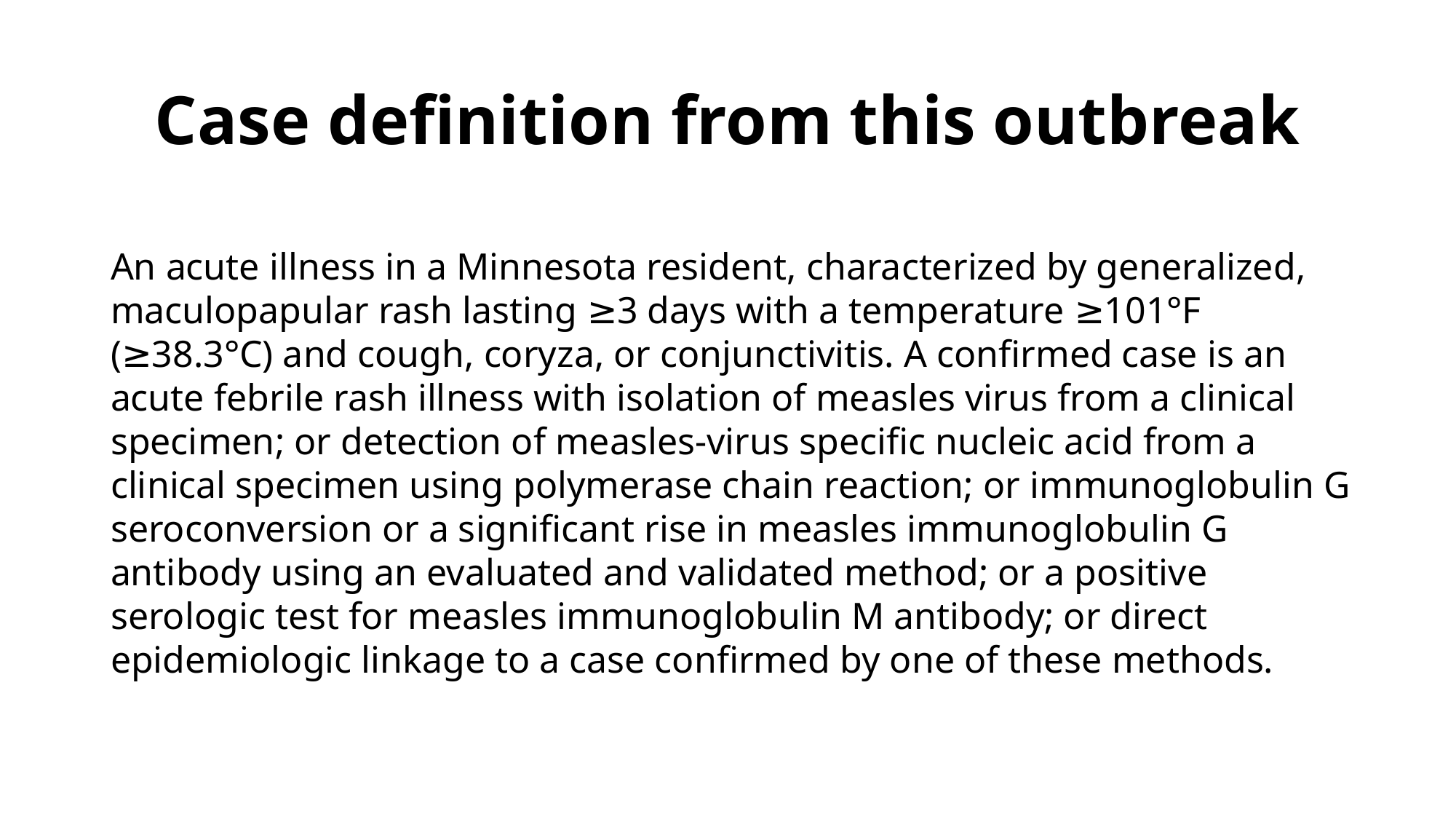

#### Case definition from this outbreak
An acute illness in a Minnesota resident, characterized by generalized, maculopapular rash lasting ≥3 days with a temperature ≥101°F (≥38.3°C) and cough, coryza, or conjunctivitis. A confirmed case is an acute febrile rash illness with isolation of measles virus from a clinical specimen; or detection of measles-virus specific nucleic acid from a clinical specimen using polymerase chain reaction; or immunoglobulin G seroconversion or a significant rise in measles immunoglobulin G antibody using an evaluated and validated method; or a positive serologic test for measles immunoglobulin M antibody; or direct epidemiologic linkage to a case confirmed by one of these methods.

### Slide 27
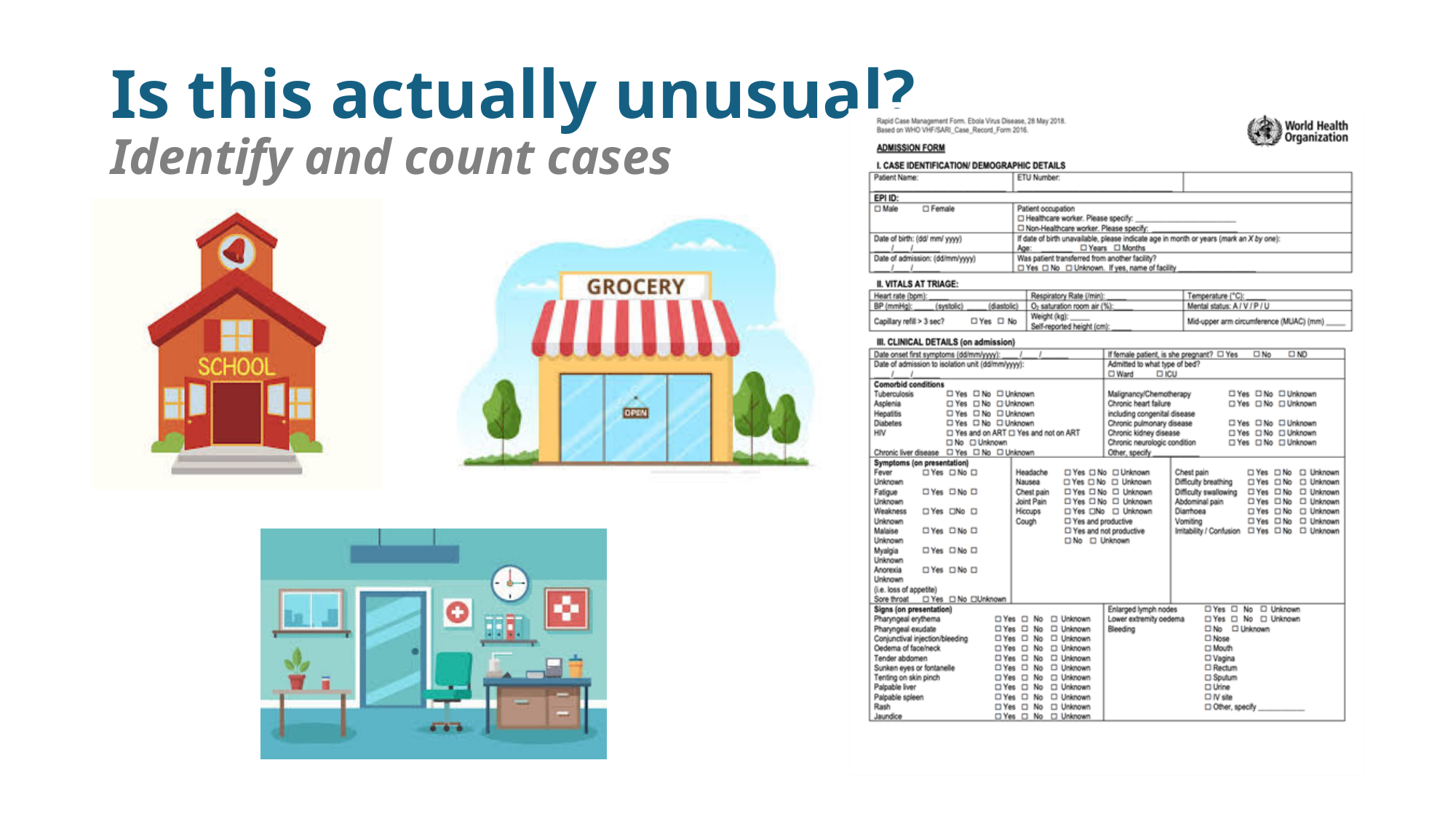

#### Is this actually unusual? Identify and count cases

### Slide 28
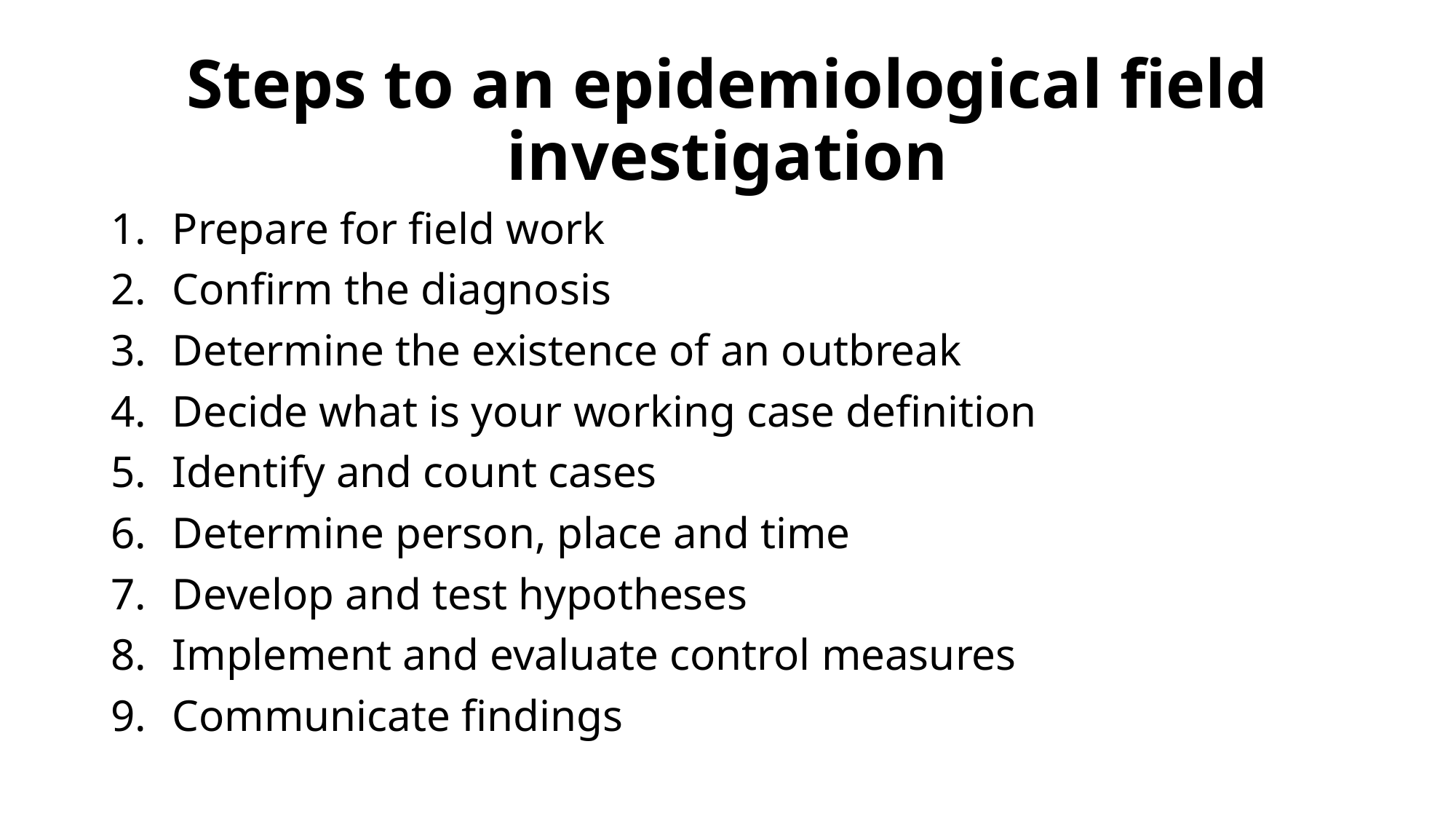

#### Steps to an epidemiological field investigation
Prepare for field work
Confirm the diagnosis
Determine the existence of an outbreak
Decide what is your working case definition
Identify and count cases
Determine person, place and time
Develop and test hypotheses
Implement and evaluate control measures
Communicate findings

### Slide 29
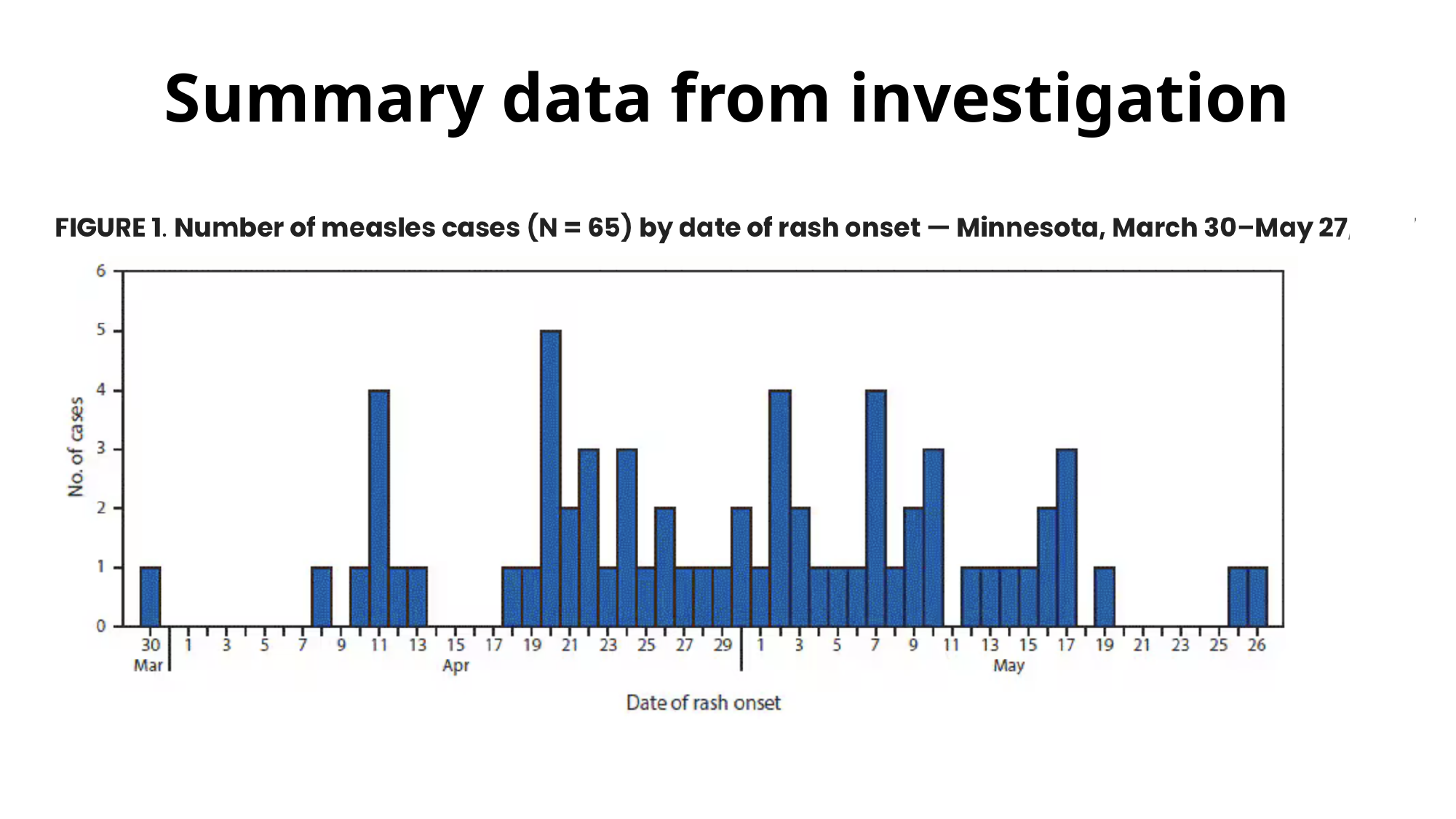

#### Summary data from investigation

### Slide 30
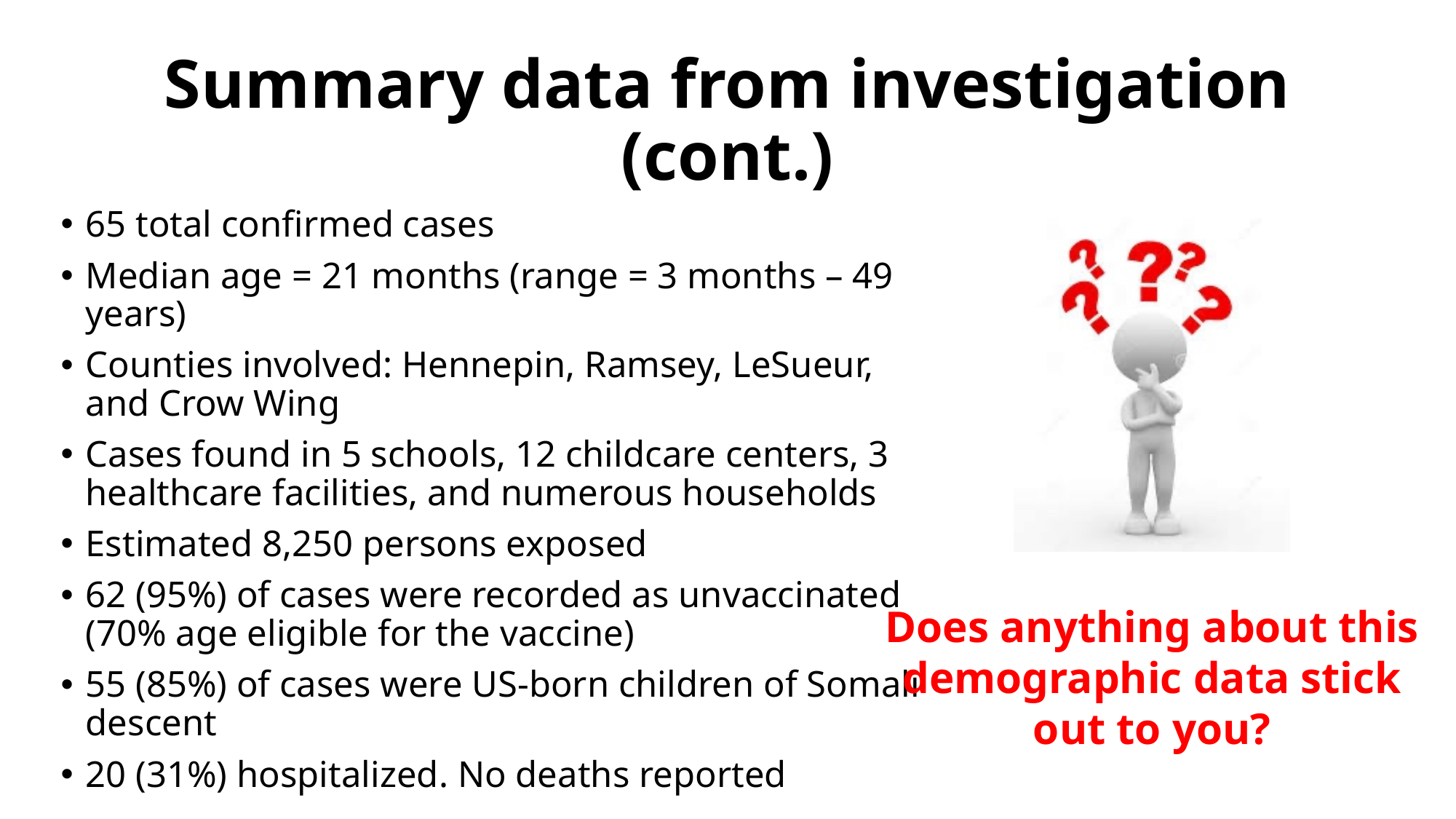

#### Summary data from investigation (cont.)
65 total confirmed cases
Median age = 21 months (range = 3 months – 49 years)
Counties involved: Hennepin, Ramsey, LeSueur, and Crow Wing
Cases found in 5 schools, 12 childcare centers, 3 healthcare facilities, and numerous households
Estimated 8,250 persons exposed
62 (95%) of cases were recorded as unvaccinated (70% age eligible for the vaccine)
55 (85%) of cases were US-born children of Somali descent
20 (31%) hospitalized. No deaths reported
Does anything about this demographic data stick out to you?

### Slide 31
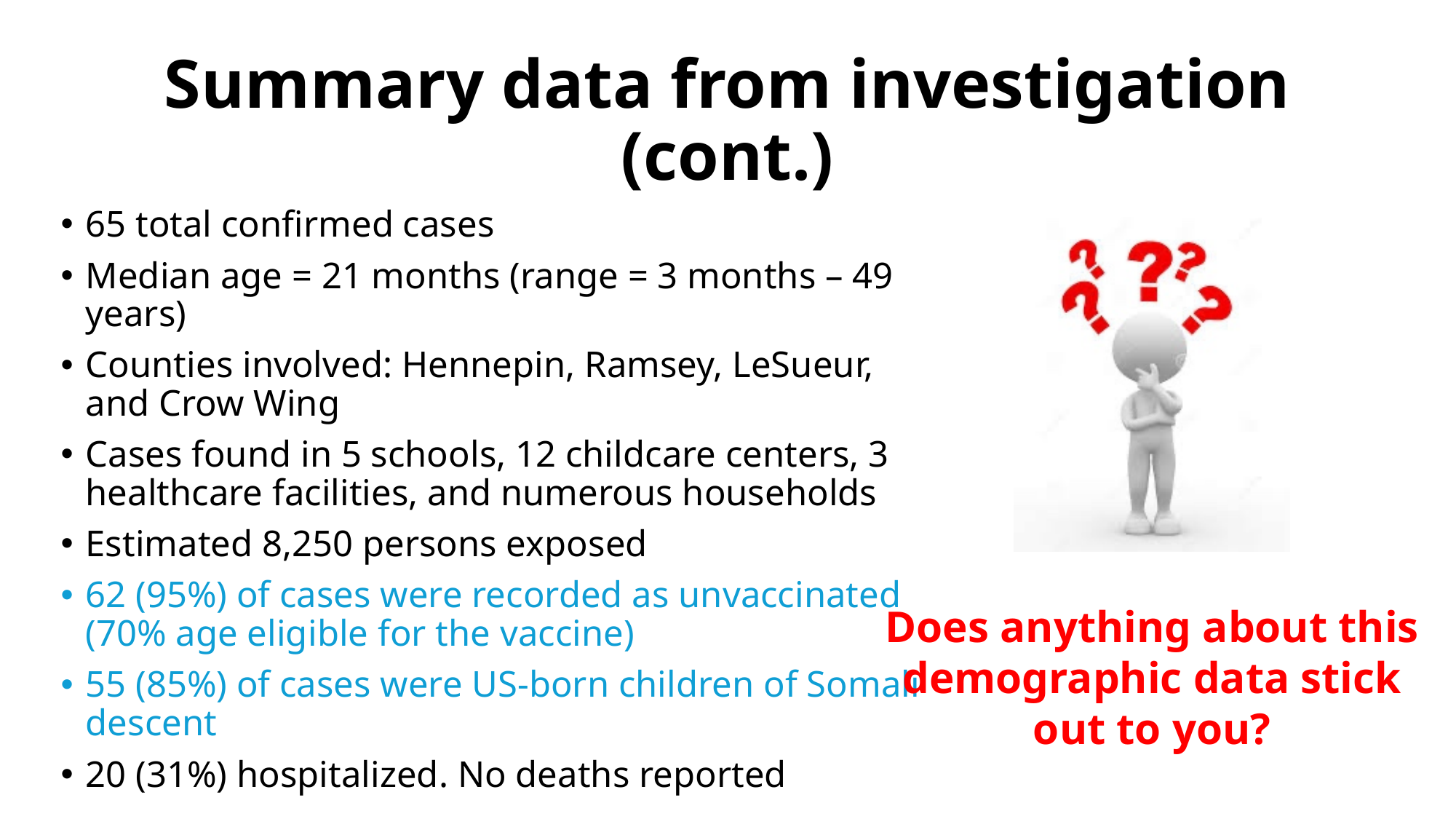

#### Summary data from investigation (cont.)
65 total confirmed cases
Median age = 21 months (range = 3 months – 49 years)
Counties involved: Hennepin, Ramsey, LeSueur, and Crow Wing
Cases found in 5 schools, 12 childcare centers, 3 healthcare facilities, and numerous households
Estimated 8,250 persons exposed
62 (95%) of cases were recorded as unvaccinated (70% age eligible for the vaccine)
55 (85%) of cases were US-born children of Somali descent
20 (31%) hospitalized. No deaths reported
Does anything about this demographic data stick out to you?

### Slide 32
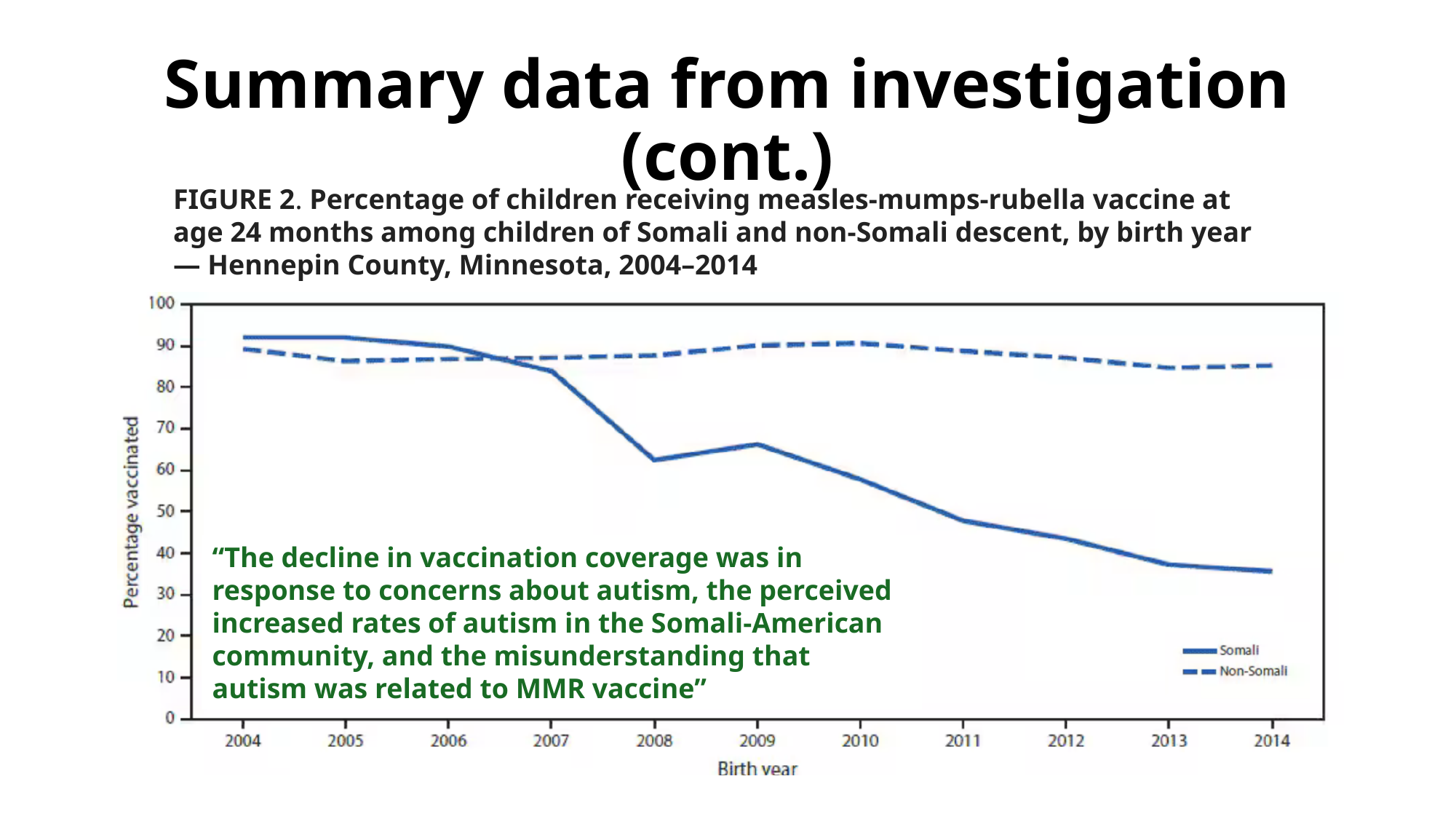

#### Summary data from investigation (cont.)
FIGURE 2. Percentage of children receiving measles-mumps-rubella vaccine at age 24 months among children of Somali and non-Somali descent, by birth year — Hennepin County, Minnesota, 2004–2014
“The decline in vaccination coverage was in response to concerns about autism, the perceived increased rates of autism in the Somali-American community, and the misunderstanding that autism was related to MMR vaccine”

### Slide 33
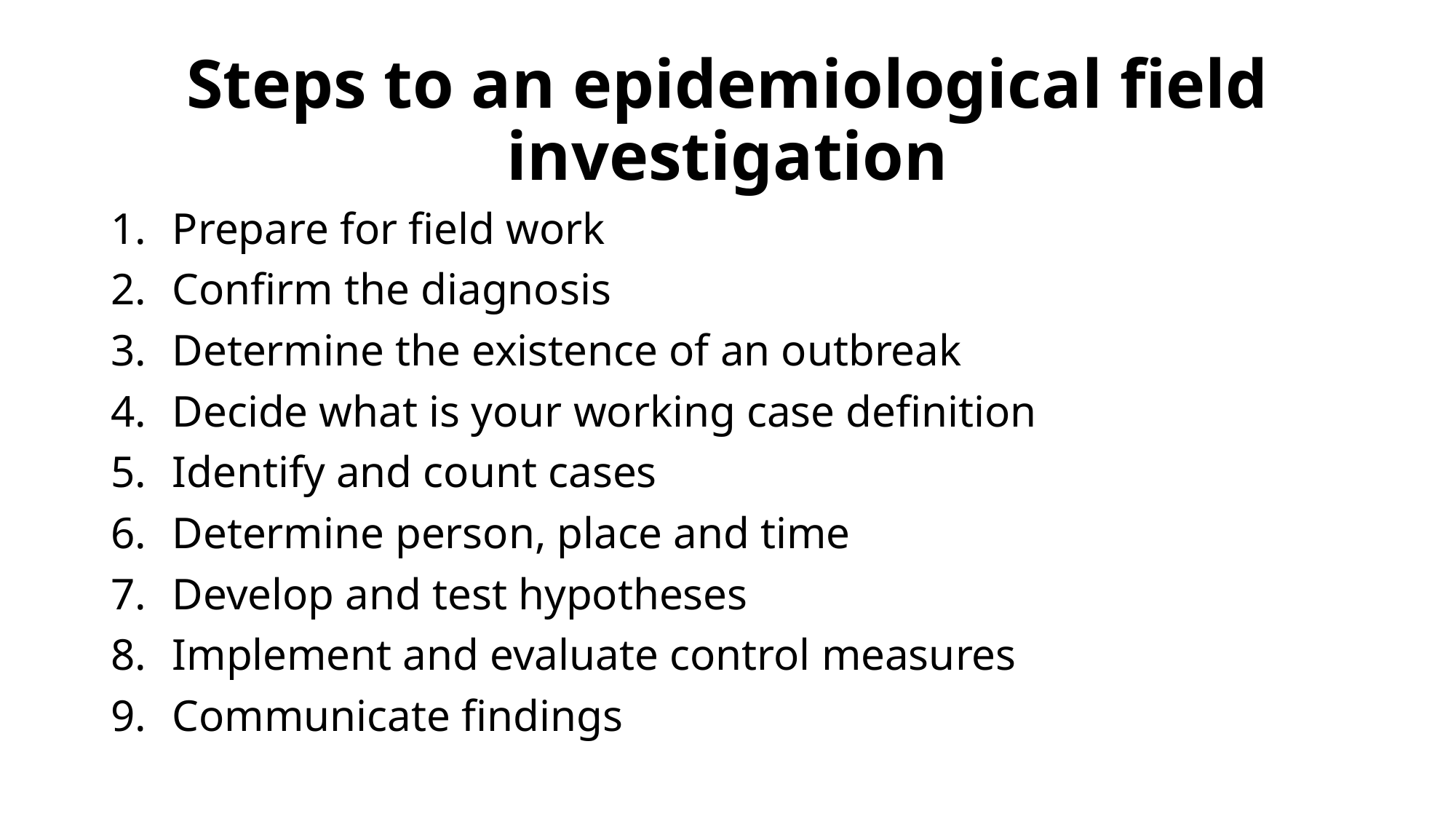

#### Steps to an epidemiological field investigation
Prepare for field work
Confirm the diagnosis
Determine the existence of an outbreak
Decide what is your working case definition
Identify and count cases
Determine person, place and time
Develop and test hypotheses
Implement and evaluate control measures
Communicate findings

### Slide 34
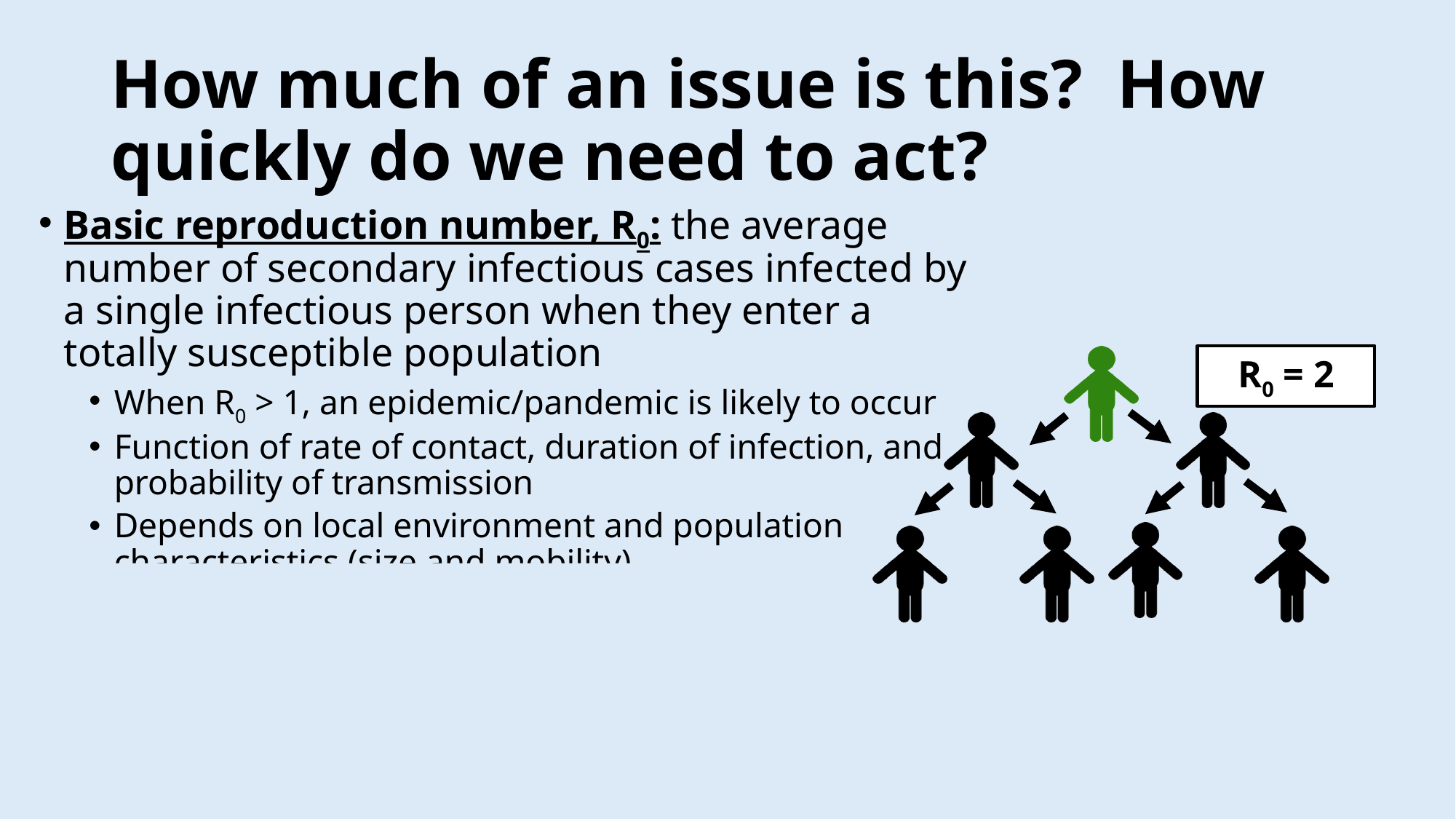

#### How much of an issue is this? How quickly do we need to act?
Basic reproduction number, R_0_: the average number of secondary infectious cases infected by a single infectious person when they enter a totally susceptible population
When R_0_ > 1, an epidemic/pandemic is likely to occur
Function of rate of contact, duration of infection, and probability of transmission
Depends on local environment and population characteristics (size and mobility)
Allows us to determine:
How quickly an infection will spread,
How big the epidemic will get without intervention
Predict when the epidemic will peak/how interventions will influence morbidity and mortality
R_0_ = 2

### Slide 35
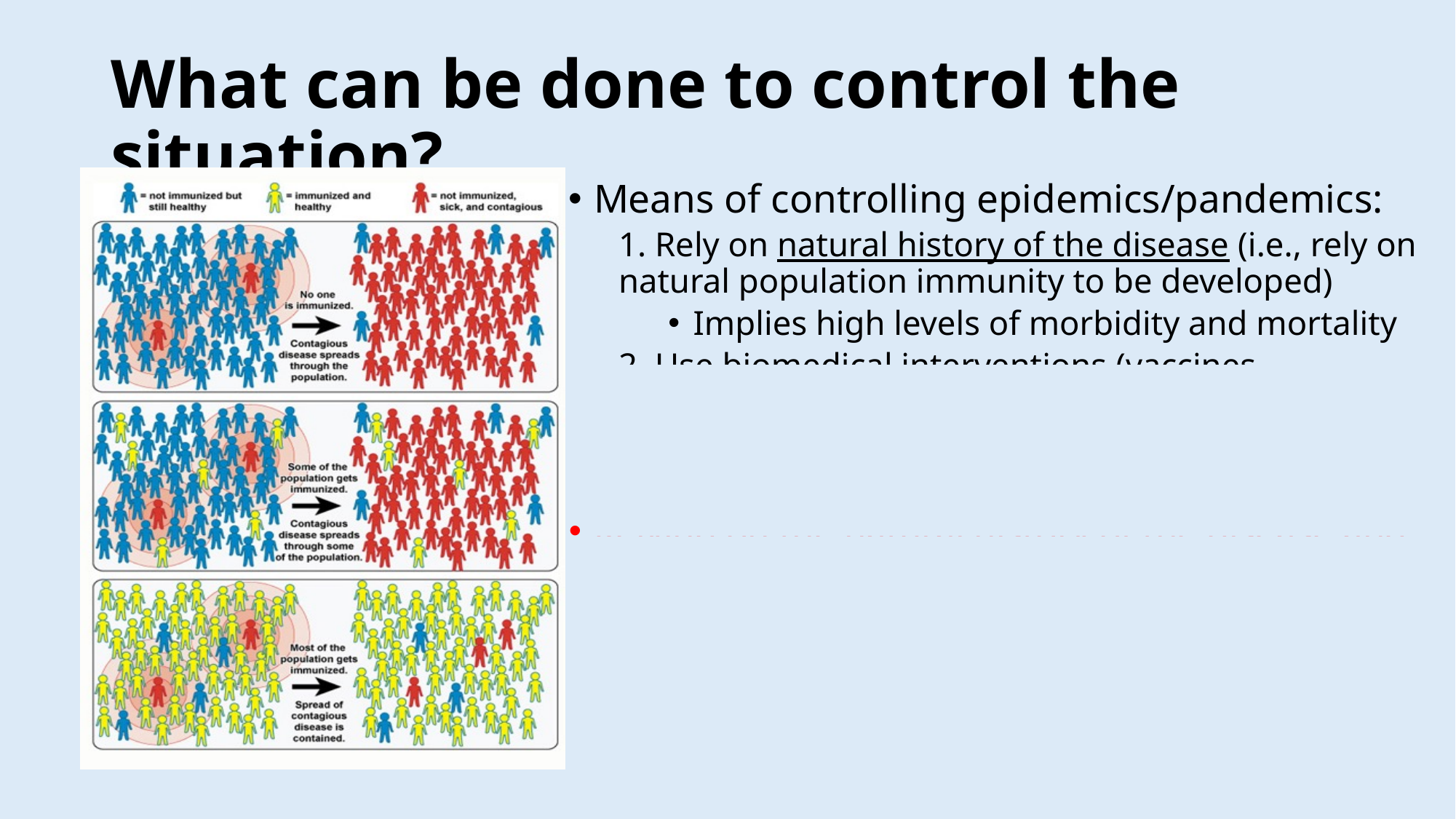

#### What can be done to control the situation?
Means of controlling epidemics/pandemics:
1. Rely on natural history of the disease (i.e., rely on natural population immunity to be developed)
Implies high levels of morbidity and mortality
2. Use biomedical interventions (vaccines, medications, etc.)
Takes time to develop in an emerging situation
Relying on the natural history of the disease and biomedical interventions (vaccines) can be used to develop herd immunity (indirect, population-level immunity to a disease)

### Slide 36
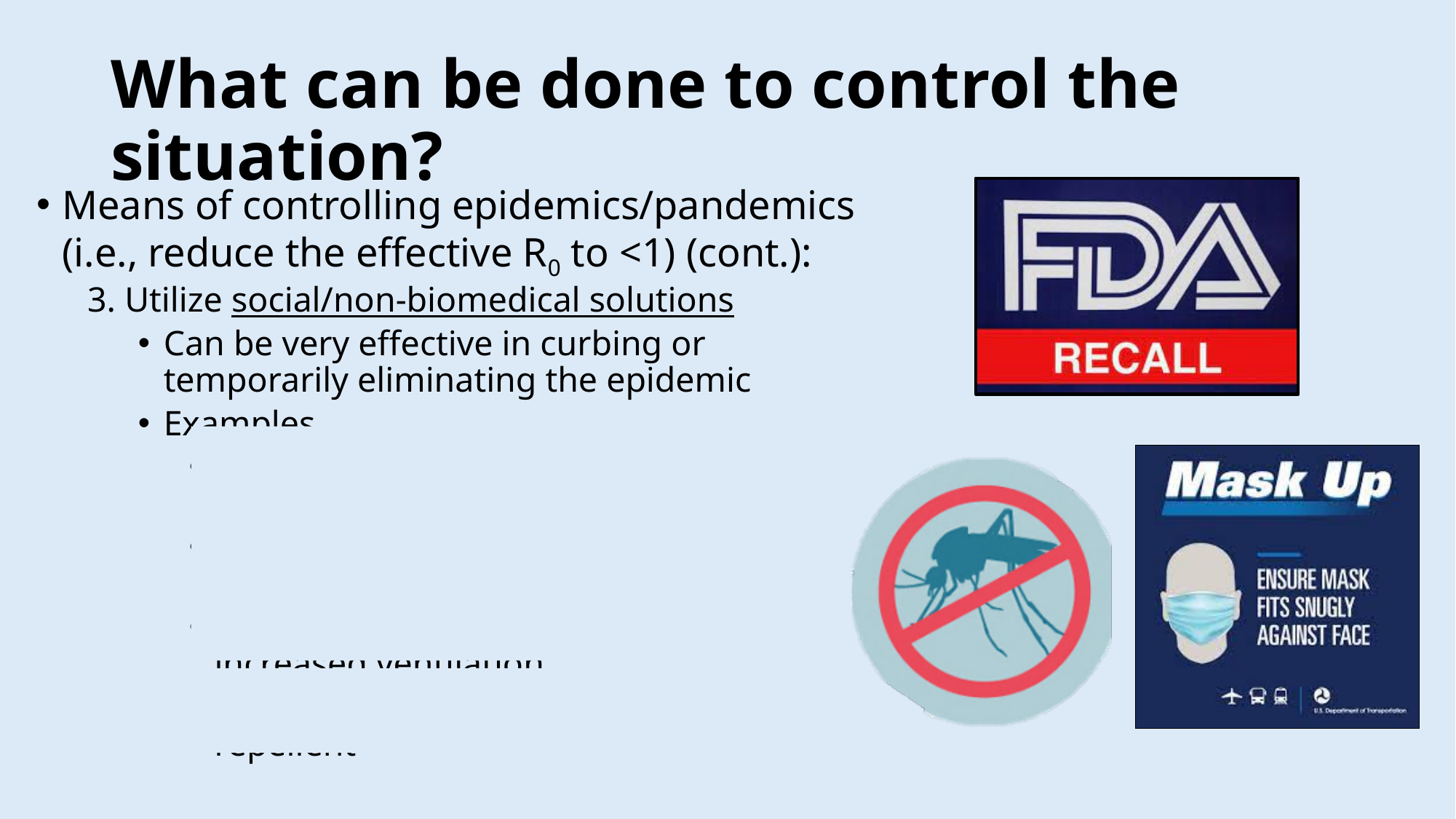

#### What can be done to control the situation?
Means of controlling epidemics/pandemics (i.e., reduce the effective R_0_ to <1) (cont.):
3. Utilize social/non-biomedical solutions
Can be very effective in curbing or temporarily eliminating the epidemic
Examples
Foodborne: Recall or eliminate infected food
Waterborne: treat contaminate water source
Airborne: social distancing, masking, increased ventilation
Vector: mass administration of insect repellent

### Slide 37
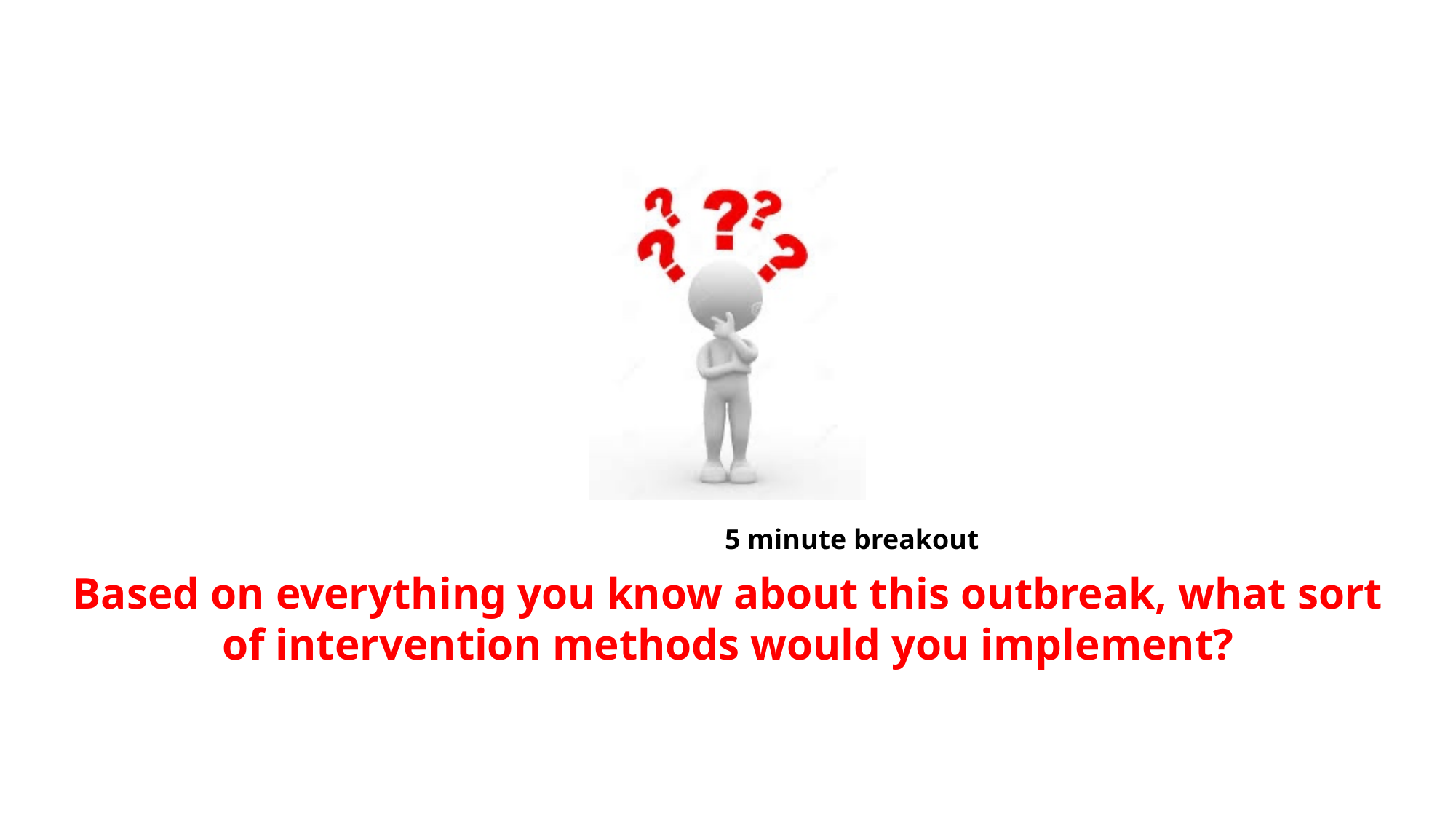

Based on everything you know about this outbreak, what sort of intervention methods would you implement?
5 minute breakout

### Slide 38
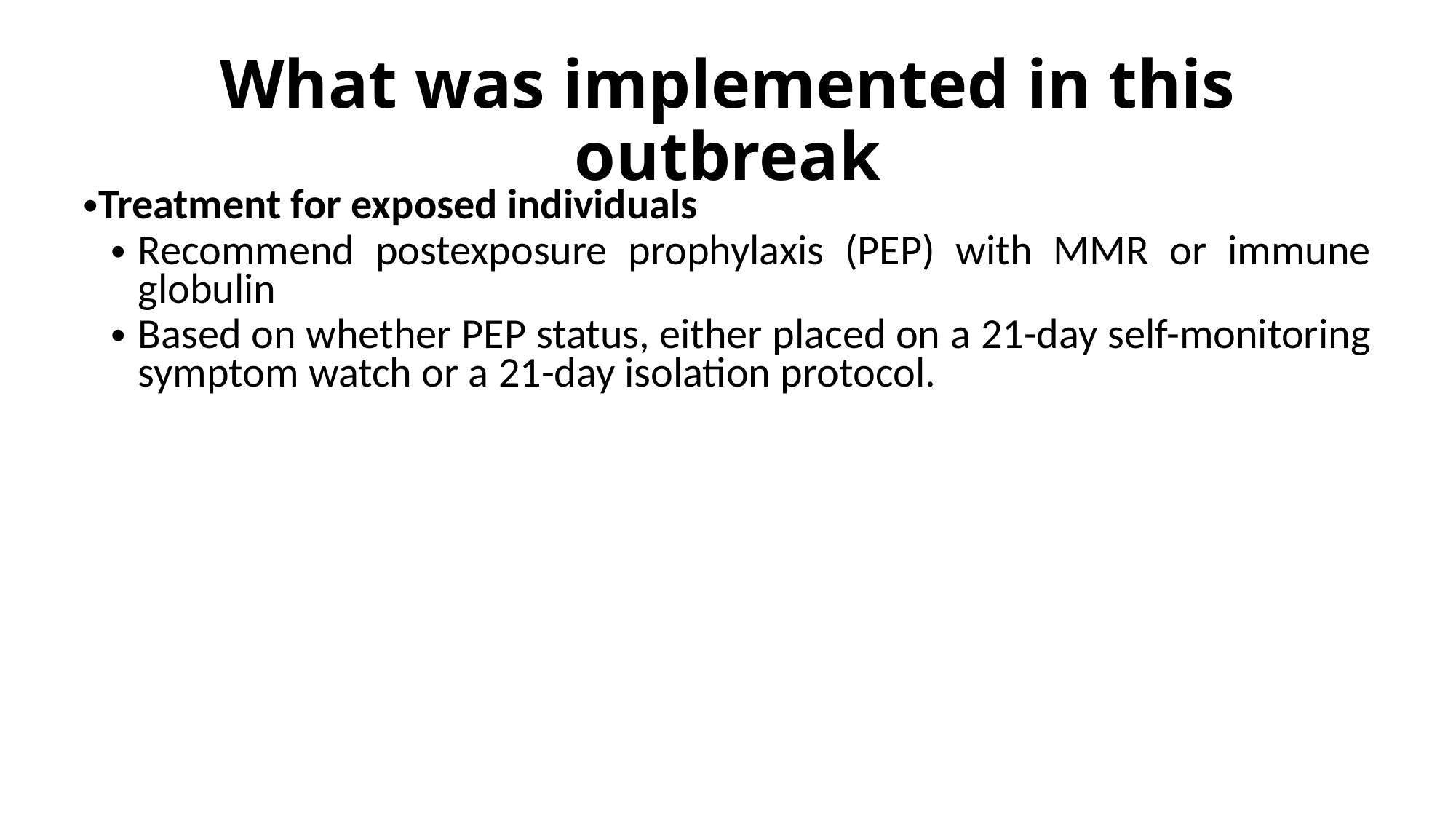

#### What was implemented in this outbreak
Treatment for exposed individuals
Recommend postexposure prophylaxis (PEP) with MMR or immune globulin
Based on whether PEP status, either placed on a 21-day self-monitoring symptom watch or a 21-day isolation protocol.
For susceptible/high risk individuals
Accelerated MMR schedule
Second dose of MMR vaccine to children who had received a first dose >28 days previously.
Initially for all children living in Hennepin County and all Minnesota Somali children regardless of county of residence (a population with known low MMR coverage rates); however, eventually recommended to all children residing in a county with at least one case was identified

### Slide 39
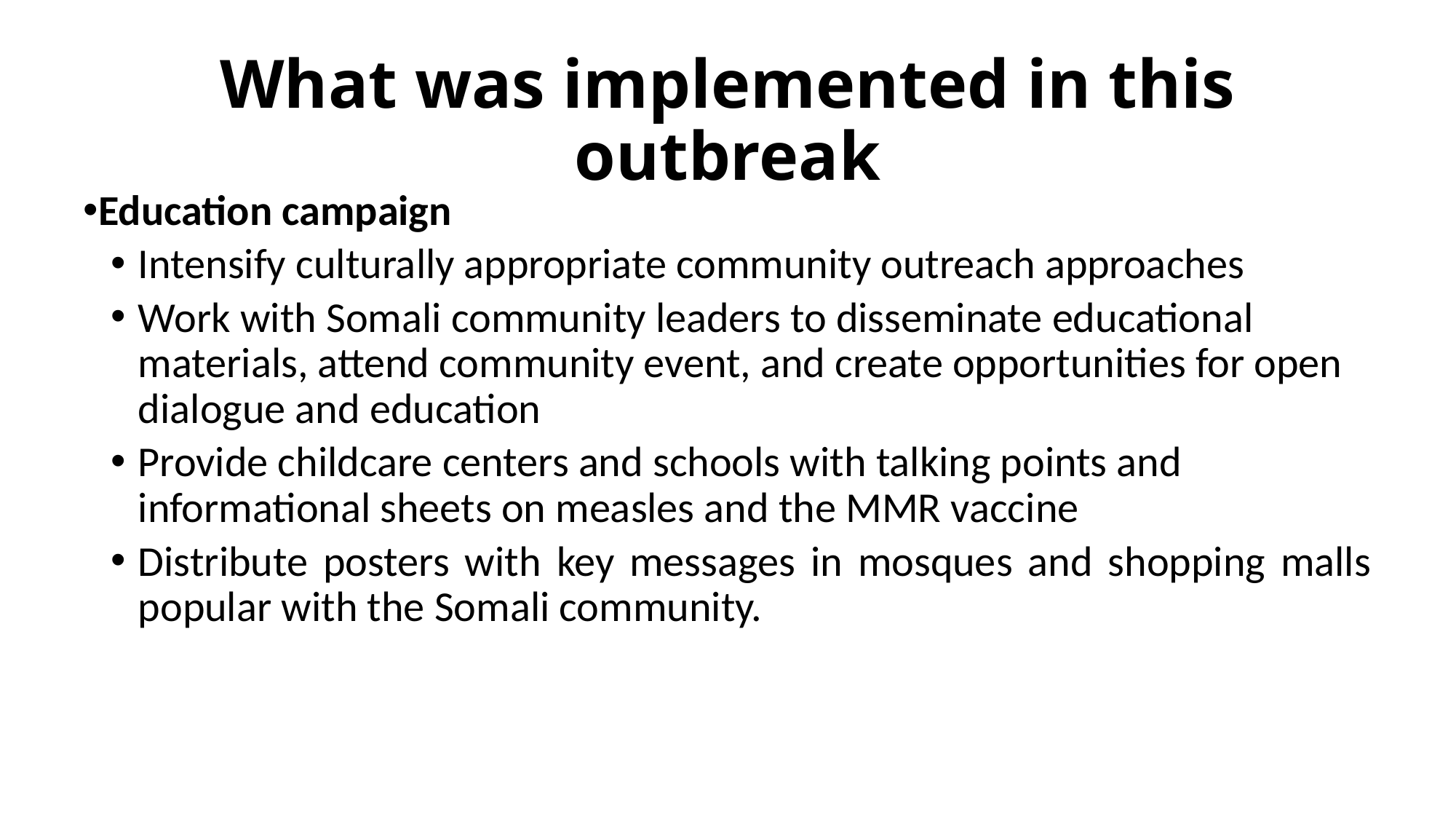

#### What was implemented in this outbreak
Education campaign
Intensify culturally appropriate community outreach approaches
Work with Somali community leaders to disseminate educational materials, attend community event, and create opportunities for open dialogue and education
Provide childcare centers and schools with talking points and informational sheets on measles and the MMR vaccine
Distribute posters with key messages in mosques and shopping malls popular with the Somali community.

### Slide 40
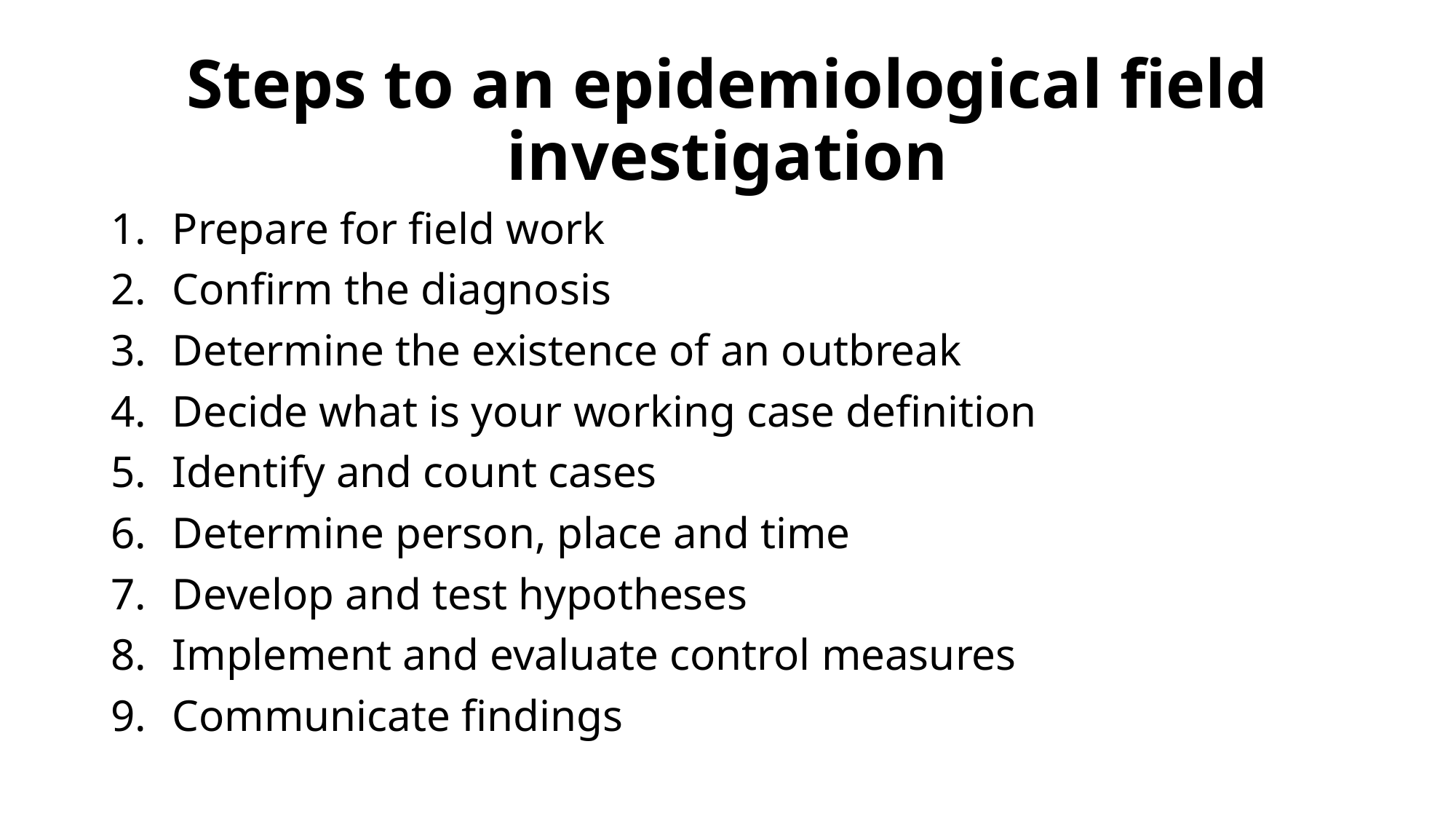

#### Steps to an epidemiological field investigation
Prepare for field work
Confirm the diagnosis
Determine the existence of an outbreak
Decide what is your working case definition
Identify and count cases
Determine person, place and time
Develop and test hypotheses
Implement and evaluate control measures
Communicate findings

### Slide 41
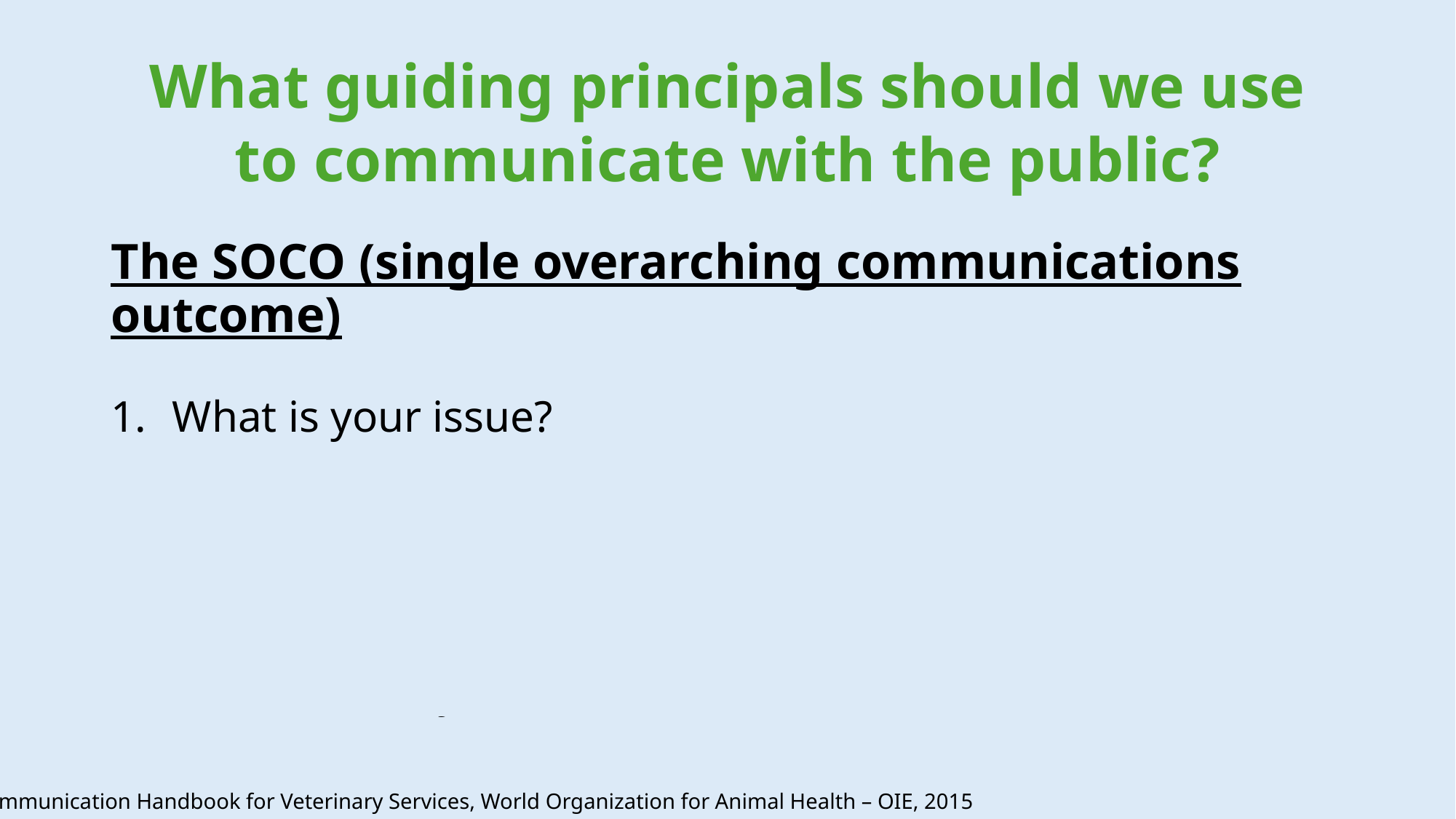

#### What guiding principals should we use to communicate with the public?
The SOCO (single overarching communications outcome)
What is your issue?
Why do you want to focus on this issue and why do you want to focus on it now?
Who needs to change their behavior (i.e., audience)?
What is the change that you want to see in your audience as a result of your communication?
Communication Handbook for Veterinary Services, World Organization for Animal Health – OIE, 2015

### Slide 42
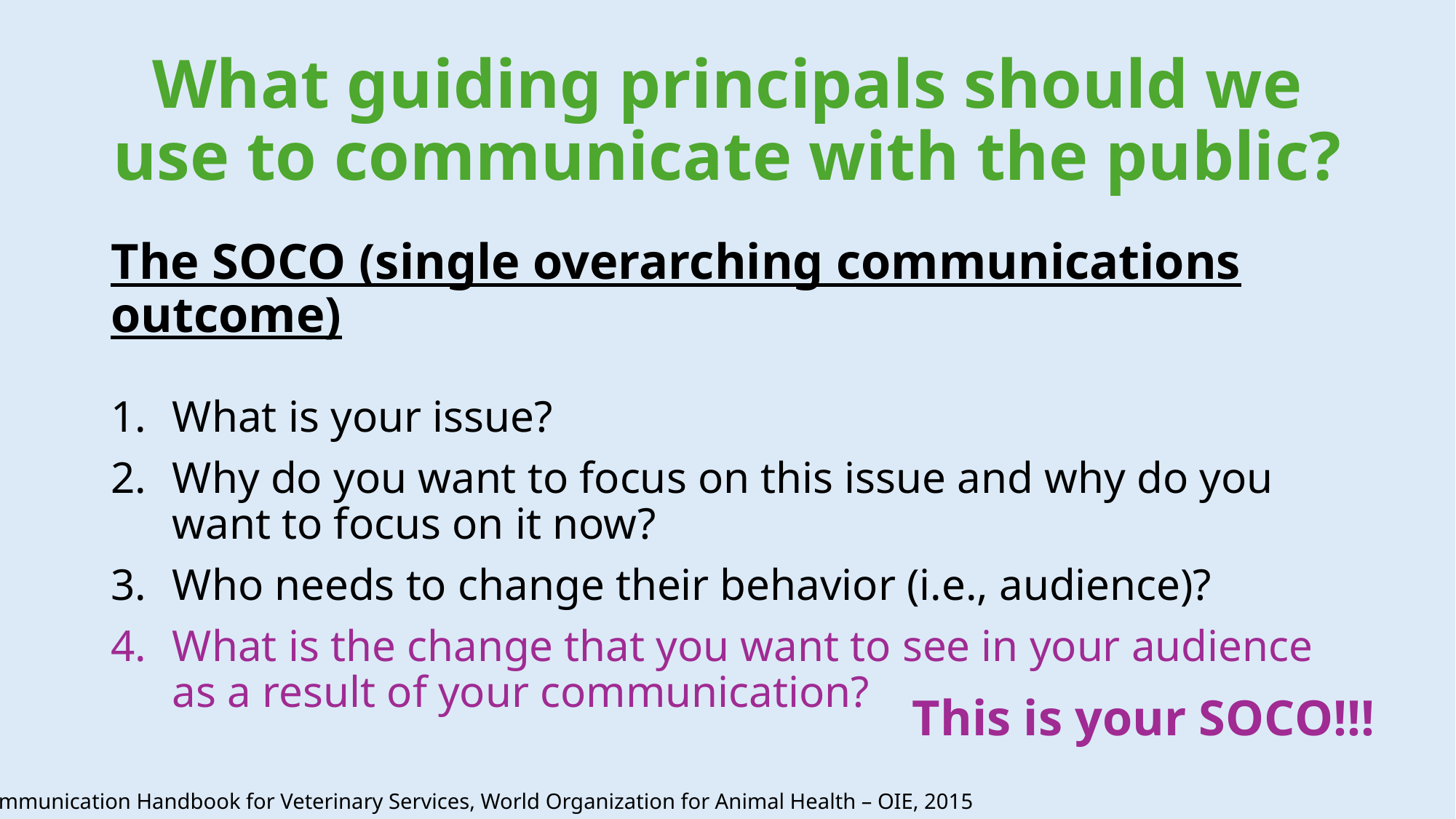

#### What guiding principals should we use to communicate with the public?
The SOCO (single overarching communications outcome)
What is your issue?
Why do you want to focus on this issue and why do you want to focus on it now?
Who needs to change their behavior (i.e., audience)?
What is the change that you want to see in your audience as a result of your communication?
This is your SOCO!!!
Communication Handbook for Veterinary Services, World Organization for Animal Health – OIE, 2015

### Slide 43
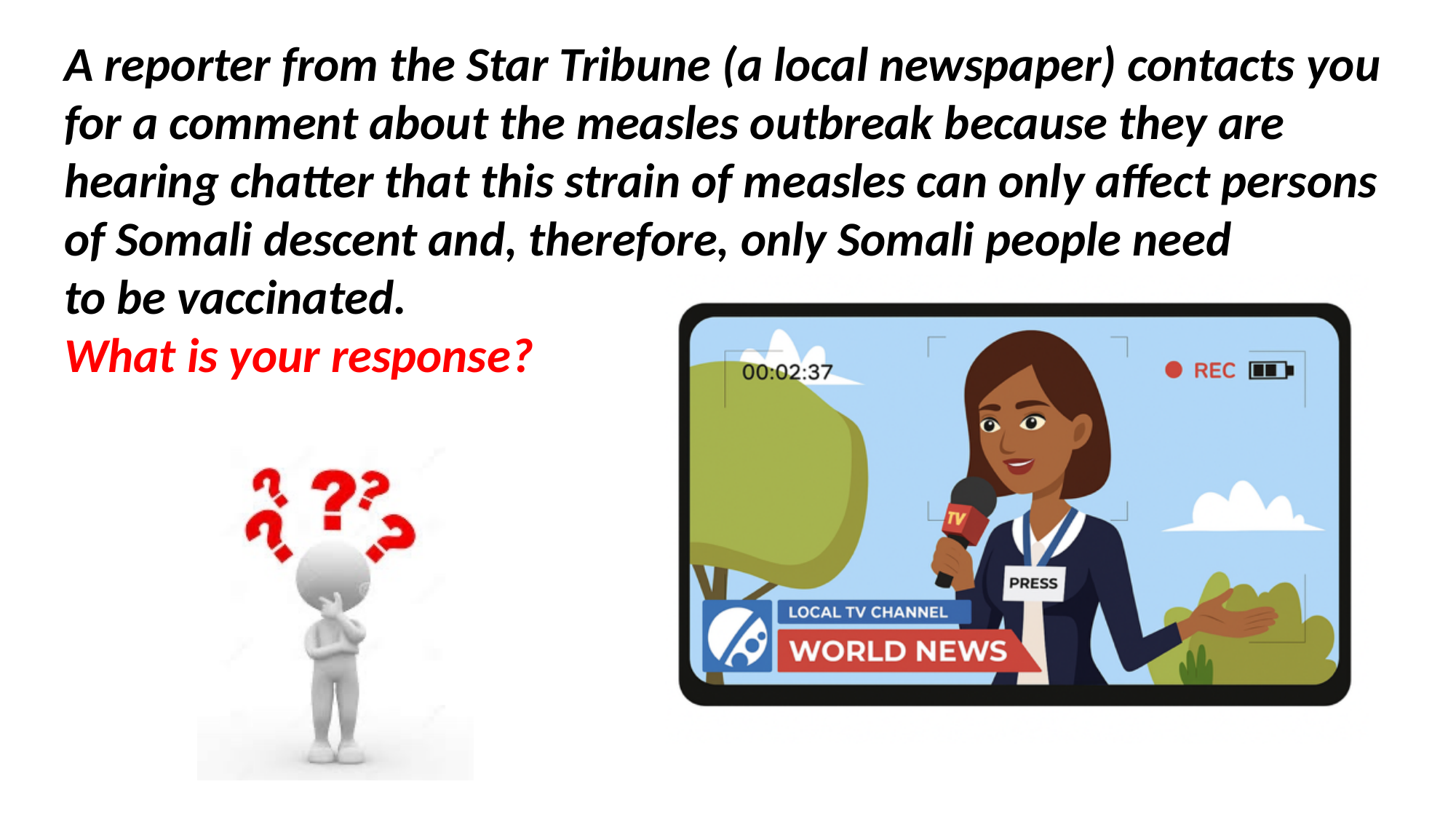

A reporter from the Star Tribune (a local newspaper) contacts you for a comment about the measles outbreak because they are hearing chatter that this strain of measles can only affect persons of Somali descent and, therefore, only Somali people need
to be vaccinated.
What is your response?

### Slide 44
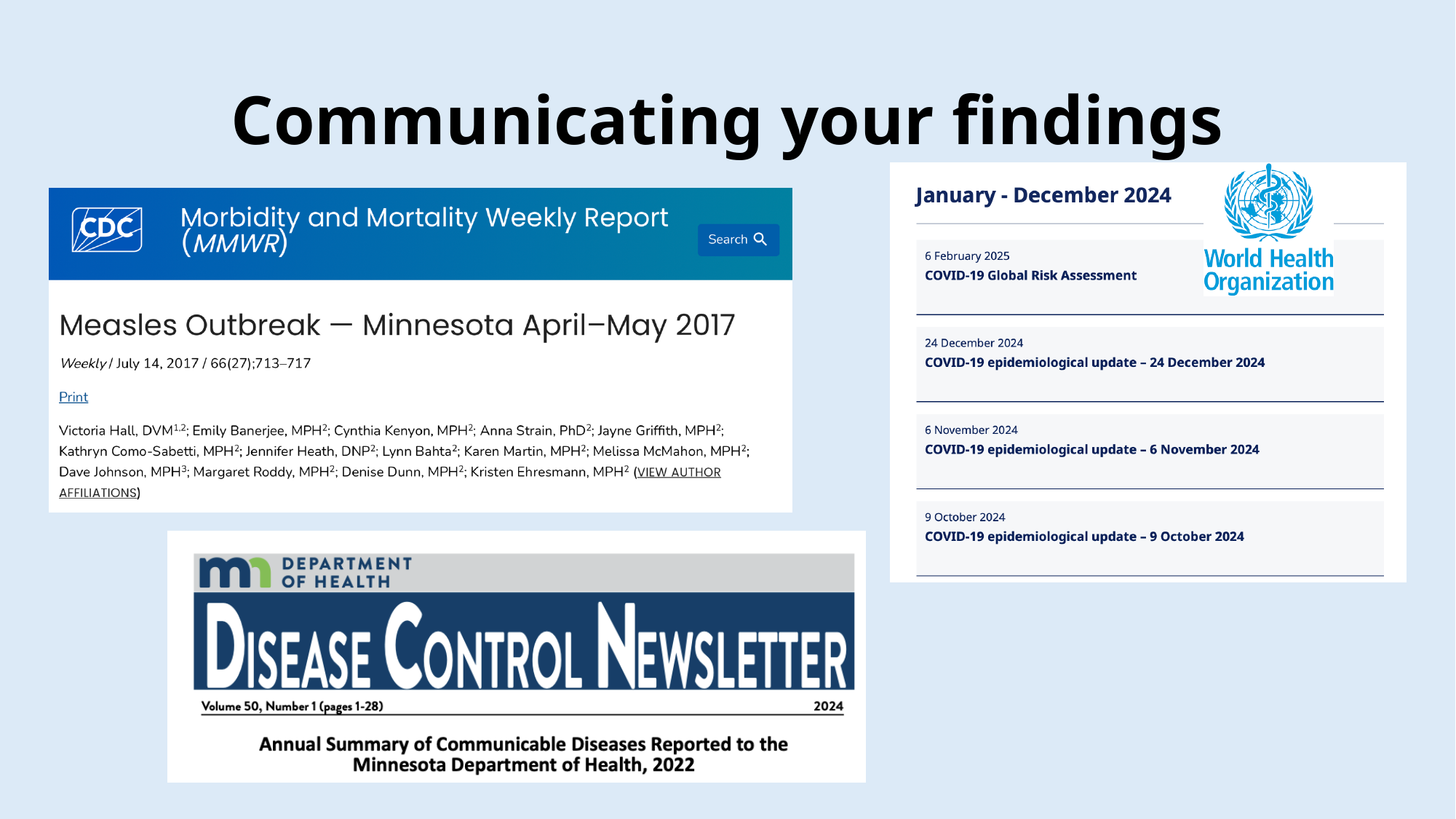

#### Communicating your findings

### Slide 45
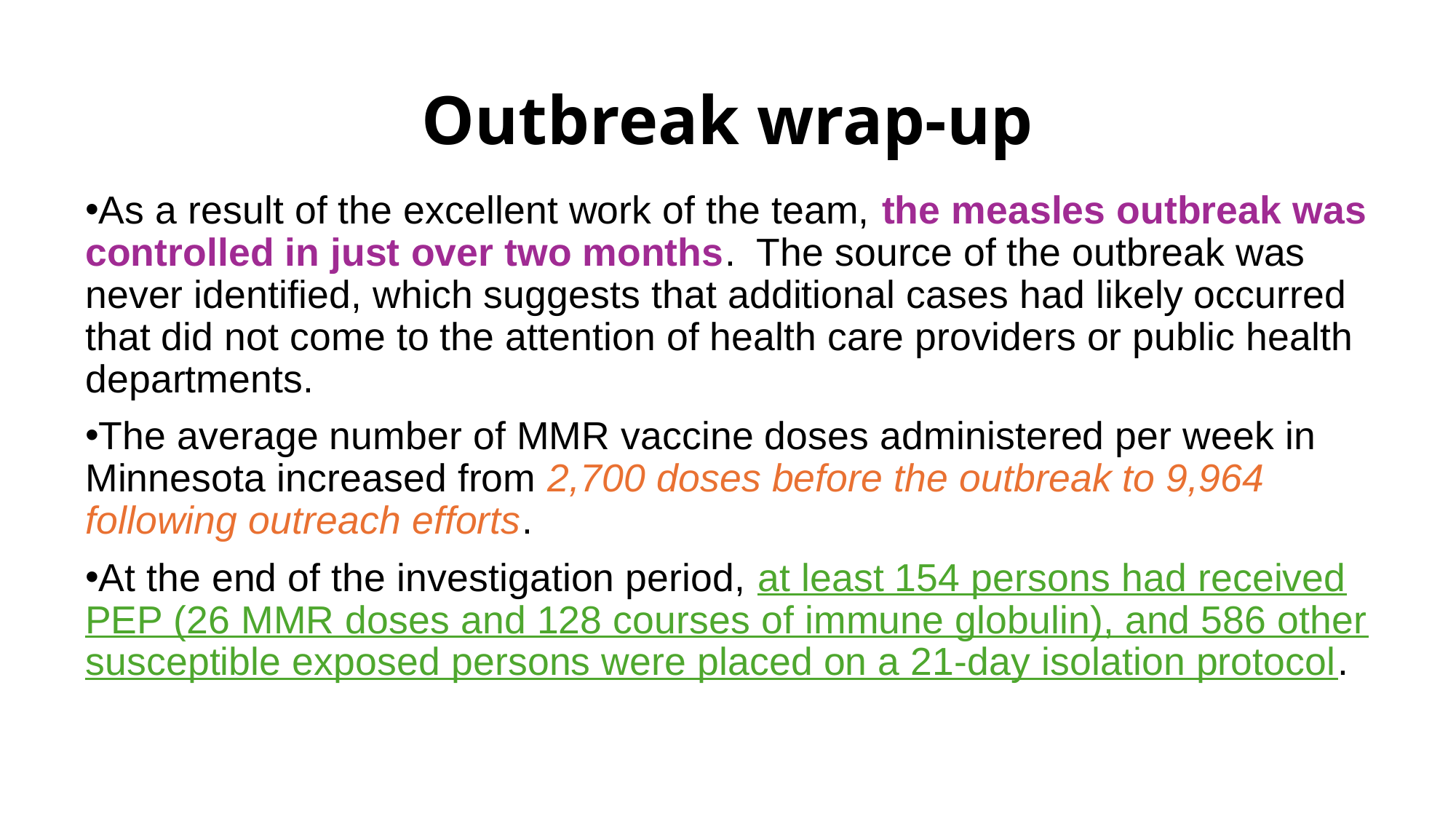

#### Outbreak wrap-up
As a result of the excellent work of the team, the measles outbreak was controlled in just over two months. The source of the outbreak was never identified, which suggests that additional cases had likely occurred that did not come to the attention of health care providers or public health departments.
The average number of MMR vaccine doses administered per week in Minnesota increased from 2,700 doses before the outbreak to 9,964 following outreach efforts.
At the end of the investigation period, at least 154 persons had received PEP (26 MMR doses and 128 courses of immune globulin), and 586 other susceptible exposed persons were placed on a 21-day isolation protocol.

### Slide 46
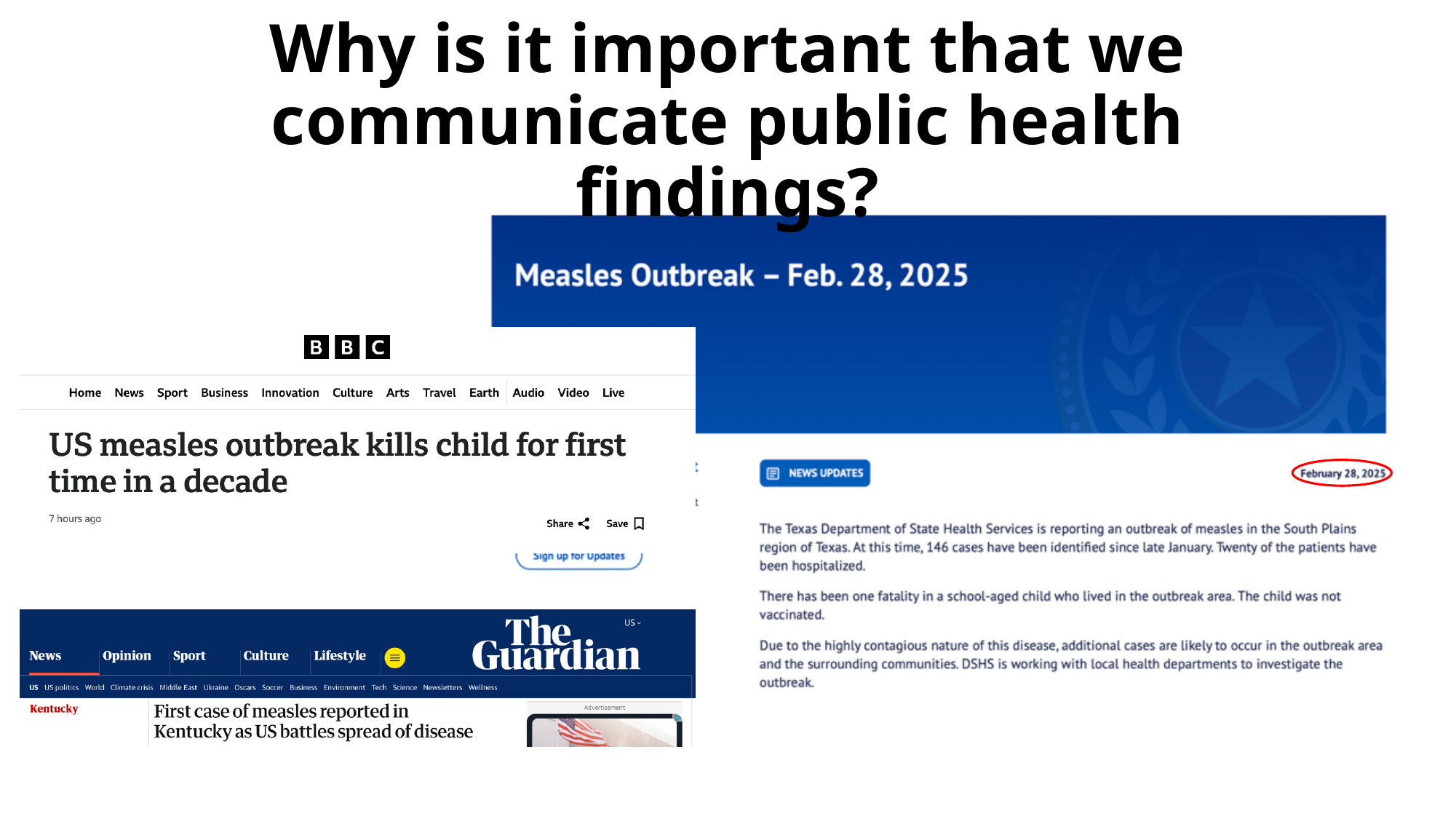

#### Why is it important that we communicate public health findings?

### Slide 47
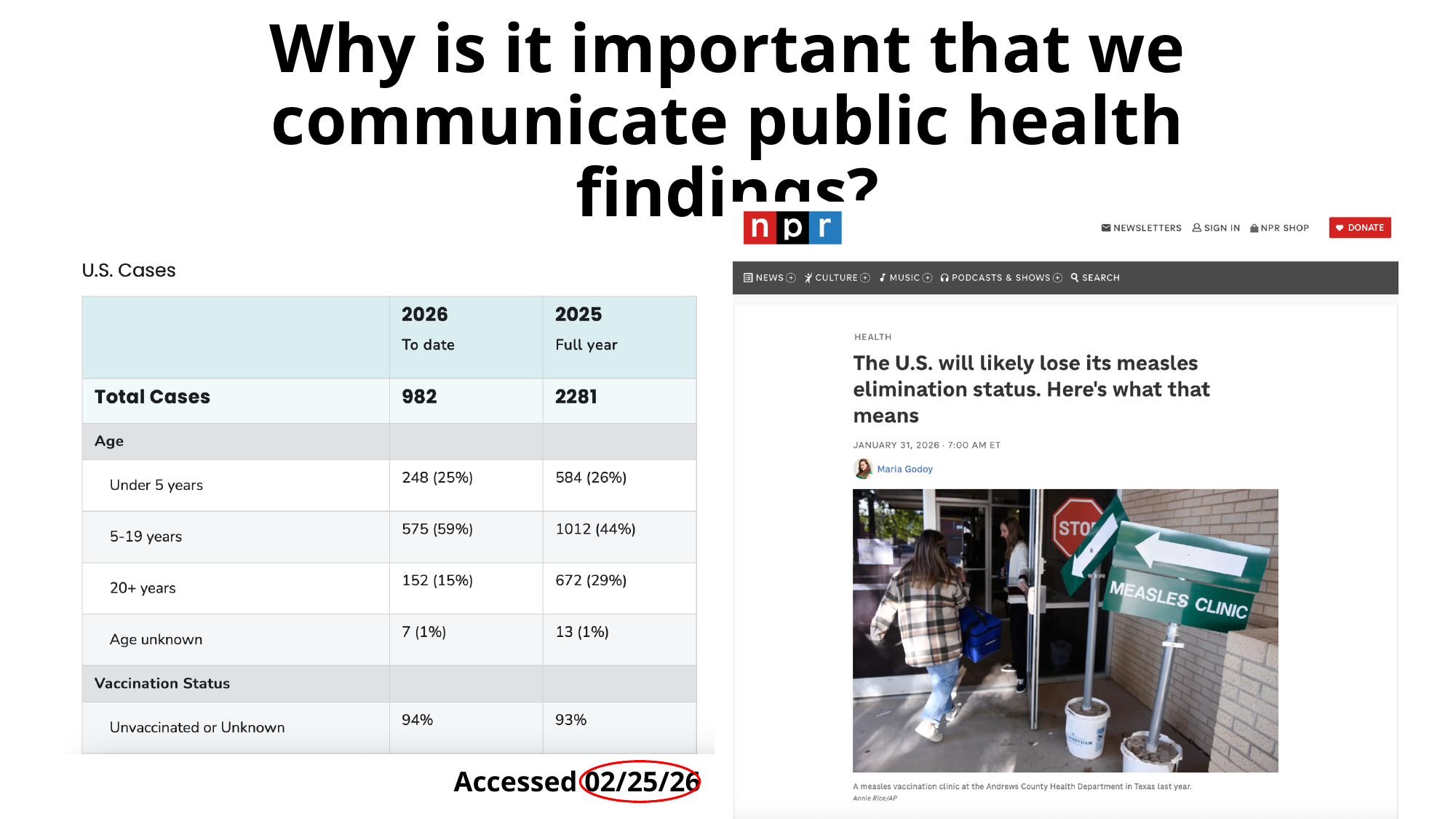

#### Why is it important that we communicate public health findings?
Accessed 02/25/26

### Slide 48
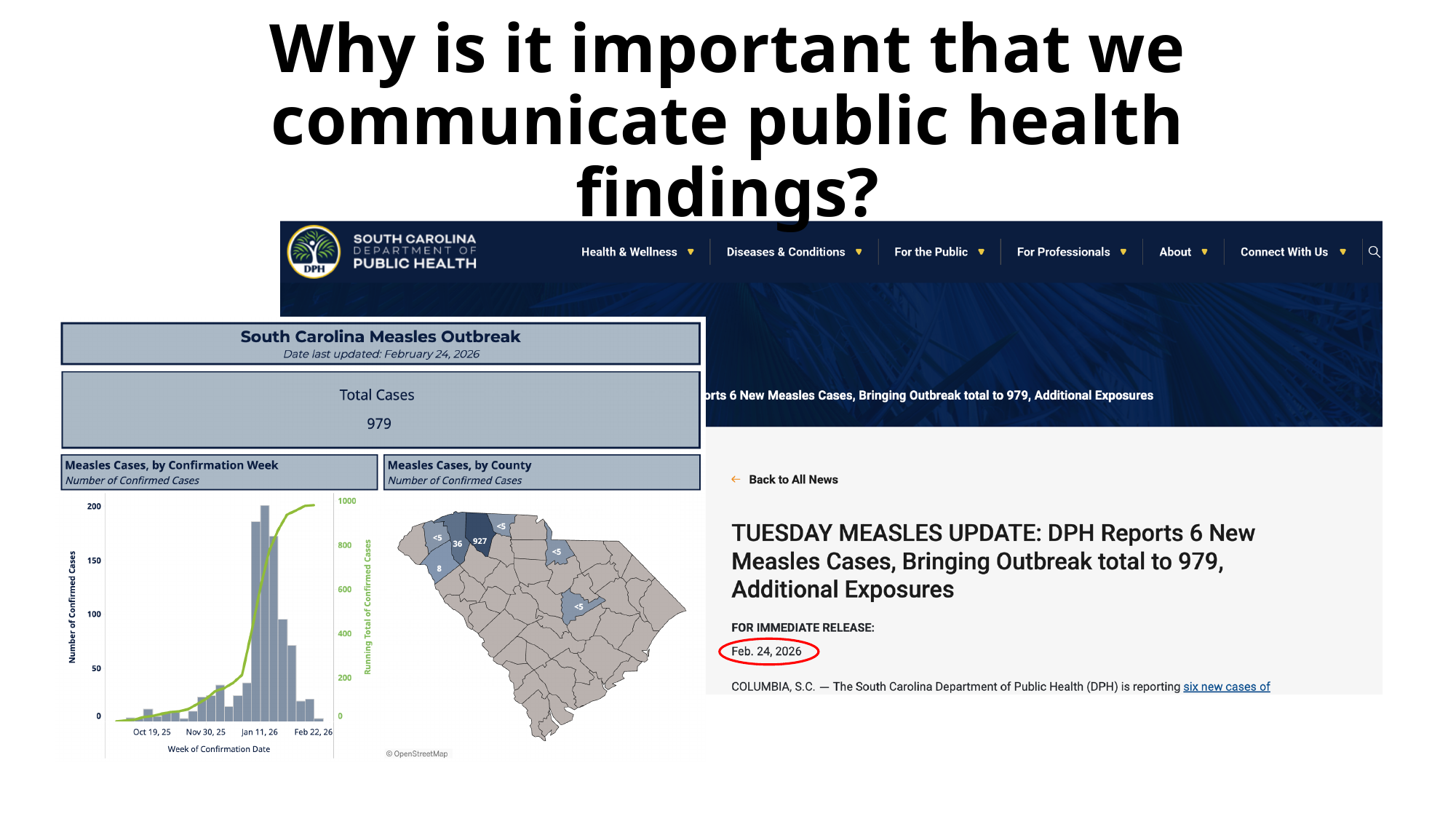

#### Why is it important that we communicate public health findings?
