## Supplementary material for "Bridging microbiology and public health through simulation-based learning": This supplement contains the outbreak notes worksheet students should download or print prior to the class session.

### Field Epidemiology Notes

Pathogen of interest: ________________________

Notes about the pathogen:

1. **Prepare for fieldwork**

   Preliminary findings from local health department about emergent public health situation:

   Have you, as an external consultant, been formally invited by the local health department? Yes/No

2. **Confirm the diagnosis**

   Known information about hypothesized pathogen of interest within this emergent public health situation:

   Preliminary control recommendations:

3. **Determine the existence of an outbreak**

   Classification of emergent public health situation:

#### 4. Identify and count cases

Case definition:

a. Person:

b. Place:

c. Time:

d. Clinical features:

Potential locations for case ascertainment:

#### 5. Determine person, place, and time

Information learned about emergent public health situation:

#### 6. Develop and test hypotheses

Current hypothesis about cause of emergent public health situation:

#### 7. Implement and evaluate control measures

Updated control recommendations:

#### 8. Communicate findings

Control messaging for media:

a. What is your issue?

b. Why do you want to focus on this issue and why do you want to focus on it now?

c. Who needs to change their behavior (i.e., audience)?

d. What is the change that you want to see in your audience as a result of your communication?

e. What is your SOCO?

*CDC's Morbidity and Mortality Weekly Report (MMWR):* https://www.cdc.gov/mmwr/index.html
*WHO's Disease Outbreak News (DONs):* https://www.who.int/emergencies/disease-outbreak-news
