## Supplementary material for "Bridging microbiology and public health through simulation-based learning": This supplement contains the additional follow-up information students should be provided following the class session.

### ***Additional Resources***

*If you found the information covered in this module interesting and would like to learn more, here are some additional resources on epidemiology and outbreak investigation.*

---

**MMWR Report from Case Study in Lecture Material:**

- Link to report (Links to an external site.)

---

**Outbreak Investigation Materials:**

- CDC "Solve the Outbreak" Game (Links to an external site.)
- CDC Guide to Outbreak Investigation (Links to an external site.)
- CDC List of Current Outbreaks (Links to an external site.)

---

**Public Health Agency Websites:**

- CDC Website (Links to an external site.)
- WHO Website (Links to an external site.)
- FDA Website (Links to an external site.)
- Michigan HHS Website (Links to an external site.)

---

**Resources for Interest Future Studies in Public Health and Epidemiology:**

- American Public Health Association Website (Links to an external site.) - a great resource for an overview of the field and different facets of it
- Association of Schools and Programs of Public Health Website (Links to an external site.)
- UM School of Public Health Program Website (Links to an external site.)

---

**Other Links:**

- Our World in Data (Links to an external site.) - open-source data explorer which covers many public health topics (including infectious diseases such as COVID-19, diarrheal disease, mortality causes, and environmental determinants of health)
